# Effects of Maternal stress on epigenetic markers on fetal tissues and neurodevelopment at various gestational ages: A Systematic review

**DOI:** 10.1101/2025.04.05.25325300

**Authors:** Shabana Jassim, Shaheen Basheer

## Abstract

The Developmental Origins of Health and Disease (DOHaD) theory hypothesized that environmental exposures during early life (particularly the in-utero period) can permanently influence health and vulnerability to disease in later life. This intriguing concept has been the subject matter of great interest in recent research, as much vital development of fetal neuronal networks and brain cell division happens in fetal life. Fetal programming, suggests that the in-utero environment is now increasingly believed to have a great impact on long term health and pattern of disease development in later life. There is also some evidence to show transgenerational epigenetic programming of genes. Various biological mechanisms are likely to be involved in fetal programming, epigenetic modifications being just one of them. This systematic study aims to review currently available primary research data to assess the significance of maternal stress in epigenetic modification affecting fetal neurodevelopment as assessed from fetal or maternal tissue sampling.

**Rationale:** Most studies have focused on impact by a particular type of stress. While some studies have studied long term outcomes, into psychiatric and intellectual follow up in adults, few have actually researched the direct effect of maternal stress or trauma on the fetus, and even fewer have documented direct sampling of genetic markers as a result of the same. There is a significant knowledge gap on how to link or classify the epigenetic impact of various kinds of maternal stress on fetal development. This systematic review hopes to evaluate the available evidence that may help us better understand the significance various types of maternal stress during pregnancy and of its diverse epigenetic influence on the developing fetus.

**Objective:** To assess and review the current available research in evaluating the role of epigenetics of the influence of maternal stress during human pregnancy on the neurodevelopment of the developing fetus as evidenced by tissue analysis.

**Methods:** A systematic review was conducted on available literature from online databases and reported following the Preferred Reporting Items for Systematic Reviews and Meta-Analysis 2020 checklist. A comprehensive search strategy was developed. Only Observational and interventional studies, which were ‘ primary research studies’ were included in assessing epigenetic markers derived from fetal or maternal tissue in conditions of maternal stress during pregnancy and were analyzed. We retrieved 492 papers most relevant to the query, mostly from PubMed, Semantic search tools and Journals. Both authors rigorously studied and summarized the abstracts to exclude the studies that did not meet the outlined criteria, even though well planned comprehensive studies were found, which lacked key inclusion criteria. Each study was further analysed for specific type of stress, gestational age (temporal characteristics), tissue sample analysed. Forty studies were shortlisted after screening and selected for the review, based on the inclusion criteria.

Only human studies, that use validated psychological or physiological measures to assess maternal stress and analyze epigenetic markers in fetal or maternal tissue were included and also must specify the gestational timing of stress exposure. Only observational, cohort, or case-control primary studies that clearly defines and categorizes the type of stress (psychological, physical, or environmental) were included.

Non-human studies, or Duplicate publications and studies that were missing key information related to the question asked were excluded.

Studies were screened by the two reviewers independently. Risk of bias was assessed using RoB 2.0 tool.

**Data was analyzed** and tabulated descriptively, using the data extraction tool designed for the study. Data extraction table is attached at the end of the article

Missing data were not analysed.

**Results:** This systematic review of literature spanning at least 40 independent primary studies, involving around 19,400 mothers indicated that different maternal stress types yield distinct epigenetic signatures in tissues linked to fetal development. Factors such as chronic psychological stress, traumatic exposure and symptoms of anxiety or depression are each associated with specific patterns of DNA methylation. Stress response genes, especially NR3C1, FKBP5 and HSD11B2, consistently show altered methylation. One investigation of maternal smoking documented global DNA methylation changes in fetal brain tissue, whereas most studies examined placenta, cord blood, or buccal cells.

In early gestations, it was found that maternal depression, perceived stress, and adverse life events relate to altered methylation of NR3C2, MEST, and other markers enriched for neurodevelopmental functions. In contrast, maternal stress during later trimesters was associated with changes in NR3C1, FKBP5, BDNF, and related genes, sometimes also exhibiting sex-specific effects. Traumatic and especially war-related stress yields robust increases in NR3C1 methylation that may persist into later life. These findings support that the type and timing of maternal stress selectively drives epigenetic modifications in gene networks critical to neuroendocrine function and neural development.

**Conclusion:** The timing and type of maternal stress determines which specific genes undergo methylation changes in feto maternal tissue, with early and late gestational periods showing distinct epigenetic signatures. Although several key similarities were noted, there were some conflicting results, which may be attributable to limited sample size or heterogeneity between studies. Further research with more robust standardization and more large scale randomized studies is needed to validate the exact mechanism and impact on epigenetics due to maternal stress.

## Introduction

Prenatal maternal stress refers to the stress that a mother experiences during her pregnancy. Prenatal stress can be chronic, linked to ongoing events in a woman’s life if acute, linked to sudden changes in a woman’s daily routine or sudden change in the environment or surroundings. It may range from perceived stress, just due to a new pregnancy, to circumstances in the family, lack of support, low socioeconomic status, psychiatric illness, racial discrimination, intimate partner violence, smoking, environmental pollution, natural disasters or conditions of war. Post-traumatic stress disorder during pregnancy has been known to increase the risk of high blood pressure having preterm birth or a low-birth-weight infant.<u>1</u>. PTSD and depression also increase the risk for behaviours such as smoking and drinking, which contribute to other problems.

In-utero exposure to maternal psychological distress is increasingly linked with disrupted fetal and neonatal brain development and long-term neurobehavioral dysfunction in children and adults. Maternal psychological distress during pregnancy is associated with changes in fetal brain structure and function, including reduced hippocampal and cerebellar volumes, increased cerebral cortical gyrification and sulcal depth, decreased brain metabolites (e.g., choline and creatine levels), and disrupted functional connectivity(Wu, Y. et al). After birth, reduced cerebral and cerebellar gray matter volumes, increased cerebral cortical gyrification, altered amygdala and hippocampal volumes, and disturbed brain microstructure and functional connectivity have been reported in the offspring months or even years after exposure to maternal distress during pregnancy. Additionally, adverse child neurodevelopment outcomes such as cognitive, language, learning, memory, social-emotional problems, and neuropsychiatric dysfunction are being increasingly reported after prenatal exposure to maternal distress.

The mechanisms by which prenatal maternal psychological distress influences early brain development include but are not limited to impaired placental function, disrupted fetal epigenetic regulation, altered microbiome and inflammation and dysregulated hypothalamic pituitary adrenal axis.

Epigenetics are inheritable alterations to genetic expression that, without affecting the genome itself (2), modulate the expression of a genotype through DNA methylation, histone modifications, and non-coding RNA. The physical structure of the DNA molecule can be transformed by alterations in DNA and histone methylation, phosphorylation, and acetylation. Non-coding RNA, such as microRNA and small interfering RNA (siRNA), impact the expression of key epigenetic regulators such as DNA methyltransferase, recruit chromatin modifiers, and attract RNA polymerase to specific genes (3, 4). As a result, epigenetic modifications play a pivotal role in cellular differentiation and may silence or activate key genes at the cellular or organism level.

## Methods

A systematic review was conducted on available literature from online databases and reported following the Preferred Reporting Items for Systematic Reviews and Meta-Analysis 2020 checklist. A comprehensive search strategy was developed. Only Observational and interventional studies, which were ‘ primary research studies’ were included in assessing epigenetic markers derived from fetal or maternal tissue in conditions of maternal stress during pregnancy and were analyzed. We retrieved 492 papers most relevant to the query, mostly from PubMed, Semantic search tools and Journals. Both authors rigorously studied and summarized the abstracts to exclude the studies that did not meet the outlined criteria, even though well planned comprehensive studies were found, which lacked key inclusion criteria. Each study was further analysed for specific type of stress, gestational age (temporal characteristics), tissue sample analysed. Forty studies were shortlisted after screening and selected for the review, based on the inclusion criteria:

Studies were screened by the two reviewers independently. Risk of bias was assessed using RoB 2.0 tool Data was analyzed and tabulated descriptively, using the data extraction tool designed for the study.

Missing data were not analysed.

Keywords: epigenetic, maternal stress, fetal, neurodevelopment, Search stategy

Search: **(((maternal stress) AND (epigenetic markers)) AND (fetal)) AND (neurodevelopment)**

(“maternally”[All Fields] OR “maternities”[All Fields] OR “maternity”[All Fields] OR “mothers”[MeSH Terms] OR “mothers”[All Fields] OR “maternal”[All Fields]) AND (”stress”[All Fields] OR “stressed”[All Fields] OR “stresses”[All Fields] OR “stressful”[All Fields] OR “stressfulness”[All Fields] OR “stressing”[All Fields]) AND ((”epigenetical”[All Fields] OR “epigenetically”[All Fields] OR “epigenomics”[MeSH Terms] OR “epigenomics”[All Fields] OR “epigenetic”[All Fields] OR “epigenetics”[All Fields]) AND (“marker”[All Fields] OR “markers”[All Fields])) AND (”fetale”[All Fields] OR “fetally”[All Fields] OR “fetals”[All Fields] OR “fetus”[MeSH Terms] OR “fetus”[All Fields] OR “fetal”[All Fields] OR “foetal”[All Fields]) AND “neurodevelopment”[All Fields]

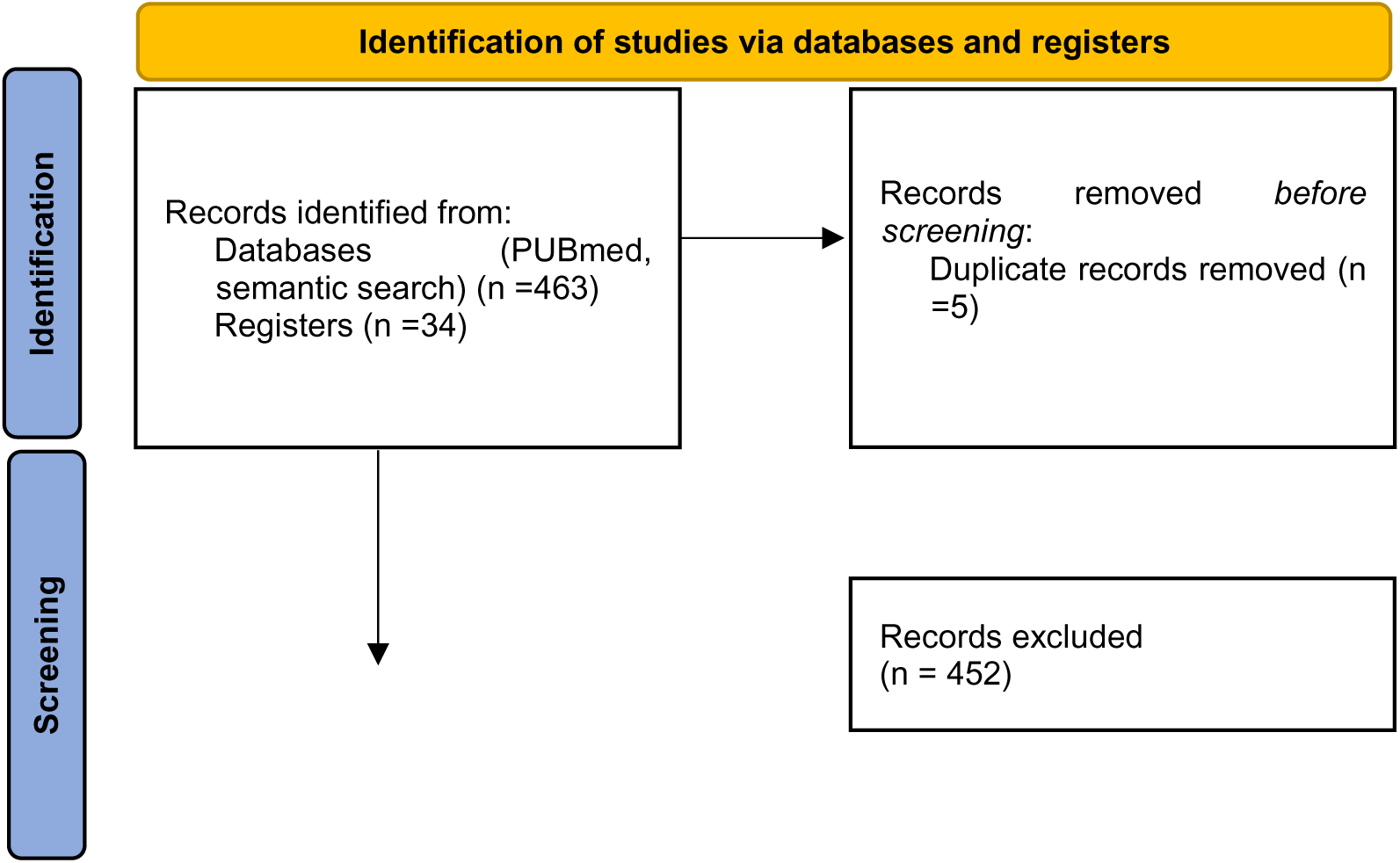
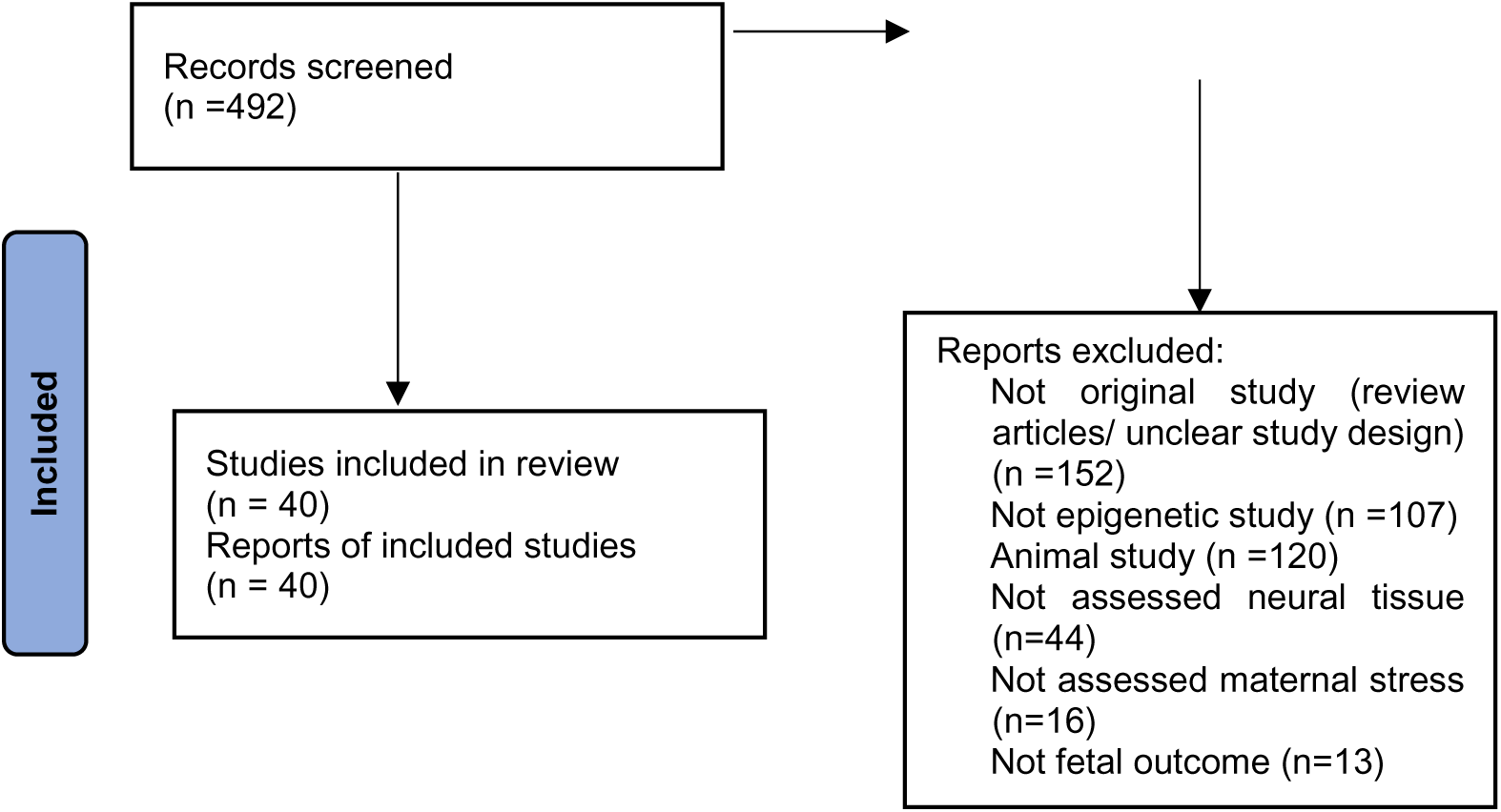

## Results

### Characteristics of included studies

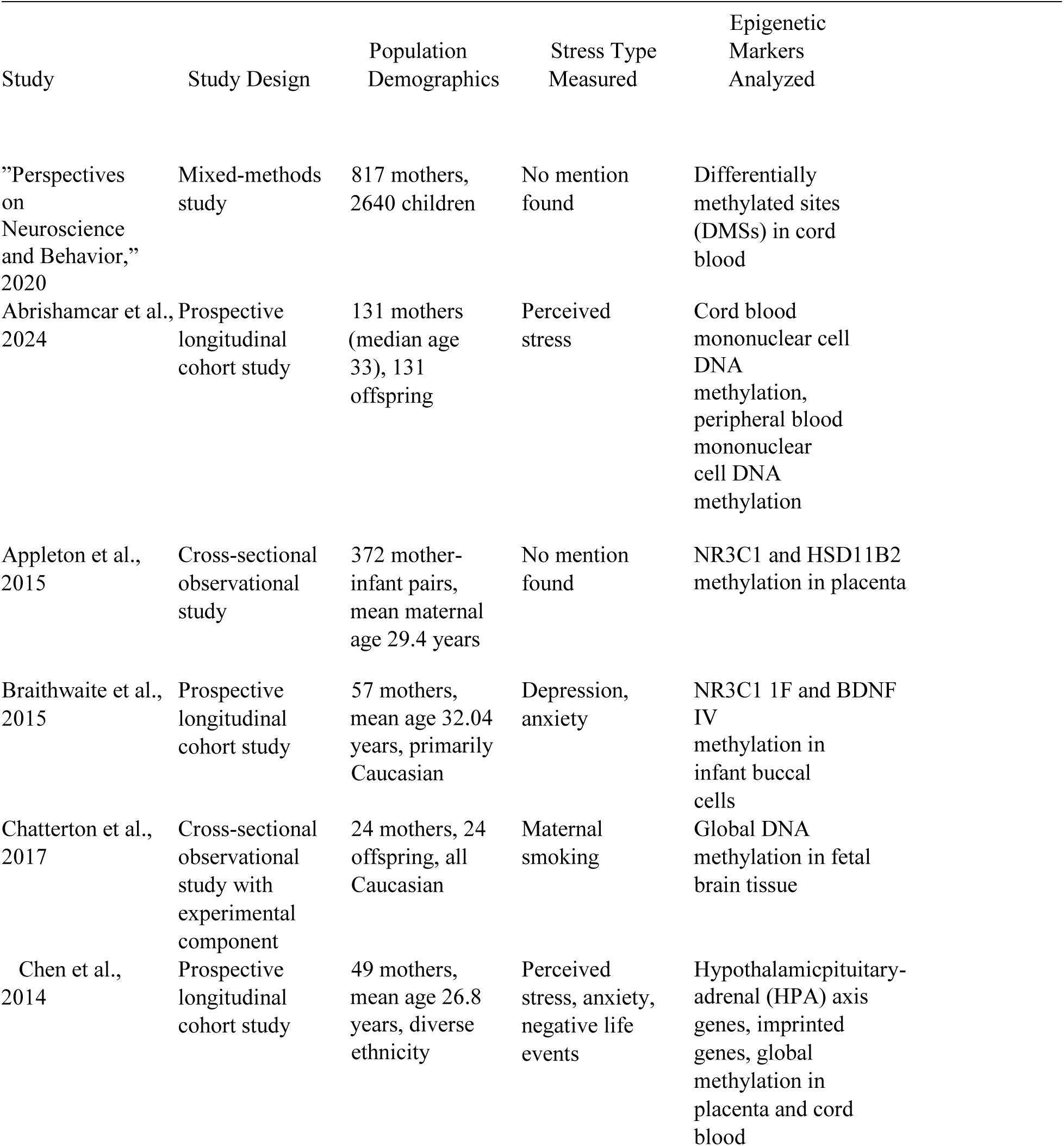
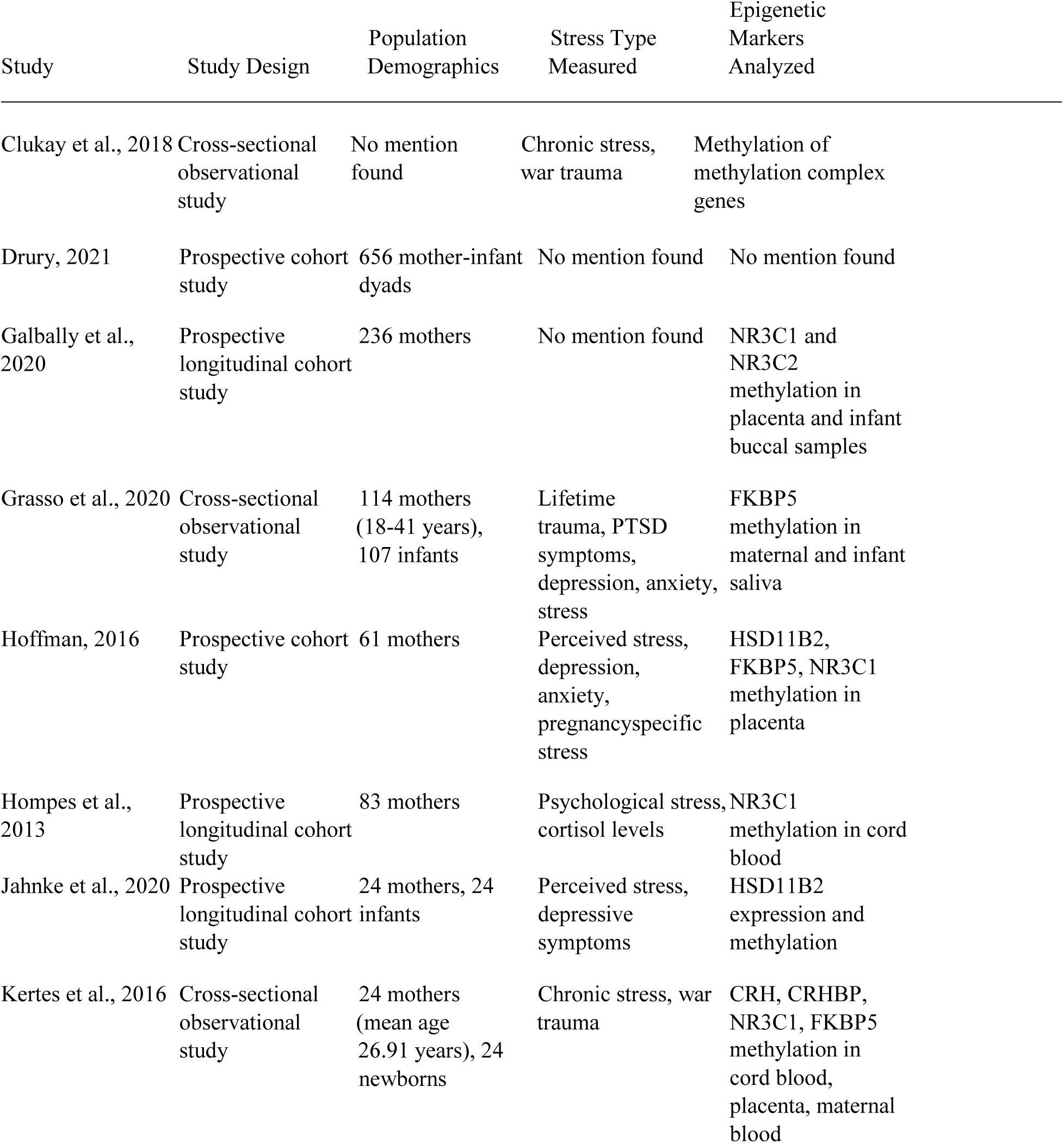
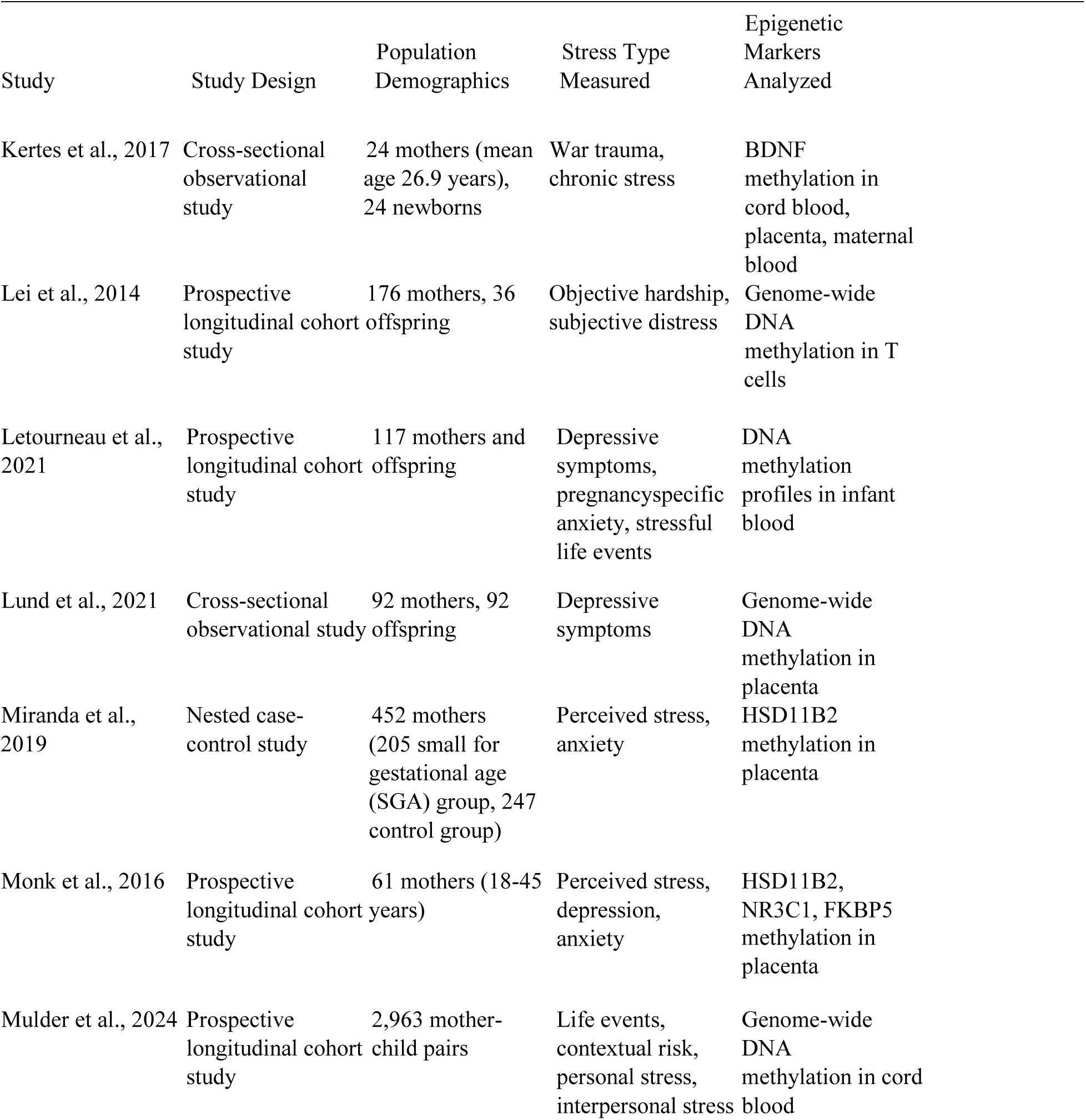
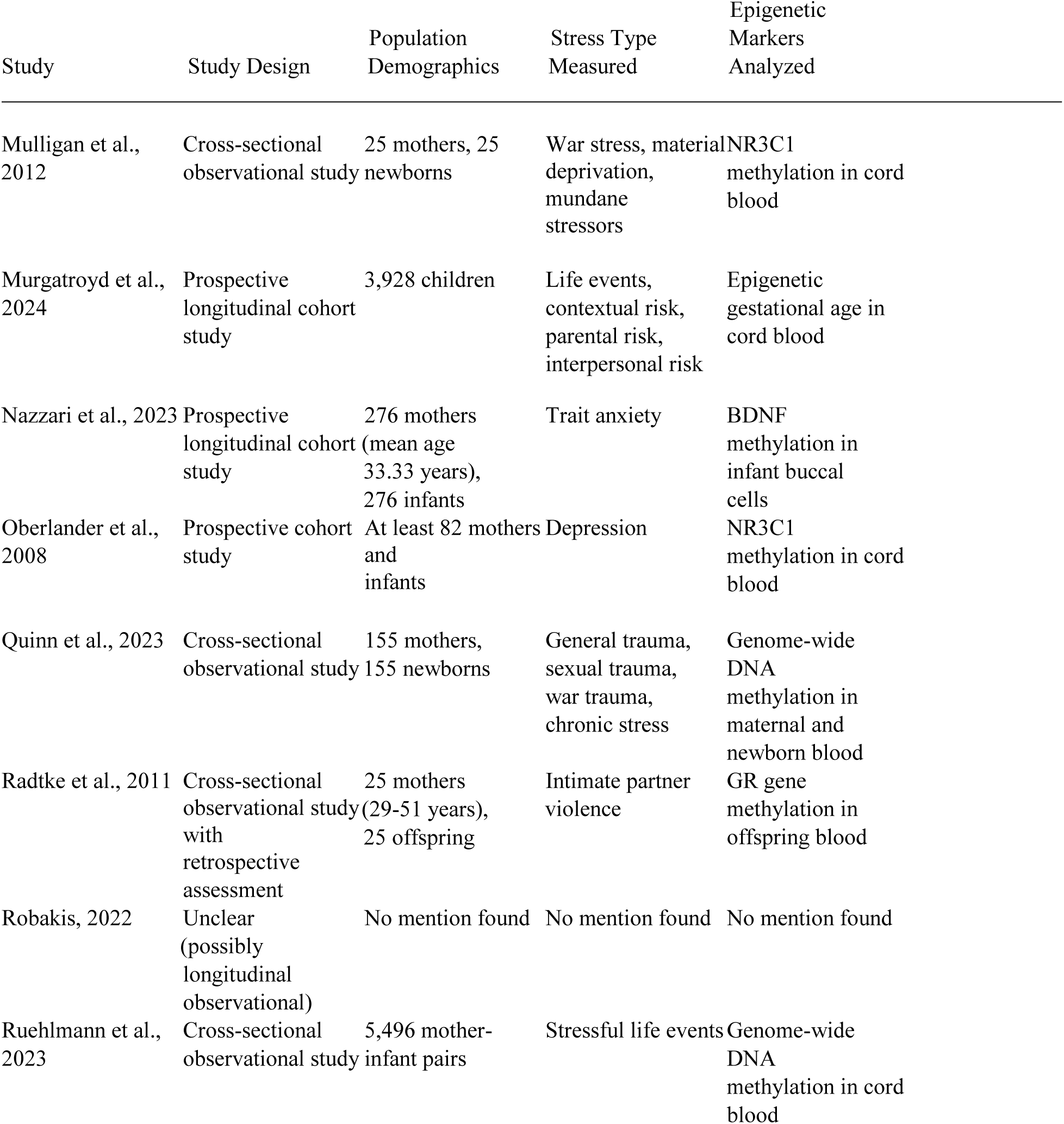
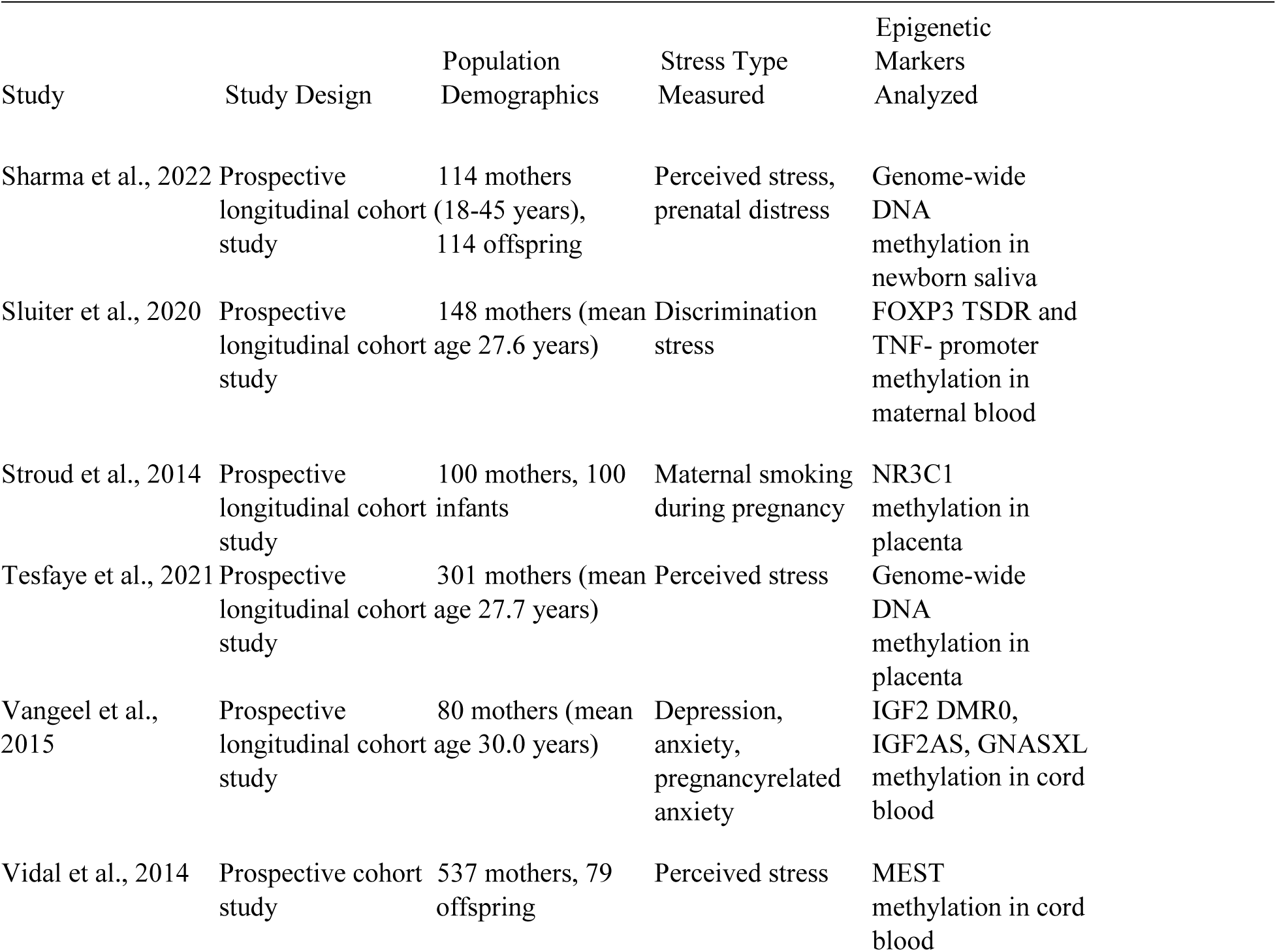
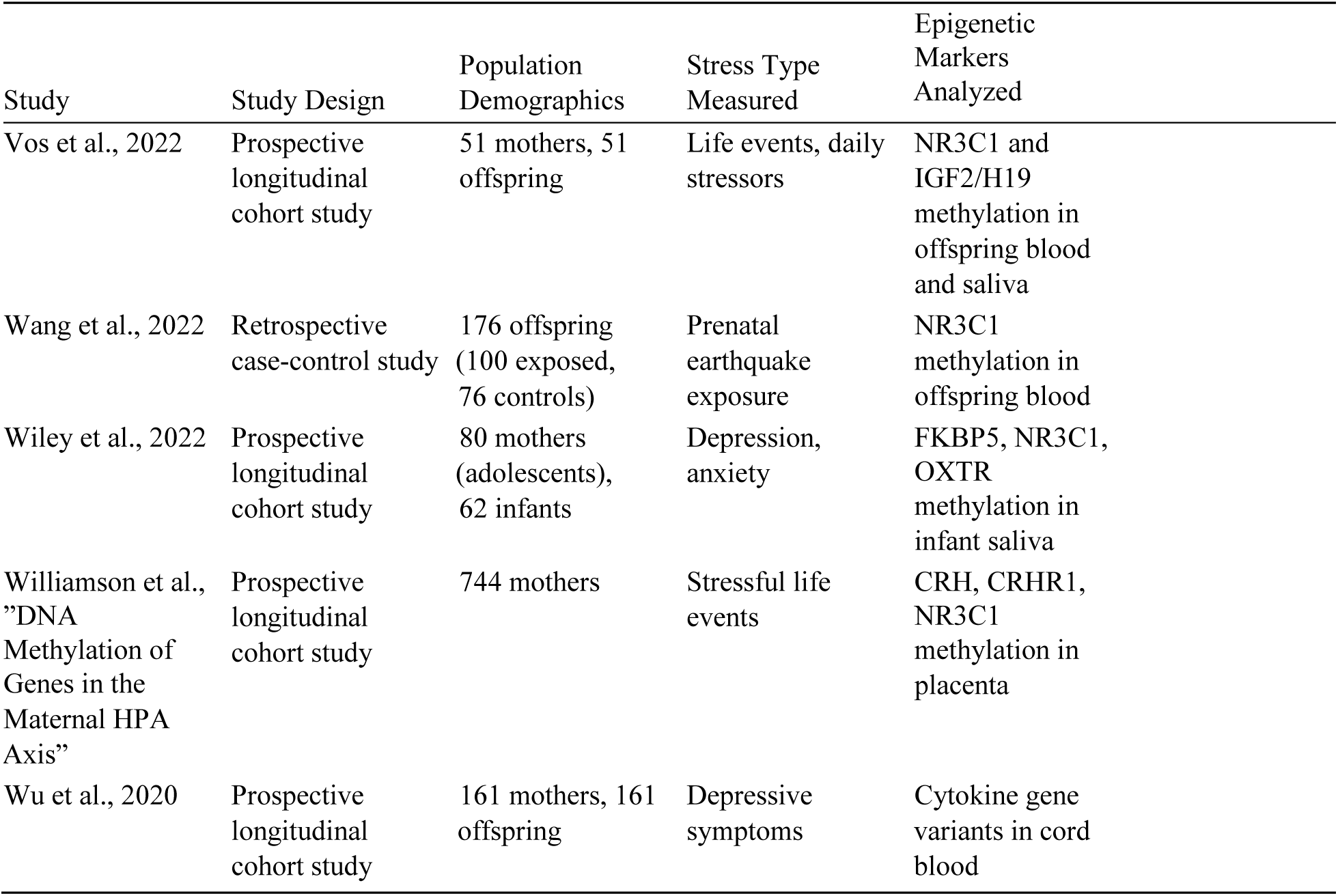

| Study | Study Design | Population Demographics | Stress Type Measured | Epigenetic Markers Analyzed |
| --- | --- | --- | --- | --- |
| "Perspectives on Neuroscience and Behavior," 2020 | Mixed-methods study | 817 mothers, 2640 children | No mention found | Differentially methylated sites (DMSs) in cord blood |
| Abrishamcar et al., 2024 | Prospective longitudinal cohort study | 131 mothers (median age 33), 131 offspring | Perceived stress | Cord blood mononuclear cell DNA methylation, peripheral blood mononuclear cell DNA methylation |
| Appleton et al., 2015 | Cross-sectional observational study | 372 mother-infant pairs, mean maternal age 29.4 years | No mention found | NR3C1 and HSD11B2 methylation in placenta |
| Braithwaite et al., 2015 | Prospective longitudinal cohort study | 57 mothers, mean age 32.04 years, primarily Caucasian | Depression, anxiety | NR3C1 1F and BDNF IV methylation in infant buccal cells |
| Chatterton et al., 2017 | Cross-sectional observational study with experimental component | 24 mothers, 24 offspring, all Caucasian | Maternal smoking | Global DNA methylation in fetal brain tissue |
| Chen et al., 2014 | Prospective longitudinal cohort study | 49 mothers, mean age 26.8 years, diverse ethnicity | Perceived stress, anxiety, negative life events | Hypothalamicpituitary-adrenal (HPA) axis genes, imprinted genes, global methylation in placenta and cord blood |
| Clukay et al., 2018 | Cross-sectional observational study | No mention found | Chronic stress, war trauma | Methylation of methylation complex genes |
| Drury, 2021 | Prospective cohort study | 656 mother-infant dyads | No mention found | No mention found |
| Galbally et al., 2020 | Prospective longitudinal cohort study | 236 mothers | No mention found | NR3C1 and NR3C2 methylation in placenta and infant buccal samples |
| Grasso et al., 2020 | Cross-sectional observational study | 114 mothers (18-41 years), 107 infants | Lifetime trauma, PTSD symptoms, depression, anxiety, stress | FKBP5 methylation in maternal and infant saliva |
| Hoffman, 2016 | Prospective cohort study | 61 mothers | Perceived stress, depression, anxiety, pregnancy-specific stress | HSD11B2, FKBP5, NR3C1 methylation in placenta |
| Hompes et al., 2013 | Prospective longitudinal cohort study | 83 mothers | Psychological stress, cortisol levels | NR3C1 methylation in cord blood |
| Jahnke et al., 2020 | Prospective longitudinal cohort study | 24 mothers, 24 infants | Perceived stress, depressive symptoms | HSD11B2 expression and methylation |
| Kertes et al., 2016 | Cross-sectional observational study | 24 mothers (mean age 26.91 years), 24 newborns | Chronic stress, war trauma | CRH, CRHBP, NR3C1, FKBP5 methylation in cord blood, placenta, maternal blood |

Characteristics of included studies
| Study | Study Design | Population Demographics | Stress Type Measured | Epigenetic Markers Analyzed |
| --- | --- | --- | --- | --- |
| Kertes et al., 2017 | Cross-sectional observational study | 24 mothers (mean age 26.9 years), 24 newborns | War trauma, chronic stress | BDNF methylation in cord blood, placenta, maternal blood |
| Lei et al., 2014 | Prospective longitudinal cohort study | 176 mothers, 36 offspring | Objective hardship, subjective distress | Genome-wide DNA methylation in T cells |
| Letourneau et al., 2021 | Prospective longitudinal cohort study | 117 mothers and offspring | Depressive symptoms, pregnancy-specific anxiety, stressful life events | DNA methylation profiles in infant blood |
| Lund et al., 2021 | Cross-sectional observational study | 92 mothers, 92 offspring | Depressive symptoms | Genome-wide DNA methylation in placenta |
| Miranda et al., 2019 | Nested case-control study | 452 mothers (205 small for gestational age (SGA) group, 247 control group) | Perceived stress, anxiety | HSD11B2 methylation in placenta |
| Monk et al., 2016 | Prospective longitudinal cohort study | 61 mothers (18-45 years) | Perceived stress, depression, anxiety | HSD11B2, NR3C1, FKBP5 methylation in placenta |
| Mulder et al., 2024 | Prospective longitudinal cohort study | 2,963 mother-child pairs | Life events, contextual risk, personal stress, interpersonal stress | Genome-wide DNA methylation in cord blood |
| Mulligan et al., 2012 | Cross-sectional observational study | 25 mothers, 25 newborns | War stress, material deprivation, mundane stressors | NR3C1 methylation in cord blood |
| Murgatroyd et al., 2024 | Prospective longitudinal cohort study | 3,928 children | Life events, contextual risk, parental risk, interpersonal risk | Epigenetic gestational age in cord blood |
| Nazzari et al., 2023 | Prospective longitudinal cohort study | 276 mothers (mean age 33.33 years), 276 infants | Trait anxiety | BDNF methylation in infant buccal cells |
| Oberlander et al., 2008 | Prospective cohort study | At least 82 mothers and infants | Depression | NR3C1 methylation in cord blood |
| Quinn et al., 2023 | Cross-sectional observational study | 155 mothers, 155 newborns | General trauma, sexual trauma, war trauma, chronic stress | Genome-wide DNA methylation in maternal and newborn blood |
| Radtke et al., 2011 | Cross-sectional observational study with retrospective assessment | 25 mothers (29-51 years), 25 offspring | Intimate partner violence | GR gene methylation in offspring blood |
| Robakis, 2022 | Unclear (possibly longitudinal observational) | No mention found | No mention found | No mention found |
| Ruehlmann et al., 2023 | Cross-sectional observational study | 5,496 mother-infant pairs | Stressful life events | Genome-wide DNA methylation in cord blood |

Characteristics of included studies
| Study | Study Design | Population Demographics | Stress Type Measured | Epigenetic Markers Analyzed |
| --- | --- | --- | --- | --- |
| Sharma et al., 2022 | Prospective longitudinal cohort study | 114 mothers (18-45 years), 114 offspring | Perceived stress, prenatal distress | Genome-wide DNA methylation in newborn saliva |
| Sluiter et al., 2020 | Prospective longitudinal cohort study | 148 mothers (mean age 27.6 years) | Discrimination stress | FOXP3 TSDR and TNF- promoter methylation in maternal blood |
| Stroud et al., 2014 | Prospective longitudinal cohort study | 100 mothers, 100 infants | Maternal smoking during pregnancy | NR3C1 methylation in placenta |
| Tesfaye et al., 2021 | Prospective longitudinal cohort study | 301 mothers (mean age 27.7 years) | Perceived stress | Genome-wide DNA methylation in placenta |
| Vangeel et al., 2015 | Prospective longitudinal cohort study | 80 mothers (mean age 30.0 years) | Depression, anxiety, pregnancy-related anxiety | IGF2 DMR0, IGF2AS, GNASXL methylation in cord blood |
| Vidal et al., 2014 | Prospective cohort study | 537 mothers, 79 offspring | Perceived stress | MEST methylation in cord blood |
| Vos et al., 2022 | Prospective longitudinal cohort study | 51 mothers, 51 offspring | Life events, daily stressors | NR3C1 and IGF2/H19 methylation in offspring blood and saliva |
| Wang et al., 2022 | Retrospective case-control study | 176 offspring (100 exposed, 76 controls) | Prenatal earthquake exposure | NR3C1 methylation in offspring blood |
| Wiley et al., 2022 | Prospective longitudinal cohort study | 80 mothers (adolescents), 62 infants | Depression, anxiety | FKBP5, NR3C1, OXTR methylation in infant saliva |
| Williamson et al., "DNA Methylation of Genes in the Maternal HPA Axis" | Prospective longitudinal cohort study | 744 mothers | Stressful life events | CRH, CRHR1, NR3C1 methylation in placenta |
| Wu et al., 2020 | Prospective longitudinal cohort study | 161 mothers, 161 offspring | Depressive symptoms | Cytokine gene variants in cord blood |

Our analysis of the information provided about 40 studies on prenatal stress and epigenetic markers revealed:

Study Design:

- 25 studies used a prospective longitudinal cohort design
- 11 studies were cross-sectional observational
- 4 studies used other designs (nested case-control, retrospective case-control, mixed-methods)

Population Demographics:

- 27 studies included both mothers and children
- 9 studies focused on mothers only
- 2 studies focused on children only
- We didn’t find mention of population information for 2 of the studies

Stress Types Measured:

- Depression was the most common (11 studies)
- Perceived stress and anxiety were each measured in 9 studies
- War trauma/stress and chronic stress were each measured in 4 studies
- We didn’t find mention of stress type information for 5 studies

Markers Analyzed:

- NR3C1 was the most commonly studied gene (14 studies)
- Cord blood (14 studies) and placenta (12 studies) were the most frequent tissue types
- 7 studies examined genome-wide methylation
- Other commonly studied markers included FKBP5 (5 studies) and HSD11B2 (5 studies)
- We didn’t find mention of epigenetic marker information for 2 studies

Many studies examined multiple stress types and epigenetic markers, reflecting the complexity of prenatal stress effects on epigenetic programming.

### Thematic analysis

#### Epigenetic Effects by Stress Type

##### Chronic Psychological Stress

| Study | Stress Measure | Primary Epigenetic Changes | Affected Genes | Gestational Period |
| --- | --- | --- | --- | --- |
| Chen et al., 2014 | Perceived Stress Scale | Hyper-methylation | IGF2/H19 ICR | No mention found |
| Clukay et al., 2018 | Chronic stress | Methylation changes | DNMT1, DNMT3A, TET3, MBD2 | No mention found |
| Kertes et al., 2016 | Chronic stress | Methylation changes | CRH, NR3C1, FKBP5 | Throughout pregnancy |
| Kertes et al., 2017 | Chronic stress | Methylation changes | BDNF | Throughout pregnancy |
| Lei et al., 2014 | Objective hardship | Lower methylation | SCG5 | Throughout pregnancy |
| Monk et al., 2016 | Perceived Stress Scale | Increased methylation | HSD11B2, FKBP5 | 24-27 weeks |
| Mulligan et al., 2012 | Material deprivation, mundane stressors | Increased methylation | NR3C1 | Throughout pregnancy |
| Quinn et al., 2023 | Chronic stress | No significant changes | No mention found | No mention found |
| Sluiter et al., 2020 | Discrimination stress | Higher methylation | FOXP3 TSDR, TNF- promoter | 24-32 weeks |
| Tesfaye et al., 2021 | Perceived Stress Scale | Lower methylation | ETV3, IER5 | Throughout pregnancy |
| Vidal et al., 2014 | Perceived Stress Scale | Elevated methylation | MEST | 12 weeks |

We found information on stress measures for 23 out of the 40 studies in the provided data. The most common measure was the Perceived Stress Scale (PSS), used in 9 studies, followed by chronic stress (4 studies), adverse life events (3 studies), and stressful life events (3 studies). Other measures were used in 1-2 studies each.

Epigenetic changes were reported in 17 studies. The most common finding was methylation changes (6 studies), followed by increased/elevated/higher methylation (4 studies). Two studies reported no significant changes, while others found various forms of methylation alterations.

We found information on affected genes for 19 studies. The most frequently affected gene was NR3C1, mentioned in 5 studies, followed by FKBP5 and HSD11B2 (2 studies each). Many other genes were mentioned in single studies.

Gestational period was specified in 17 studies. Seven studies examined stress throughout pregnancy, while six mentioned specific weeks. Other studies focused on particular trimesters or combinations of trimesters.

We didn’t find mention of stress measures for 17 studies, epigenetic changes for 23 studies, affected genes for 21 studies, and gestational period for 23 studies.

##### Traumatic/War-Related Stress

| Study | Stress Measure | Primary Epigenetic Changes | Affected Genes | Gestational Period |
| --- | --- | --- | --- | --- |
| Kertes et al., 2016 | War trauma | Methylation changes | CRH, CRHBP, NR3C1, FKBP5 | Throughout pregnancy |
| Kertes et al., 2017 | War trauma | Methylation changes | BDNF | Throughout pregnancy |
| Lei et al., 2014 | Ice storm exposure | Methylation changes | 1675 CpG sites across 957 genes | Throughout pregnancy |
| Mulligan et al., 2012 | War stress | Increased methylation | NR3C1 | Throughout pregnancy |
| Quinn et al., 2023 | War trauma, sexual trauma | Differential methylation | No mention found | No mention found |
| Wang et al., 2022 | Prenatal earthquake exposure | Increased methylation | NR3C1 | Throughout pregnancy |
| Clukay et al., 2018 | War trauma | Methylation changes | DNMT1, DNMT3A, TET3, MBD2 | No mention found |
| Grasso et al., 2020 | Lifetime trauma | Reduced methylation (mothers), Increased methylation (infants) | FKBP5 | Third trimester |
| Radtke et al., 2011 | Intimate partner violence | Increased methylation | GR gene (NR3C1) | During pregnancy |

Key insights from the data:

- Consistently Affected Genes:

NR3C1 (Glucocorticoid Receptor): All studies reporting on this gene found increased methylation associated with various types of traumatic stress.

FKBP5: Methylation changes reported in multiple studies, with one study (Grasso et al., 2020) finding opposite directions of change in mothers versus infants.

- Types of Traumatic Stress included:
- War trauma/stress (5 studies)
- Natural disasters ( storm, earthquake)
- Intimate partner violence
- Sexual trauma
- Lifetime trauma

**–** Wide ranging Epigenetic effects:
**–** Lei et al. (2014) reported methylation changes in 1675 CpG sites across 957 genes in response to ice storm exposure, suggesting broad epigenetic impacts of severe traumatic stress.

Tissue Specificity:

**–** Studies examined methylation in various tissues, including cord blood, placenta, maternal blood, and infant saliva.
**–** Grasso et al. (2020) found opposite directions of methylation changes in mothers (reduced) versus infants (increased) for the FKBP5 gene, highlighting the importance of considering tissue-specific effects.

#### Anxiety and Depression

| *Study* | *Stress Measure* | *Primary Epigenetic Changes* | *Affected Genes* | *Gestational Period* |
| --- | --- | --- | --- | --- |
| Braithwaite et al., 2015 | Depression, anxiety | Increased NR3C1 1F methylation (males), Decreased BDNF IV methylation | NR3C1 1F, BDNF IV | Second or third trimester |
| Galbally et al., 2020 | Depression | Lower methylation | NR3C2 | Early pregnancy |
| Hompes et al., 2013 | Anxiety | Methylation changes | NR3C1 | Each trimester |
| Lund et al., 2021 | Depressive symptoms | Methylation changes | CpG sites | weeks |
| Nazzari et al., 2023 | Trait anxiety | Increased methylation (males) | BDNF | Throughout pregnancy |
| Oberlander et al., 2008 | Depression | Increased methylation | NR3C1 | Third trimester |
| Vangeel et al., 2015 | Depression, anxiety, pregnancy-related anxiety | Decreased methylation, Increased methylation | IGF2 DMR0, IGF2AS, GNASXL | Each trimester |
| Wiley et al., 2022 | Depression, anxiety | Lower methylation | OXTR2 | Early and late pregnancy |
| Wu et al., 2020 | Depressive symptoms | No mention found (gene variants) | Cytokine genes | 26 weeks |

Key insights from the data:

- Consistently Affected Genes:

**–** NR3C1: Multiple studies report methylation changes, with some showing increased methylation (Braithwaite et al., 2015; Oberlander et al., 2008) and others showing more complex patterns.
**–** BDNF: Decreased methylation reported by Braithwaite et al. (2015) and increased methylation in males reported by Nazzari et al. (2023).
**–** NR3C2: Lower methylation associated with depression in early pregnancy (Galbally et al., 2020).
**–** Imprinted Genes: Vangeel et al. (2015) reported methylation changes in IGF2 DMR0, IGF2AS, and GNASXL.
- Timing of Stress Exposure:

Studies examined stress at various points during pregnancy:

Some focused on specific trimesters (e.g., Galbally et al., 2020 - early pregnancy; Oberlander et al., 2008 - third trimester).Others assessed stress throughout pregnancy (e.g., Nazzari et al., 2023). Vangeel et al. (2015) examined stress in each trimester, allowing for comparison across gestation.

- Wide-ranging Epigenetic Effects:

Lund et al. (2021) reported methylation changes in 2833 CpG sites associated with maternal depressive symptoms, suggesting broad epigenetic impacts of depression.

- Tissue Specificity:

Studies examined methylation in various tissues:

Cord blood (e.g., Oberlander et al., 2008; Vangeel et al., 2015. Placenta (e.g., Galbally et al., 2020). Infant buccal cells (e.g., Nazzari et al., 2023)

Functional Implications:

**–** Galbally et al. (2020) associated NR3C2 methylation changes with infant cortisol reactivity.
**–** Nazzari et al. (2023) linked BDNF methylation to infant negative emotionality.

### Tissue-Specific Methylation Patterns

#### Placental Tissue Findings

**Tissue-Specific Methylation Patterns Placental Tissue Findings**
| Study | Key Methylation Changes | Associated Stress Types | Developmental Impact |
| --- | --- | --- | --- |
| Appleton et al., 2015 | NR3C1 and HSD11B2 methylation changes | No mention found | Associated with infant neurobehavior |
| Chen et al., 2014 | Higher expression of HPA genes, lower expression of EGR1 and LUMA | Perceived stress, anxiety, negative life events | No mention found |
| Galbally et al., 2020 | Lower NR3C2 methylation at CpG_24 | Maternal depression | Associated with infant cortisol reactivity |
| Hoffman, 2016 | Increased HSD11B2 and FKBP5 methylation | Perceived stress | Associated with fetal coupling |
| Kertes et al., 2016 | Methylation changes in CRH, CRHBP, NR3C1, FKBP5 | Chronic stress, war trauma | Associated with birth weight |
| Kertes et al., 2017 | Methylation changes in BDNF | War trauma, chronic stress | No mention found |
| Lund et al., 2021 | Methylation changes in 2833 CpG sites | Maternal depressive symptoms | Enriched for neurodevelopmental functions |
| Miranda et al., 2019 | Decreased HSD11B2 methylation in SGA cases | Maternal stress | Associated with fetal growth restriction |
| Monk et al., 2016 | Increased HSD11B2 and FKBP5 methylation | Perceived stress | Associated with fetal behavior |
| Stroud et al., 2014 | Altered NR3C1 methylation | Maternal smoking during pregnancy | Associated with infant cortisol levels |
| Tesfaye et al., 2021 | Lower methylation of ETV3 and IER5 | Perceived stress | No mention found |
| Williamson et al., "DNA Methylation of Genes in the Maternal HPA Axis" | Methylation changes in CRH, CRHR1, NR3C1 | Stressful life events | Associated with gestational age and birth weight |

Key insights from the data:

- Consistently Affected Genes:

**–** HPA axis-related genes: NR3C1, NR3C2, CRH, CRHBP, and FKBP5 show consistent methylation changes across multiple studies.
**–** HSD11B2: Several studies report methylation changes in this gene, which is crucial for regulating fetal exposure to maternal cortisol.
**–** Neurodevelopmental genes: BDNF methylation changes are reported by Kertes et al. (2017).
- Types of Maternal Stress:

**–** A wide range of stress types are associated with placental methylation changes, including perceived stress, depression, anxiety, war trauma, and chronic stress.
**–** Stroud et al. (2014) specifically examined the impact of maternal smoking during pregnancy.
- Wide-ranging Epigenetic Effects:

**–** Lund et al. (2021) reported methylation changes in 2833 CpG sites associated with maternal depressive symptoms, suggesting broad epigenetic impacts on placental tissue.
- Functional Implications:

**–** Chen et al. (2014) found that maternal stress was associated with higher expression of HPA genes and lower expression of EGR1 in placental tissue, suggesting functional consequences of these epigenetic changes.
**–** The enrichment of neurodevelopmental functions in stress-associated methylation changes (Lund et al., 2021) highlights the potential long-term impacts on offspring development.
- Stress-Specific Effects:

**–** Different types of stress appear to have distinct methylation patterns. For example, war trauma and chronic stress were associated with methylation changes in multiple genes (Kertes et al., 2016, 2017), while perceived stress was often associated with changes in HSD11B2 and FKBP5 (Hoffman, 2016; Monk et al., 2016).

#### Cord blood markers

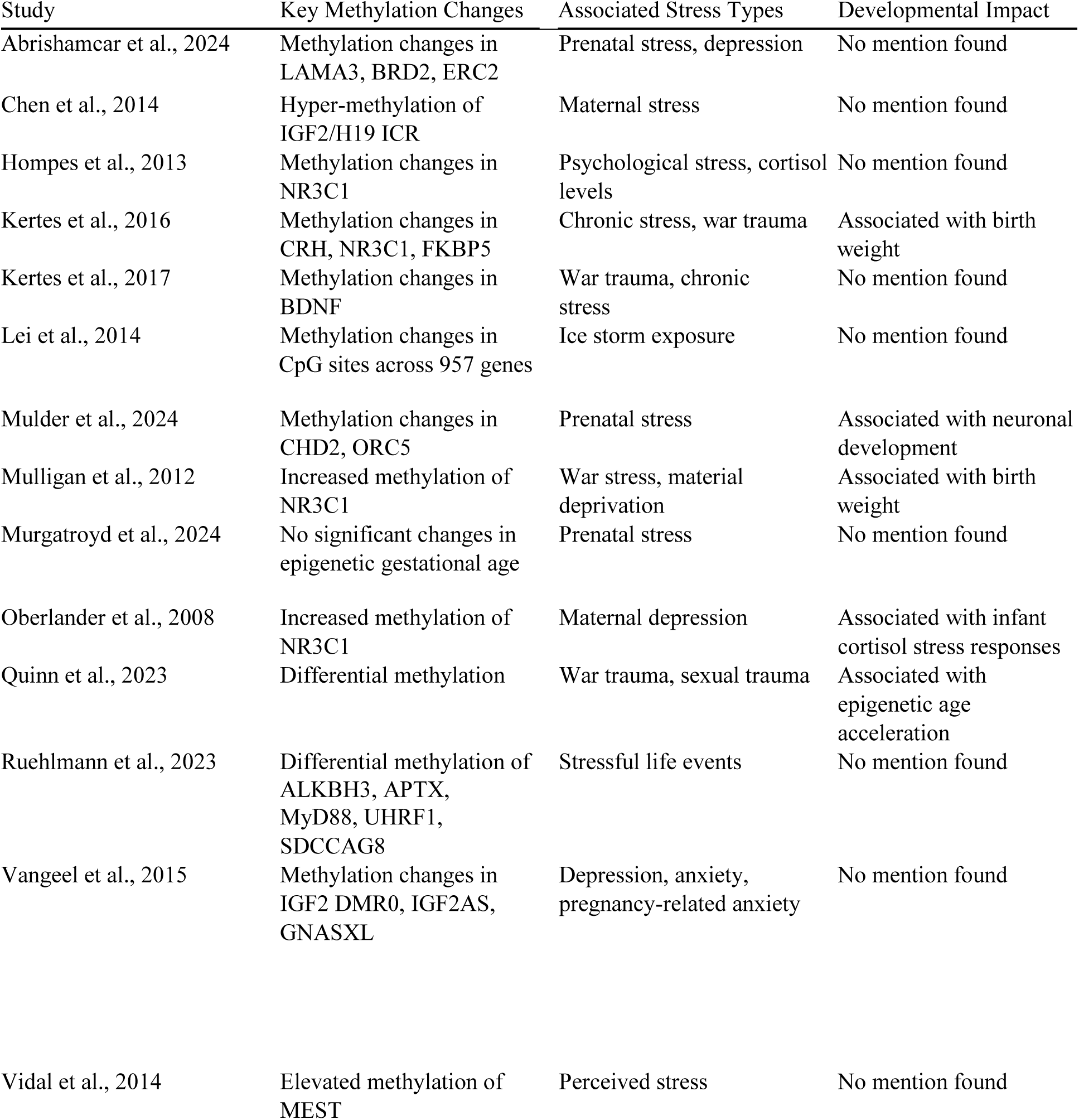

Key insights from the data:

- Consistently Affected Genes:

**–** NR3C1 (Glucocorticoid Receptor): Multiple studies report methylation changes in this gene, often showing increased methylation in response to various types of maternal stress (Hompes et al., 2013; Kertes et al., 2016; Mulligan et al., 2012; Oberlander et al., 2008).
**–** FKBP5: Methylation changes reported by Kertes et al. (2016).
**–** BDNF: Methylation changes associated with war trauma and chronic stress (Kertes et al., 2017).
**–** Imprinted Genes: IGF2/H19 ICR (Chen et al., 2014), IGF2 DMR0, IGF2AS, GNASXL (Vangeel et al., 2015), and MEST (Vidal et al., 2014) show methylation changes.
- Wide-ranging Epigenetic Effects:

**–** Lei et al. (2014) reported methylation changes in 1675 CpG sites across 957 genes in response to ice storm exposure, suggesting broad epigenetic impacts.
**–** Abrishamcar et al. (2024) and Ruehlmann et al. (2023) identified methylation changes in multiple genes, including some novel targets (e.g., LAMA3, BRD2, ALKBH3).
- Types of Maternal Stress:

**–** A wide range of stress types are associated with cord blood methylation changes, including:

∗ Psychological stress and cortisol levels (Hompes et al., 2013)
∗ War trauma and chronic stress (Kertes et al., 2016, 2017; Mulligan et al., 2012)
∗ Natural disasters (Lei et al., 2014)
∗ Depression and anxiety (Oberlander et al., 2008; Vangeel et al., 2015)
∗ Stressful life events (Ruehlmann et al., 2023)
- Developmental Impacts:

**–** Several studies link cord blood methylation changes to important developmental outcomes:

∗ Birth weight (Kertes et al., 2016; Mulligan et al., 2012)
∗ Infant cortisol stress responses (Oberlander et al., 2008)
∗ Epigenetic age acceleration (Quinn et al., 2023)
∗ Potential impacts on neuronal development (Mulder et al., 2024)
- Epigenetic Age:

**–** Quinn et al. (2023) found associations between maternal trauma exposure and epigenetic age acceleration in newborns.
**–** However, Murgatroyd et al. (2024) reported no significant changes in epigenetic gestational age associated with prenatal stress.

#### Maternal Blood Patterns

| Study | Key Methylation Changes | Associated Stress Types | Developmental Impact |
| --- | --- | --- | --- |
| Chen et al., 2014 | Hyper-methylation of IGF2/H19 ICR | Maternal stress | No mention found |
| Kertes et al., 2016 | Methylation changes in CRH, FKBP5 | Chronic stress, war trauma | No mention found |
| Kertes et al., 2017 | Methylation changes in BDNF | War trauma, chronic stress | No mention found |
| Quinn et al., 2023 | Differential methylation | General trauma, sexual trauma, war trauma | Associated with epigenetic age acceleration |
| Sluiter et al., 2020 | Higher methylation of FOXP3 TSDR and TNF- promoter | Discrimination stress | Associated with postnatal emotional distress |

Key insights from the data:

- Affected Genes:

**–** IGF2/H19 ICR: Chen et al. (2014) reported hyper-methylation associated with maternal stress.
**–** CRH and FKBP5: Kertes et al. (2016) found methylation changes related to chronic stress and war trauma.
**–** BDNF: Kertes et al. (2017) observed methylation changes associated with war trauma and chronic stress.
**–** FOXP3 TSDR and TNF- promoter: Sluiter et al. (2020) reported higher methylation related to discrimination stress.
- Types of Maternal Stress:

**–** A range of stress types are associated with maternal blood methylation changes, including:

∗ General maternal stress (Chen et al., 2014)
∗ Chronic stress and war trauma (Kertes et al., 2016, 2017)
∗ Discrimination stress (Sluiter et al., 2020) ∗

Various trauma types (Quinn et al., 2023)

- Developmental Impacts:

**–** Most studies did not directly link maternal blood methylation changes to offspring developmental outcomes.
Sluiter et al. (2020) associated maternal blood methylation changes with postnatal emotional distress.
**–** Quinn et al. (2023) found associations between maternal trauma exposure and epigenetic age acceleration.
- Epigenetic Age:

**–** Quinn et al. (2023) reported that sexual trauma was positively associated with epigenetic age acceleration in mothers.
- Immune System Involvement:

**–** Sluiter et al. (2020) focused on genes related to immune function (FOXP3 TSDR and TNF- promoter), suggesting a potential link between maternal stress, immune regulation, and epigenetic changes.
- Comparison with Other Tissues:

**–** Some studies (e.g., Chen et al., 2014; Kertes et al., 2016, 2017) examined methylation patterns in maternal blood alongside other tissues like placenta and cord blood, allowing for comparison across tissue types.

### Temporal Patterns and Critical Periods

#### Early Gestational Effects

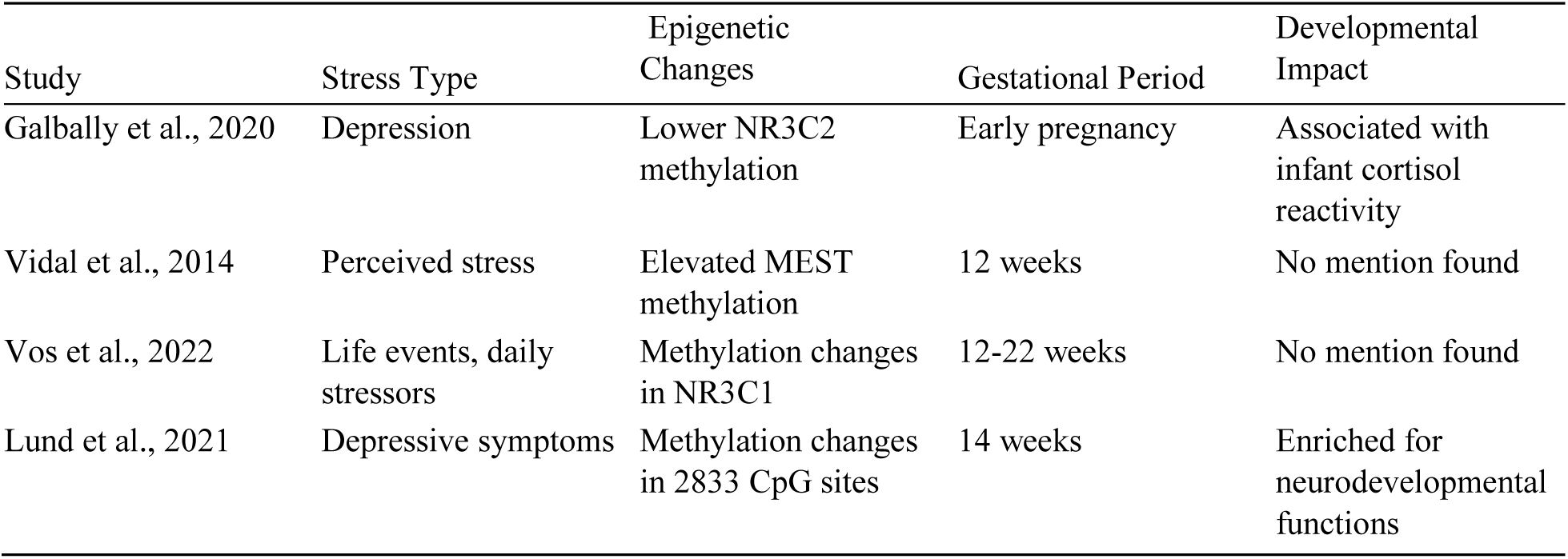

Key insights from the data:

NR3C2: Galbally et al. (2020) reported lower methylation associated with maternal depression in early pregnancy.
**–** MEST: Vidal et al. (2014) found elevated methylation related to perceived stress at 12 weeks gestation.
**–** NR3C1: Vos et al. (2022) observed methylation changes associated with life events and daily stressors between 12-22 weeks gestation.
**–** Genome-wide effects: Lund et al. (2021) reported methylation changes in 2833 CpG sites associated with maternal depressive symptoms at 14 weeks gestation.

- Types of Maternal Stress:

**–** The studies examined various forms of early gestational stress, including:

∗ Depression (Galbally et al., 2020)
∗ Perceived stress (Vidal et al., 2014)
∗ Life events and daily stressors (Vos et al., 2022)
∗ Depressive symptoms (Lund et al., 2021)

**–**

- Developmental Impacts:

**–** Galbally et al. (2020) linked early pregnancy stress-related epigenetic changes to infant cortisol reactivity.
**–** Lund et al. (2021) found that genes affected by early gestational stress were enriched for neurodevelopmental functions.
- Tissue Specificity:

**–** The studies examined epigenetic changes in various tissues:

∗ Placenta (Galbally et al., 2020; Lund et al., 2021)
∗ Cord blood (Vidal et al., 2014)
∗ Offspring blood and saliva (Vos et al., 2022)

| Study | Stress Type | Epigenetic Changes | Late Gestational Period | Developmental Impact |
| --- | --- | --- | --- | --- |
| Braithwaite et al., 2015 | Depression, anxiety | Increased NR3C1 1F methylation (males), Decreased BDNF IV methylation | Second or third trimester | No mention found |
| Grasso et al., 2020 | Lifetime trauma, PTSD symptoms | Reduced FKBP5 methylation (mothers), Increased FKBP5 methylation (infants) | Third trimester | No mention found |
| Hoffman, 2016 | Perceived stress, depression, anxiety | Increased HSD11B2 and FKBP5 methylation | 24-27 weeks | Associated with fetal coupling |
| Monk et al., 2016 | Perceived stress, depression, anxiety | Increased HSD11B2 and FKBP5 methylation | 24-27 weeks | Associated with fetal behavior |
| Oberlander et al., 2008 | Depression | Increased NR3C1 methylation | Third trimester | Associated with infant cortisol stress responses |
| Sharma et al., 2022 | Perceived stress, prenatal distress | Hypermethylation of YAP1 | Third trimester | No mention found |
| Sluiter et al., 2020 | Discrimination stress | Higher FOXP3 TSDR and TNF- promoter methylation | 24-32 weeks | Associated with postnatal emotional distress |
| Tesfaye et al., 2021 | Perceived stress | Lower methylation of ETV3 and IER5 | Third trimester | No mention found |

Key insights from the data:

- Consistently Affected Genes:

**–** NR3C1: Increased methylation reported in multiple studies (Braithwaite et al., 2015; Oberlander et al., 2008)
**–** FKBP5: Increased methylation in infants, but reduced in mothers (Grasso et al., 2020; Hoffman, 2016; Monk et al., 2016)
**–** HSD11B2: Increased methylation (Hoffman, 2016; Monk et al., 2016)
**–** BDNF: Decreased methylation (Braithwaite et al., 2015)
**–** Other genes: YAP1 (Sharma et al., 2022), FOXP3 TSDR and TNF- promoter (Sluiter et al., 2020), ETV3 and IER5 (Tesfaye et al., 2021)
- Types of Maternal Stress:

**–** Various forms of late gestational stress were examined, including:

∗ Depression and anxiety (Braithwaite et al., 2015; Hoffman, 2016; Monk et al., 2016; Oberlander et al., 2008)
∗ Trauma and PTSD symptoms (Grasso et al., 2020)
∗ Perceived stress and prenatal distress (Hoffman, 2016; Monk et al., 2016; Sharma et al., 2022; Tesfaye et al., 2021)
∗ Discrimination stress (Sluiter et al., 2020)
- Developmental Impacts:

**–** Fetal behavior and coupling (Hoffman, 2016; Monk et al., 2016)
**–** Infant cortisol stress responses (Oberlander et al., 2008)
**–** Postnatal emotional distress (Sluiter et al., 2020)
- Tissue Specificity:

**–** Studies examined epigenetic changes in various tissues:

∗ Cord blood (Oberlander et al., 2008)
∗ Placenta (Hoffman, 2016; Monk et al., 2016; Tesfaye et al., 2021)
∗ Infant buccal cells (Braithwaite et al., 2015)
∗ Maternal and infant saliva (Grasso et al., 2020)
∗ Newborn saliva (Sharma et al., 2022)
∗ Maternal blood (Sluiter et al., 2020)
- Sex-Specific Effects:

**–** Braithwaite et al. (2015) reported increased NR3C1 1F methylation only in male infants.
**–** The consistent involvement of stress-related genes (NR3C1, FKBP5, HSD11B2) suggests that late gestational stress may have significant impacts on the development of the fetal stress response system.
**–** Changes in HSD11B2 methylation may alter placental barrier function, potentially exposing the fetus to higher levels of maternal cortisol.

### Persistent Changes

| Study | Stress Type | Epigenetic Changes | Follow-up Period | Developmental Impact |
| --- | --- | --- | --- | --- |
| Radtke et al., 2011 | Intimate partner violence | Increased GR gene (NR3C1) methylation | 10-19 years after birth | No mention found |
| Vos et al., 2022 | Life events, daily stressors | Methylation changes in NR3C1 and IGF2/H19 | 28-29 years after birth | No mention found |
| Wang et al., 2022 | Prenatal earthquake exposure | Increased NR3C1 methylation | Adulthood | Associated with lower visuospatial memory performance |

Key insights from the data:

- Consistently Affected Genes:

**–** NR3C1 (Glucocorticoid Receptor): All three studies reporting persistent changes found increased methylation of the NR3C1 gene (Radtke et al., 2011; Vos et al., 2022; Wang et al., 2022). This consistency suggests that alterations in the stress response system may be a key mechanism for the long-term effects of prenatal stress.
- Types of Prenatal Stress:

**–** The studies examined various forms of prenatal stress: ∗ Intimate partner violence (Radtke et al., 2011)
∗ Life events and daily stressors (Vos et al., 2022)
∗ Earthquake exposure (Wang et al., 2022)
- Follow-up Periods:

**–** The studies demonstrate remarkably long-lasting effects, with follow-up periods ranging from 10-19 years (Radtke et al., 2011) to 28-29 years (Vos et al., 2022) after birth.
- Tissue Specificity:

**–** The studies examined epigenetic changes in different tissues:

∗ Peripheral blood (Radtke et al., 2011)
∗ Blood and saliva (Vos et al., 2022)
∗ Blood (Wang et al., 2022)
- Additional Genes:

**–** Vos et al. (2022) also reported persistent methylation changes in the IGF2/H19 imprinted region, suggesting that prenatal stress may have long-lasting effects on growth-related genes as well.
- Potential Mechanisms:

**–** The persistent methylation changes in NR3C1 suggest that prenatal stress may lead to long-term alterations in the regulation of the hypothalamic-pituitary-adrenal (HPA) axis.
**–** The involvement of IGF2/H19 (Vos et al., 2022) indicates that prenatal stress may also have lasting impacts on growth and metabolic regulation.

Summary of affected genes:

- Gene categories affected:

**–** Stress response genes: NR3C1, FKBP5, CRH, HSD11B2
**–** Neurodevelopmental genes: BDNF
**–** Imprinted genes: IGF2/H19, MEST
**–** Epigenetic regulators: DNMT1, DNMT3A, TET3
**–** Immune-related genes: FOXP3, TNF-
- Direction of Methylation Changes:

**–** Hypermethylation: IGF2/H19, HSD11B2, FKBP5
**–** Hypomethylation: SCG5, ETV3, IER5
**–** The variability suggests gene-specific epigenetic responses to chronic stress

## DISCUSSION

Most studies examining traumatic stress considered exposure throughout pregnancy, most likely due to the persistent nature of the stressors studied (e.g., war, natural disasters). The consistent involvement of stress-related genes (NR3C1, FKBP5) suggests that traumatic stress may impact the development of the fetal stress response system. Clukay et al. (2018) found methylation changes in genes involved in the methylation/demethylation complex (DNMT1, DNMT3A, TET3, MBD2), suggesting that traumatic stress may alter the epigenetic machinery itself.

Long-term Effects; Radtke et al. (2011) demonstrated that maternal exposure to intimate partner violence during pregnancy was associated with increased NR3C1 methylation in adolescent offspring, suggesting long-lasting epigenetic effects of prenatal trauma exposure. Maternal anxiety and depression was found to have Sex-Specific Effects. Braithwaite et al. (2015) found increased NR3C1 1F methylation only in male infants. Nazzari et al. (2023) reported increased BDNF methylation in males associated with maternal trait anxiety

Wiley et al. (2022) reported lower methylation of OXTR2 (oxytocin receptor) associated with maternal anxiety, suggesting a potential role for the oxytocin system in the ‘intergenerational transmission’ of stress effects. Whereas Wu et al. (2020) examined cytokine gene variants rather than methylation, highlighting the potential role of inflammatory pathways in mediating the effects of maternal depression on fetal development

The consistent involvement of HPA axis-related genes suggests that alterations in stress response systems may be a key mechanism by which maternal stress impacts fetal development.

Changes in HSD11B2 methylation may alter the placental barrier function, potentially exposing the fetus to higher levels of maternal cortisol.

The consistent involvement of HPA axis-related genes (NR3C1, FKBP5) suggests that alterations in stress response systems may be a key mechanism by which maternal stress impacts fetal development.

Methylation changes in imprinted genes (e.g., IGF2/H19) may affect fetal growth and development.

The involvement of stress-related genes (CRH, FKBP5) and growth-related genes (IGF2/H19) suggests that maternal stress may impact both stress response systems and growth regulation.

The findings related to immune-related genes (FOXP3, TNF-) highlight the potential role of immune system modulation in the maternal stress response.

Involvement of stress-related genes (NR3C1, NR3C2) suggests that early gestational stress may impact the development of the fetal stress response system. The wide-ranging effects observed by Lund et al. (2021) indicate that early gestational stress may have broad impacts on fetal epigenetic programming. While most studies did not examine long-term outcomes, the association with infant cortisol reactivity (Galbally et al., 2020) suggests that early gestational stress may have lasting effects on offspring stress physiology.

Several studies link placental methylation changes to important developmental outcomes. Infant neurobehavior (Appleton et al., 2015) Fetal coupling and behavior (Hoffman, 2016; Monk et al., 2016) Birth weight (Kertes et al., 2016; Williamson et al.) Fetal growth restriction (Miranda et al., 2019) Infant cortisol reactivity (Galbally et al., 2020; Stroud et al., 2014)

Wang et al. (2022) found that persistent NR3C1 methylation changes were associated with lower visuospatial memory performance in adulthood.

The other studies did not specify developmental impacts, highlighting a need for more research on the functional consequences of these persistent epigenetic changes.

## Conclusion

- Gestational timing: Studies assessed stress at various points during pregnancy, from specific trimesters to through-out gestation. This variation allows exploration of potential critical periods but complicates direct comparisons between studies.
- Inconsistent findings: Not all studies reported significant associations between chronic stress and epigenetic changes (e.g., Quinn et al., 2023), highlighting the complex nature of stress-epigenetic relationships.
- Methodological diversity: Studies employed various methods for epigenetic analysis, including candidate gene approaches and genome-wide analyses. This diversity contributes to the richness of the data but also makes direct comparisons challenging.
- Tissue Specificity: Epigenetic changes were assessed in various tissues, including placenta, cord blood, and maternal blood. The tissue-specific nature of epigenetic modifications should be considered when interpreting results across studies.

## Supporting information

https://docs.google.com/spreadsheets/d/15GPPPcWGwn1SKmsnFtXvmRt-dCPO3QH5/edit?usp=drive_link&ouid=115264355068055278464&rtpof=true&sd=true

## Data Availability

All data produced in the present study are available upon reasonable request to the authors

---

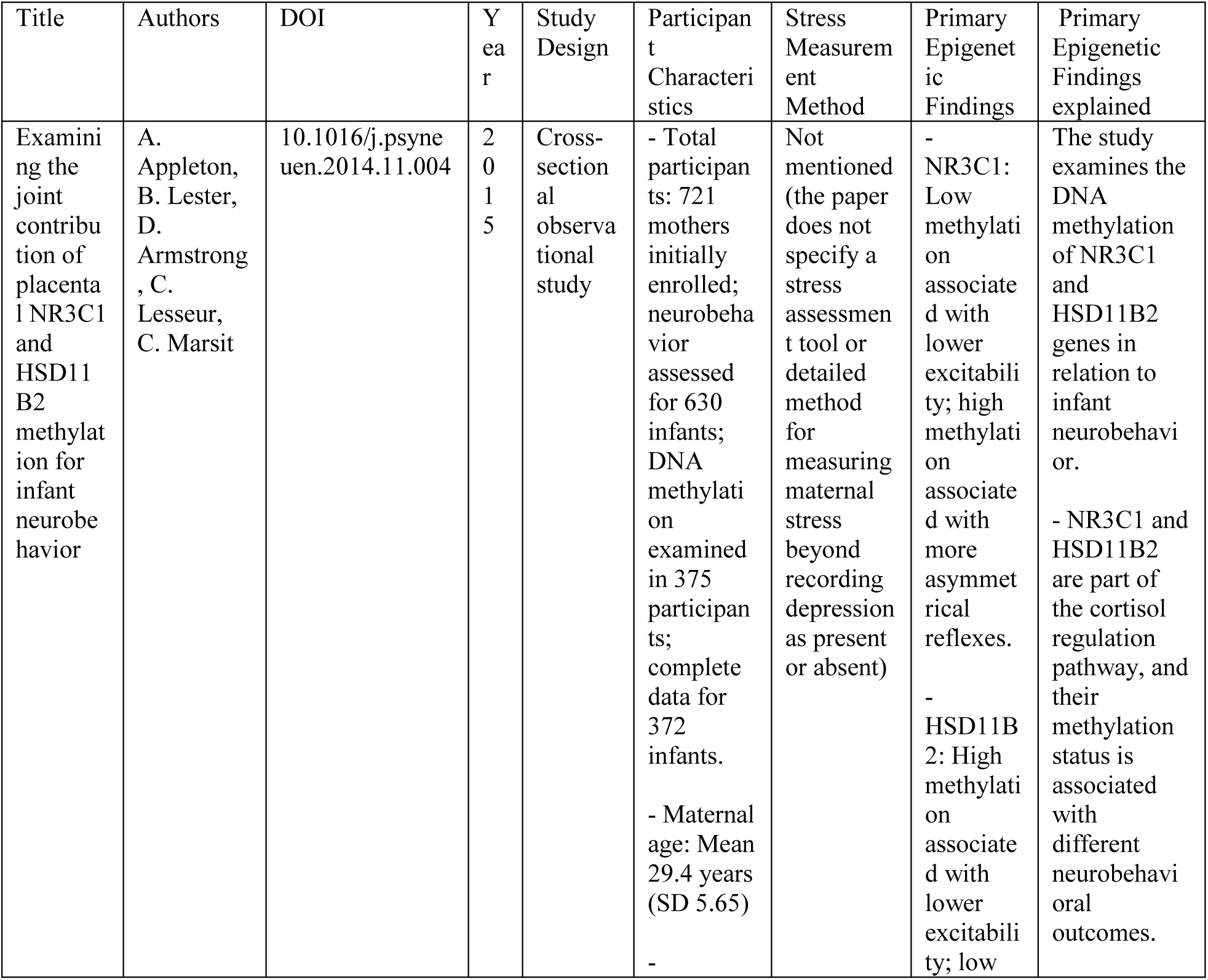
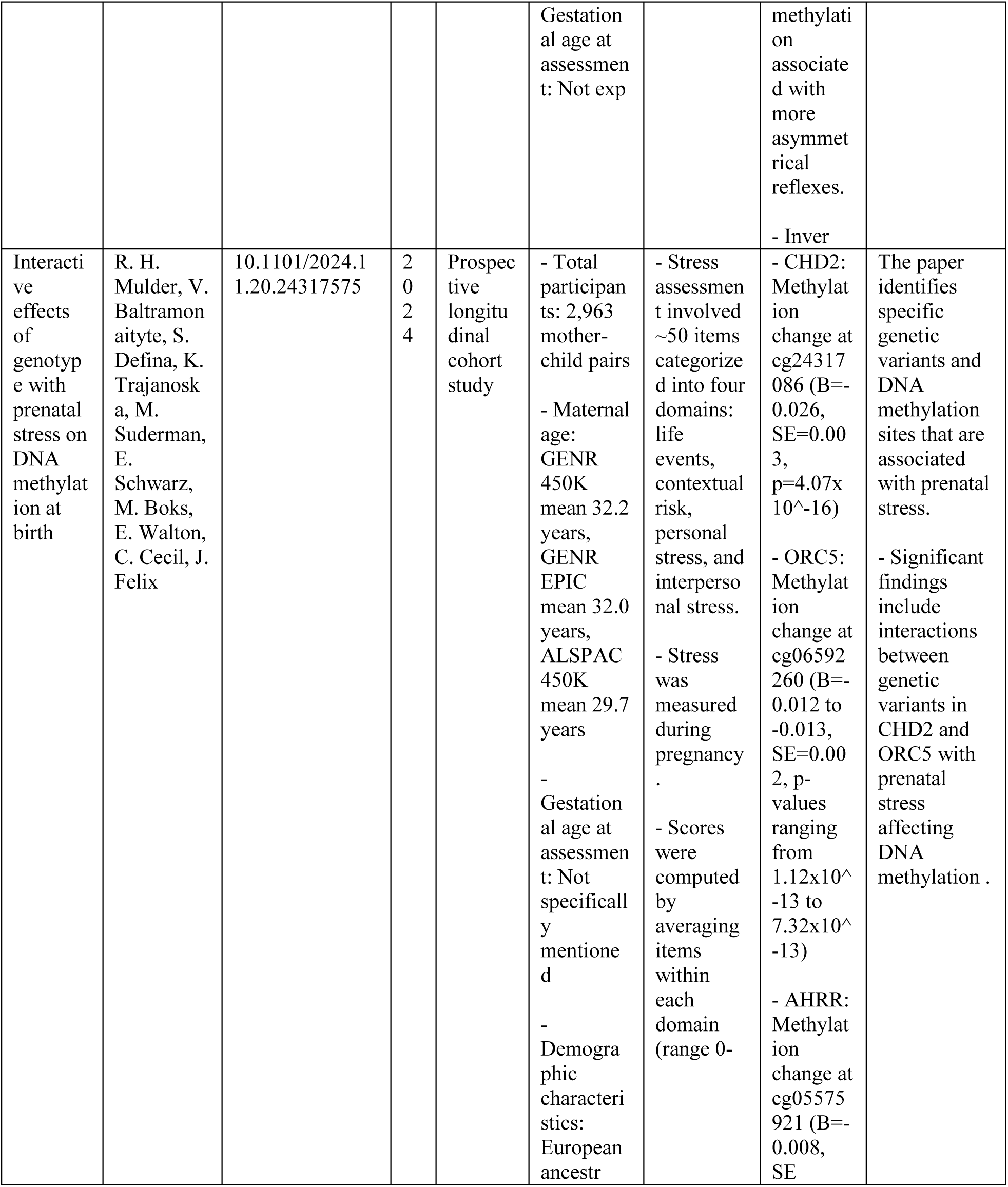
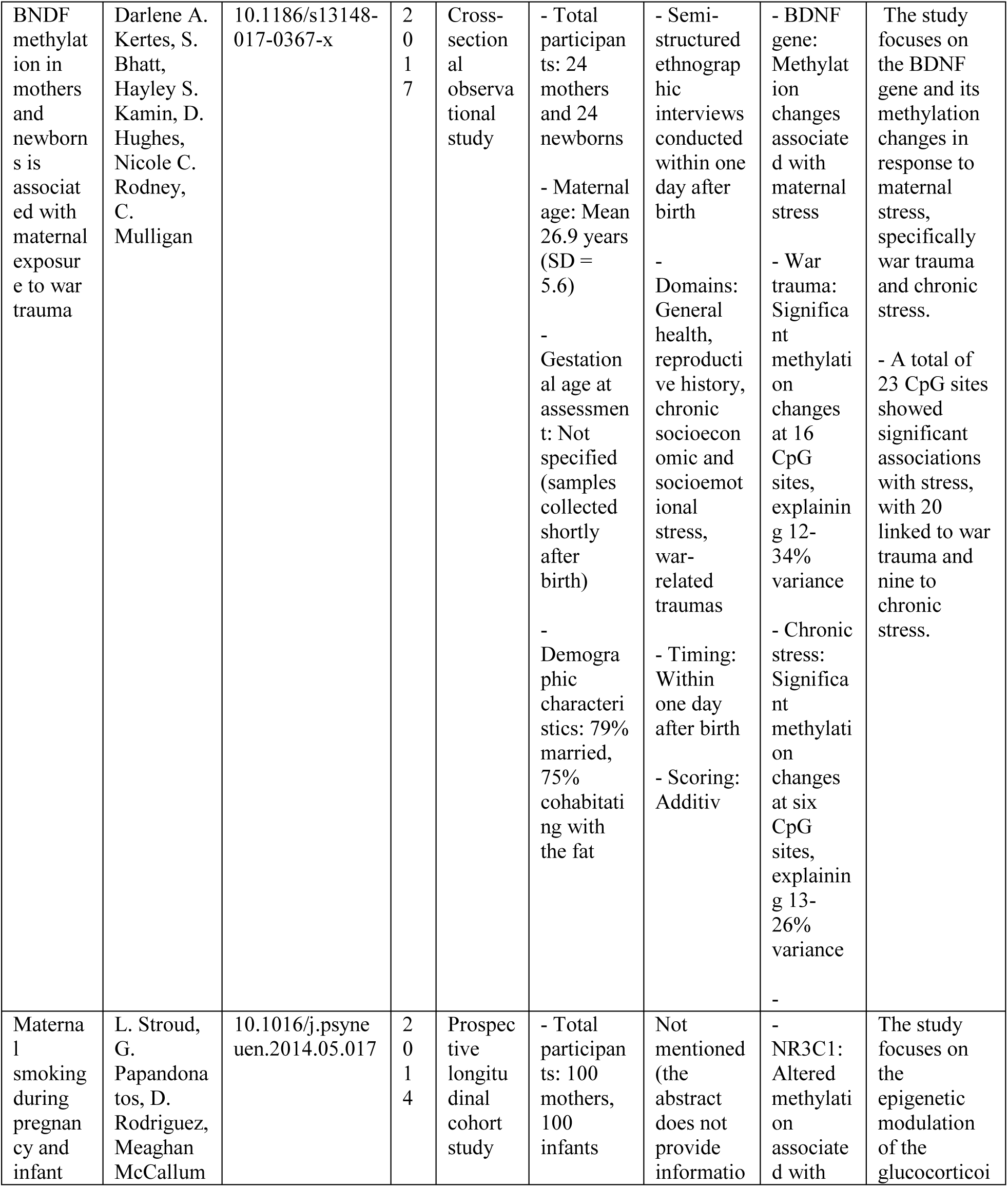
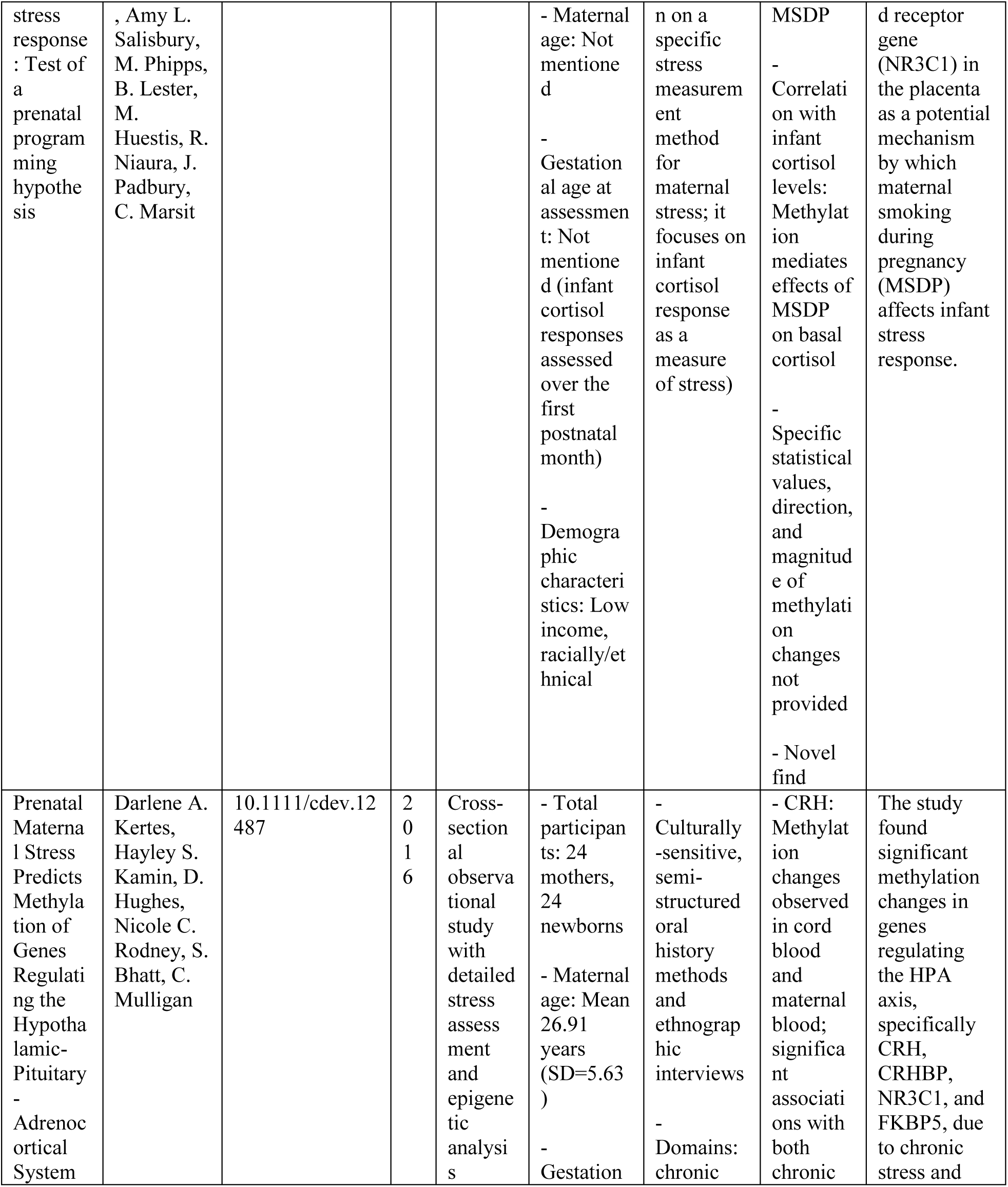
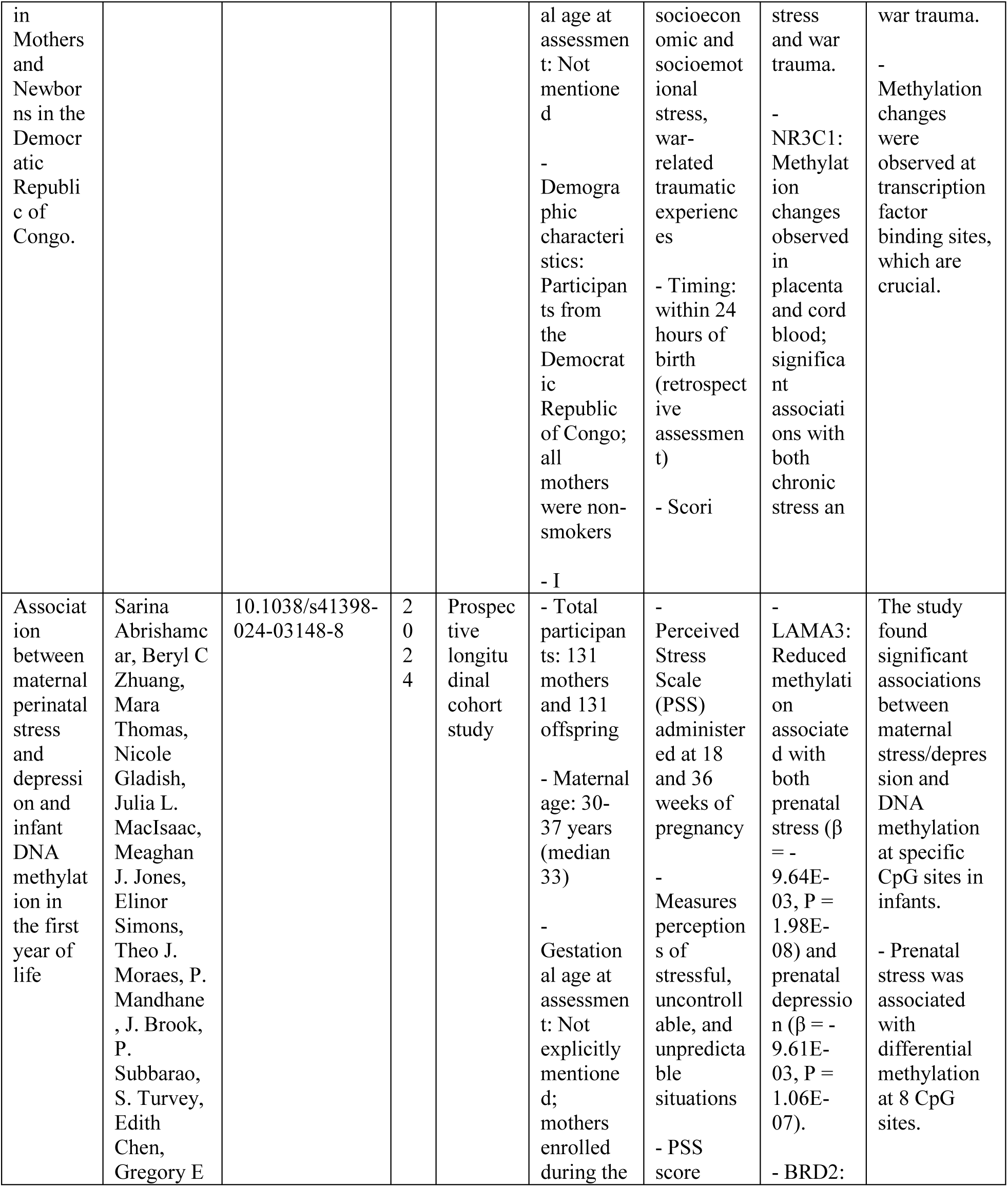
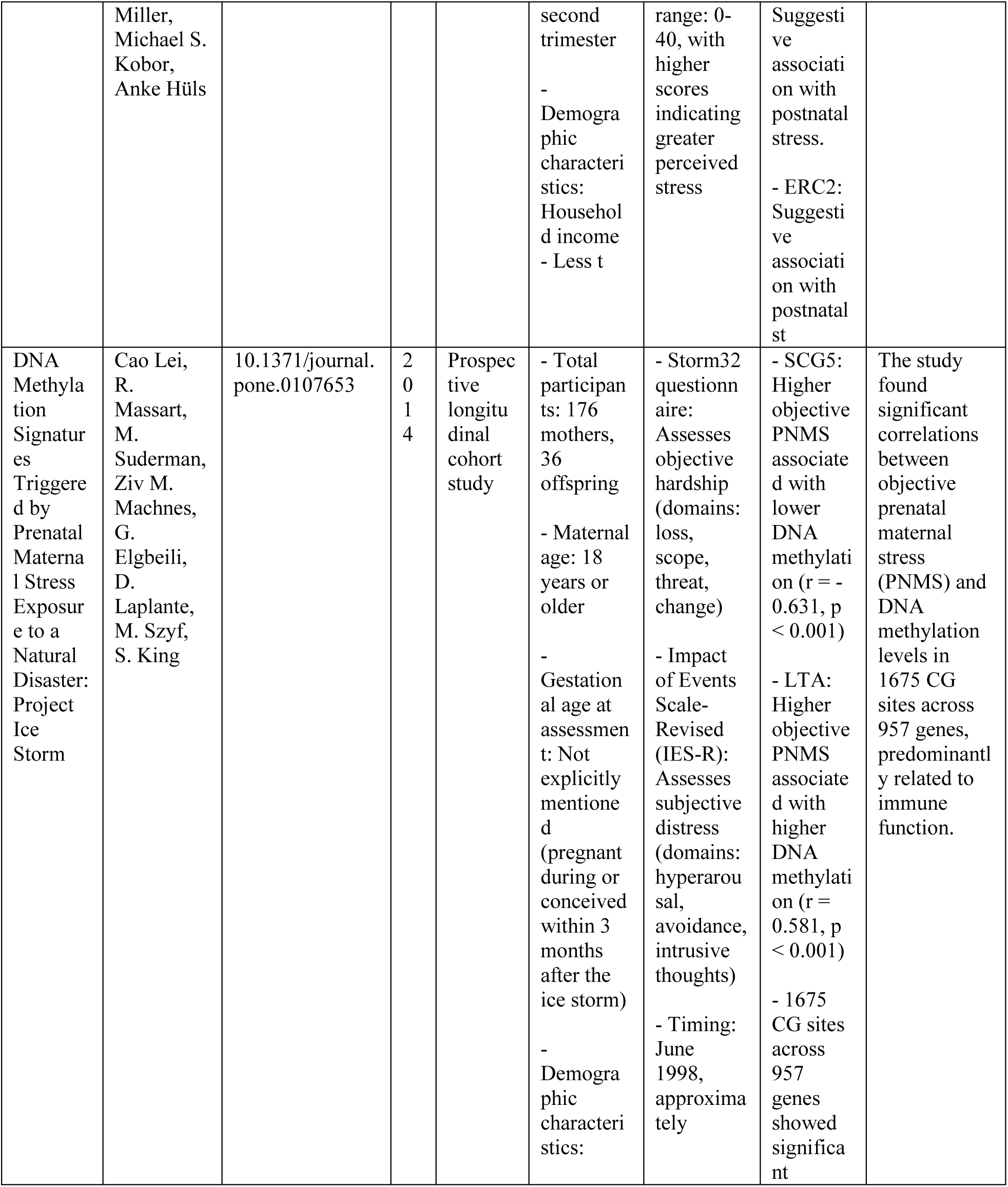
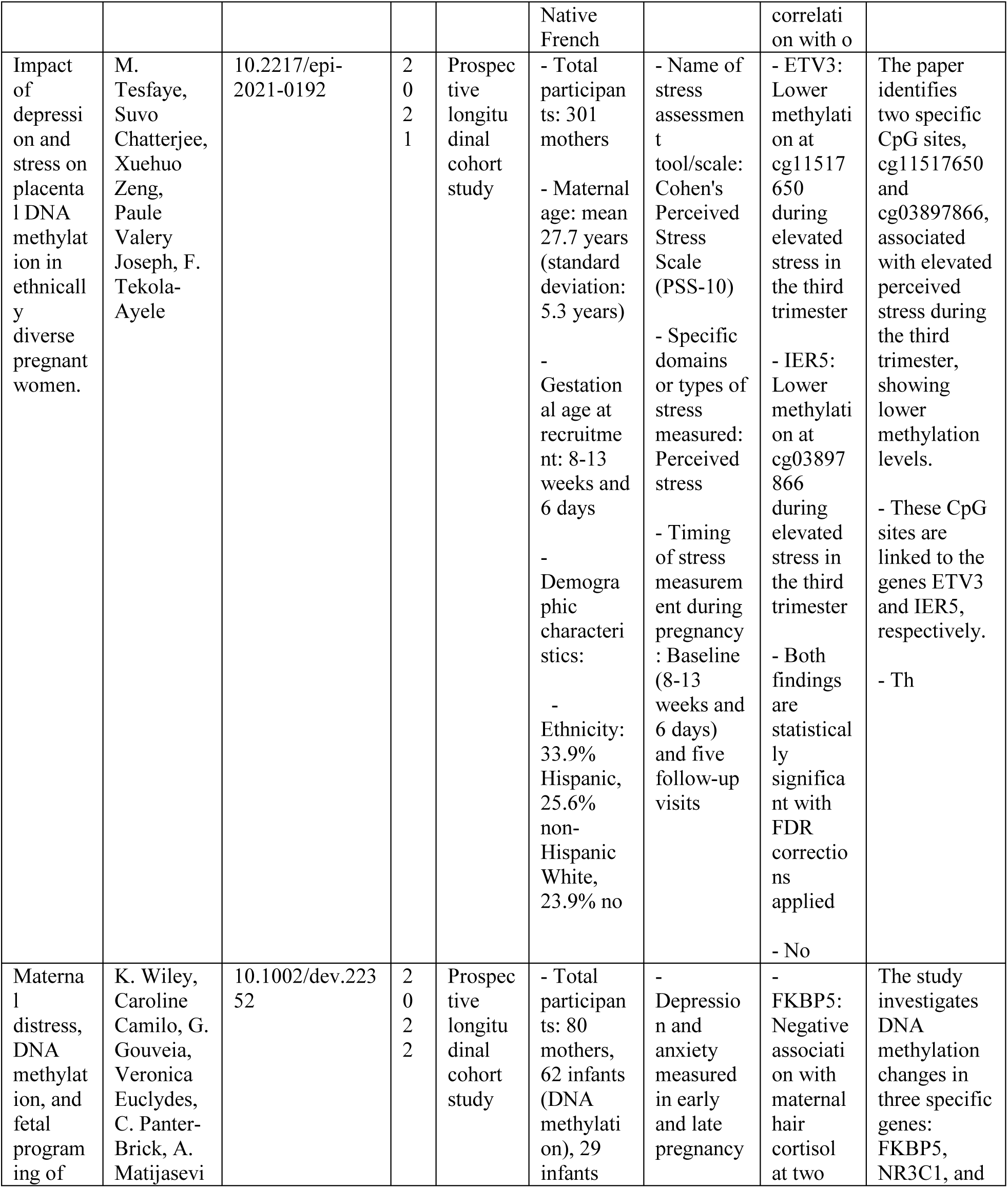
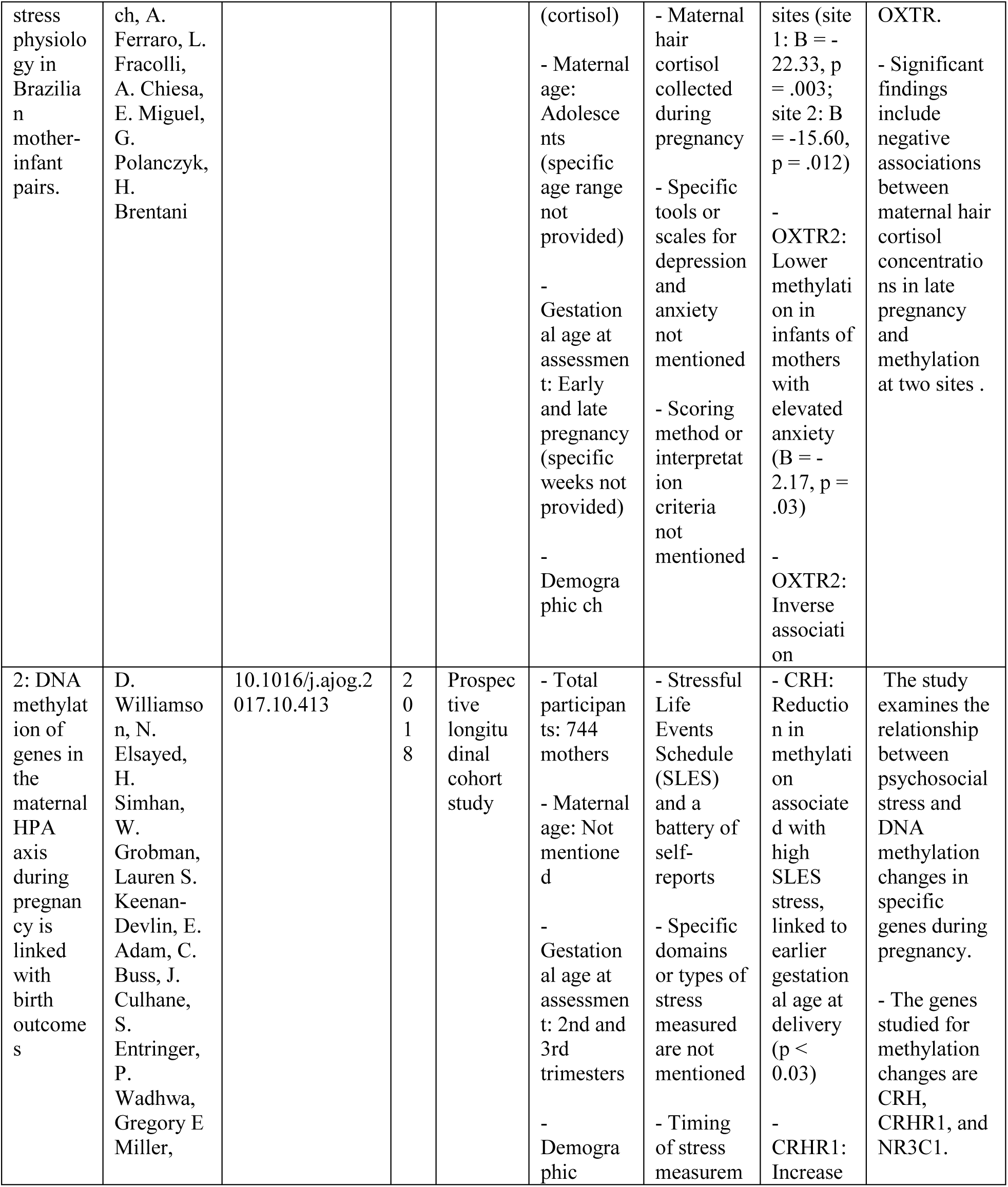
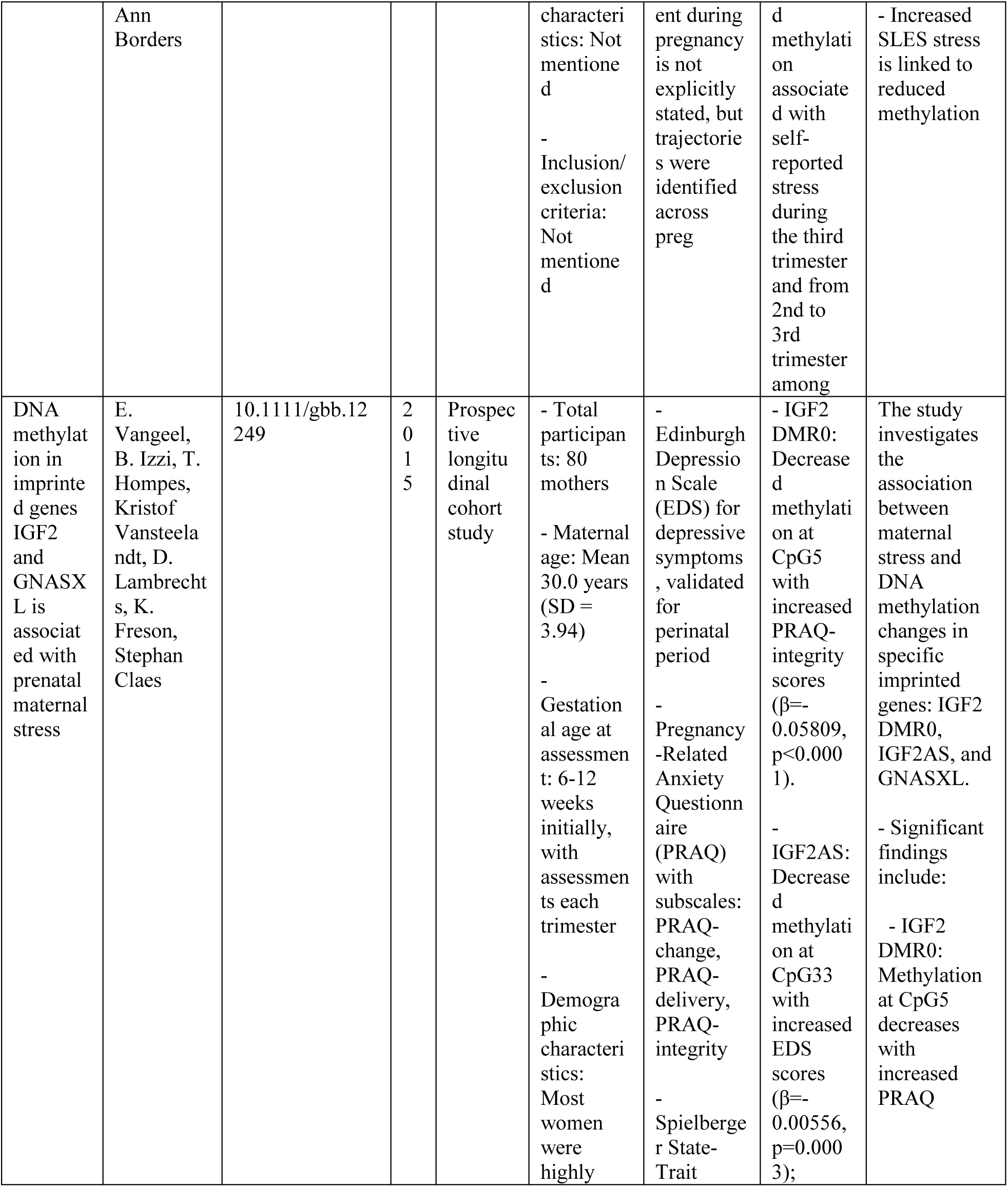
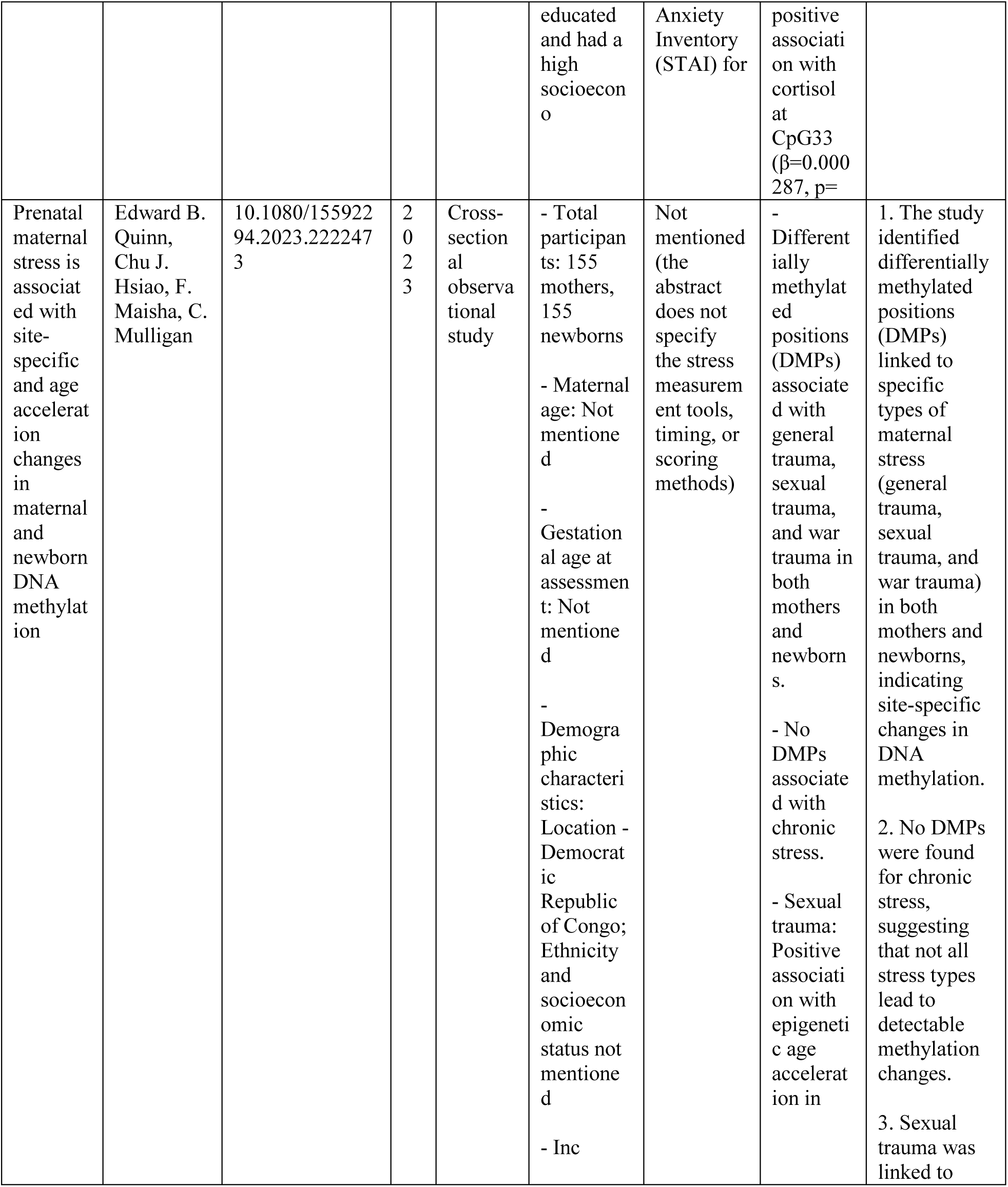
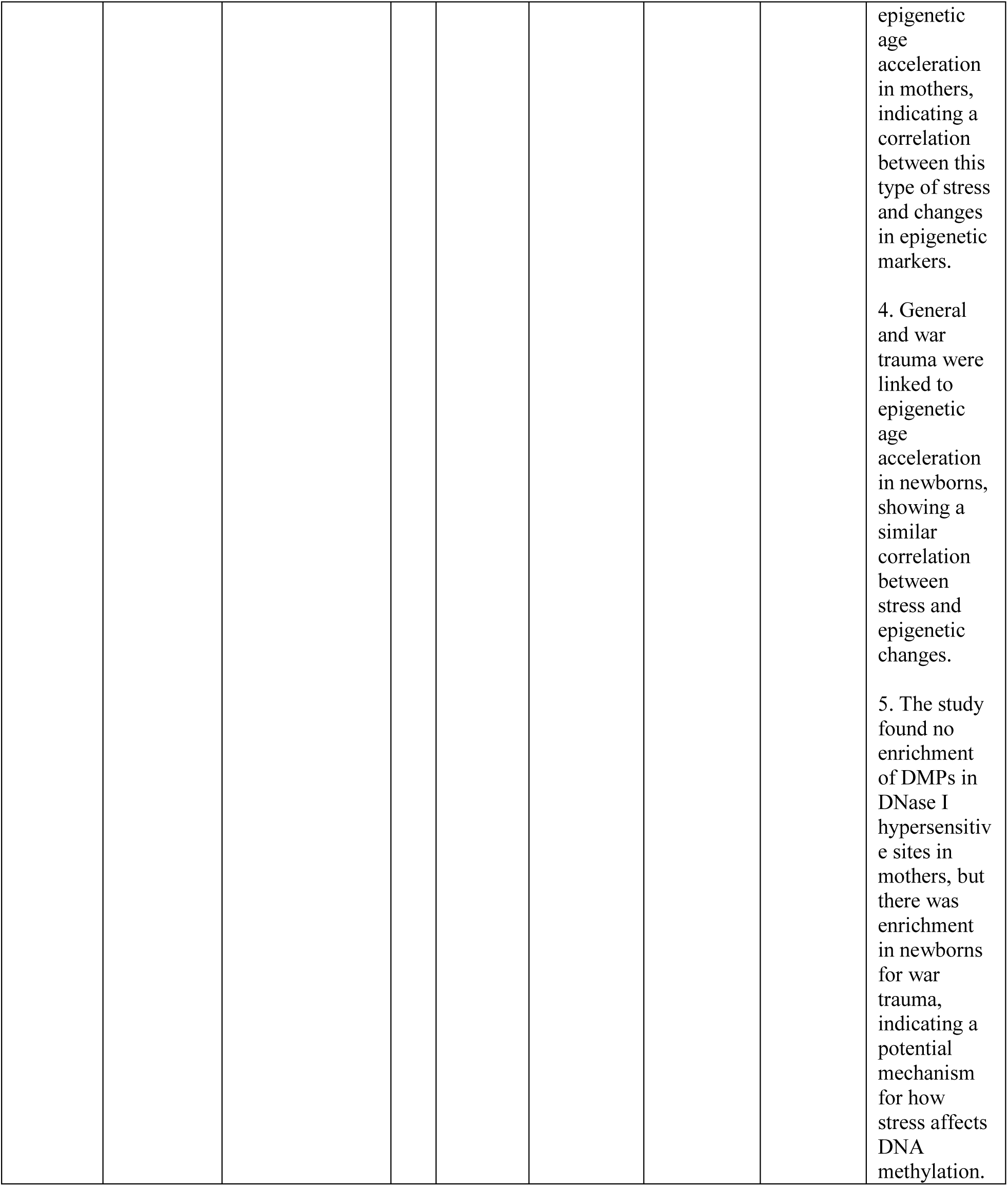
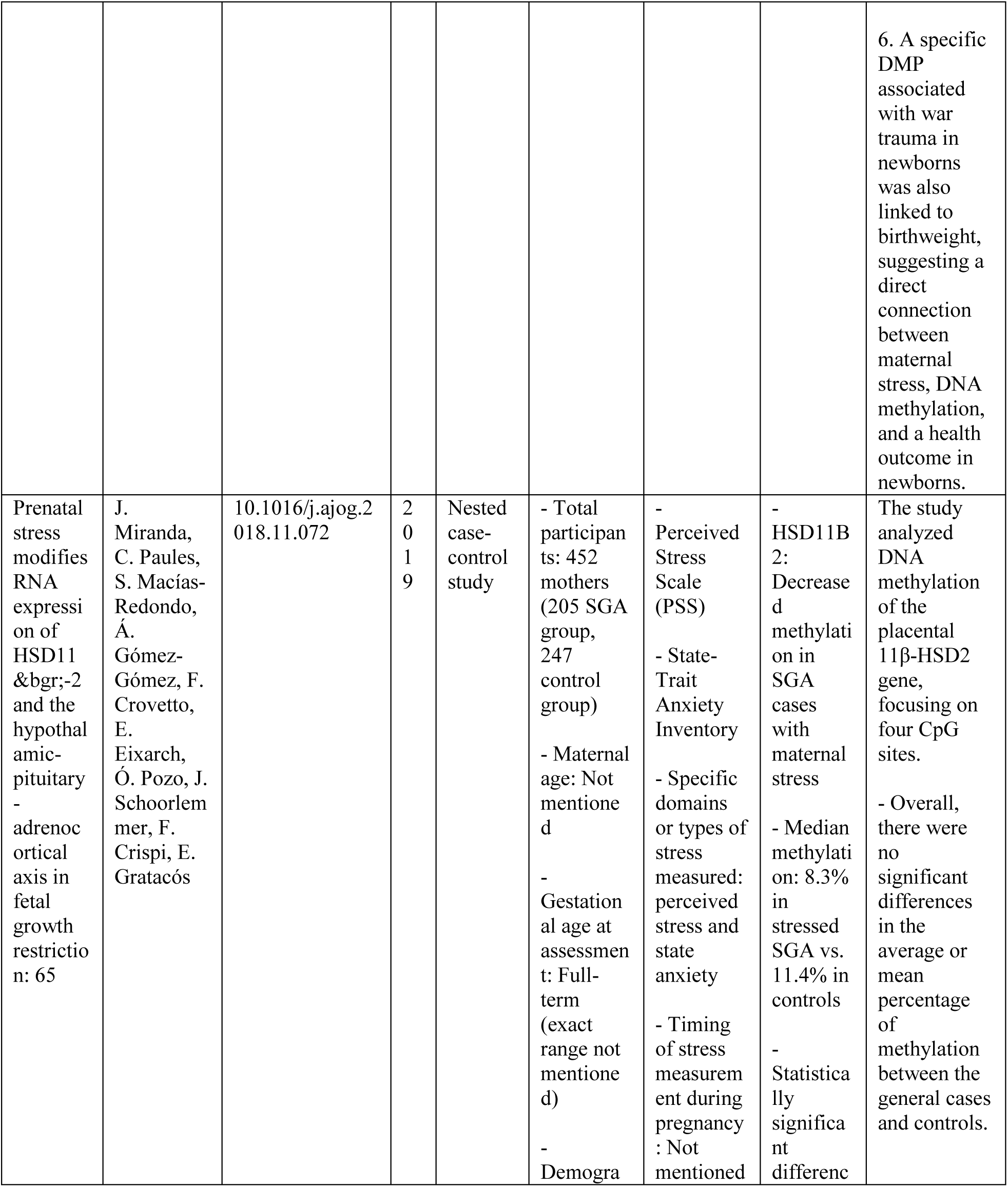
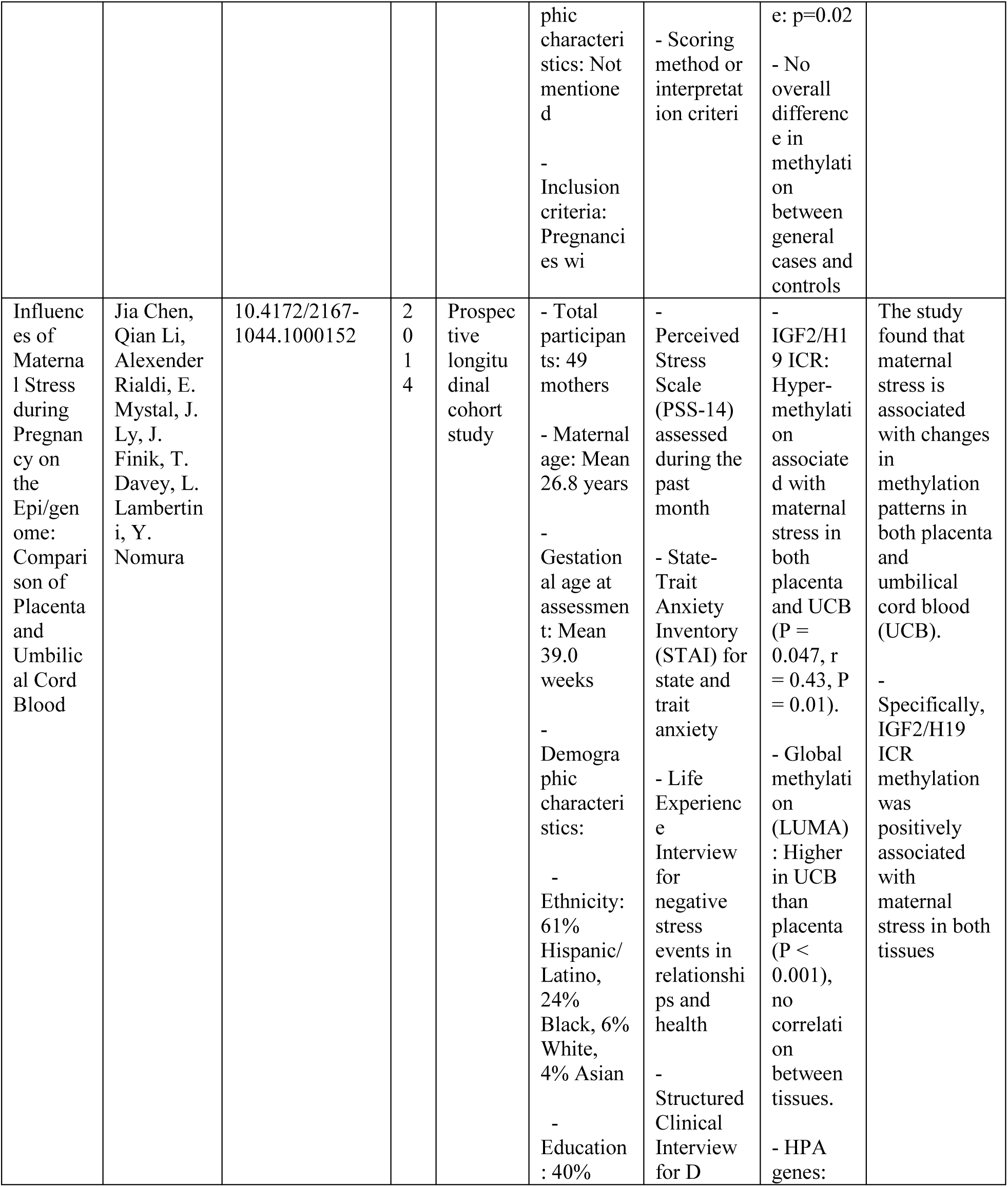
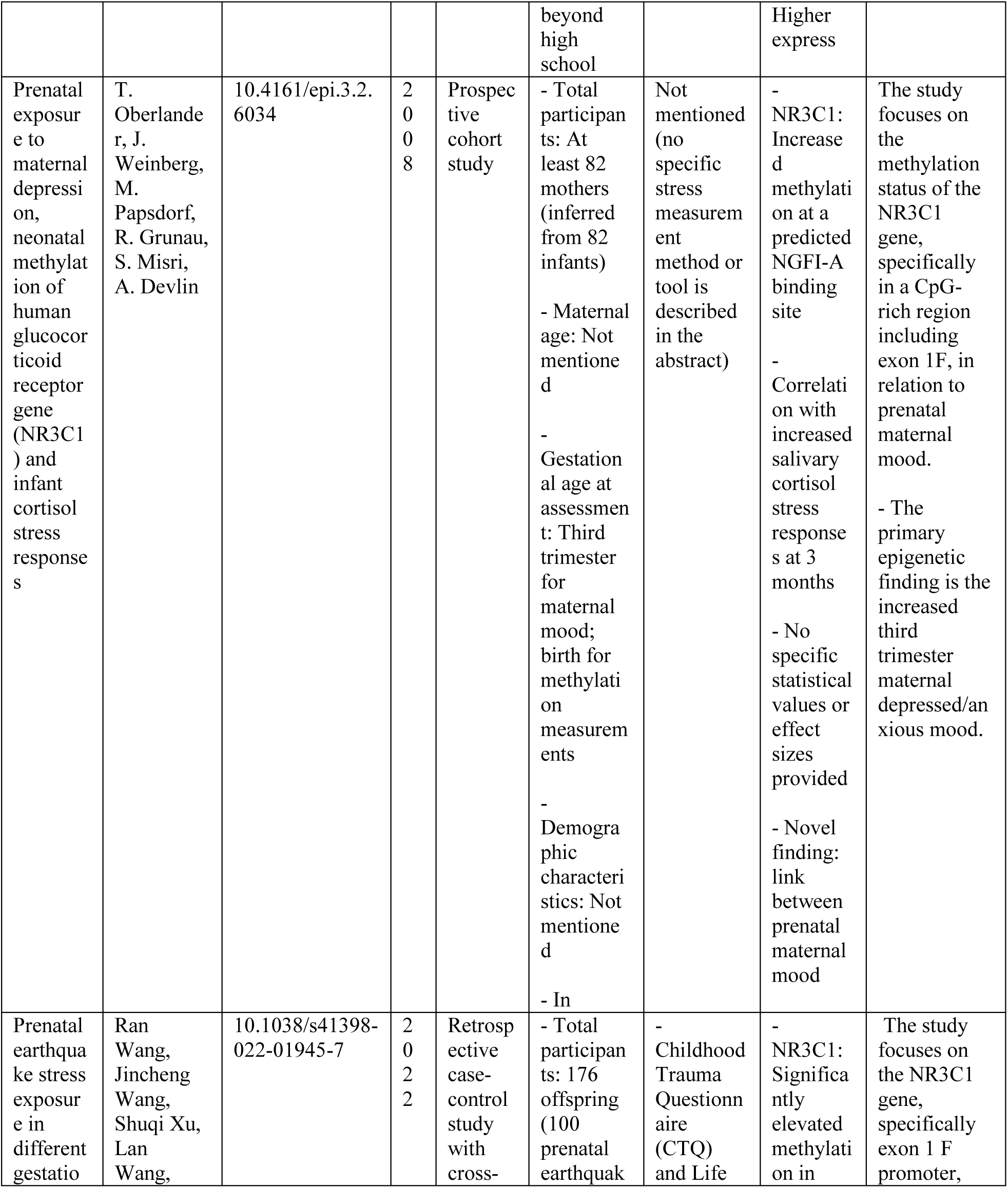
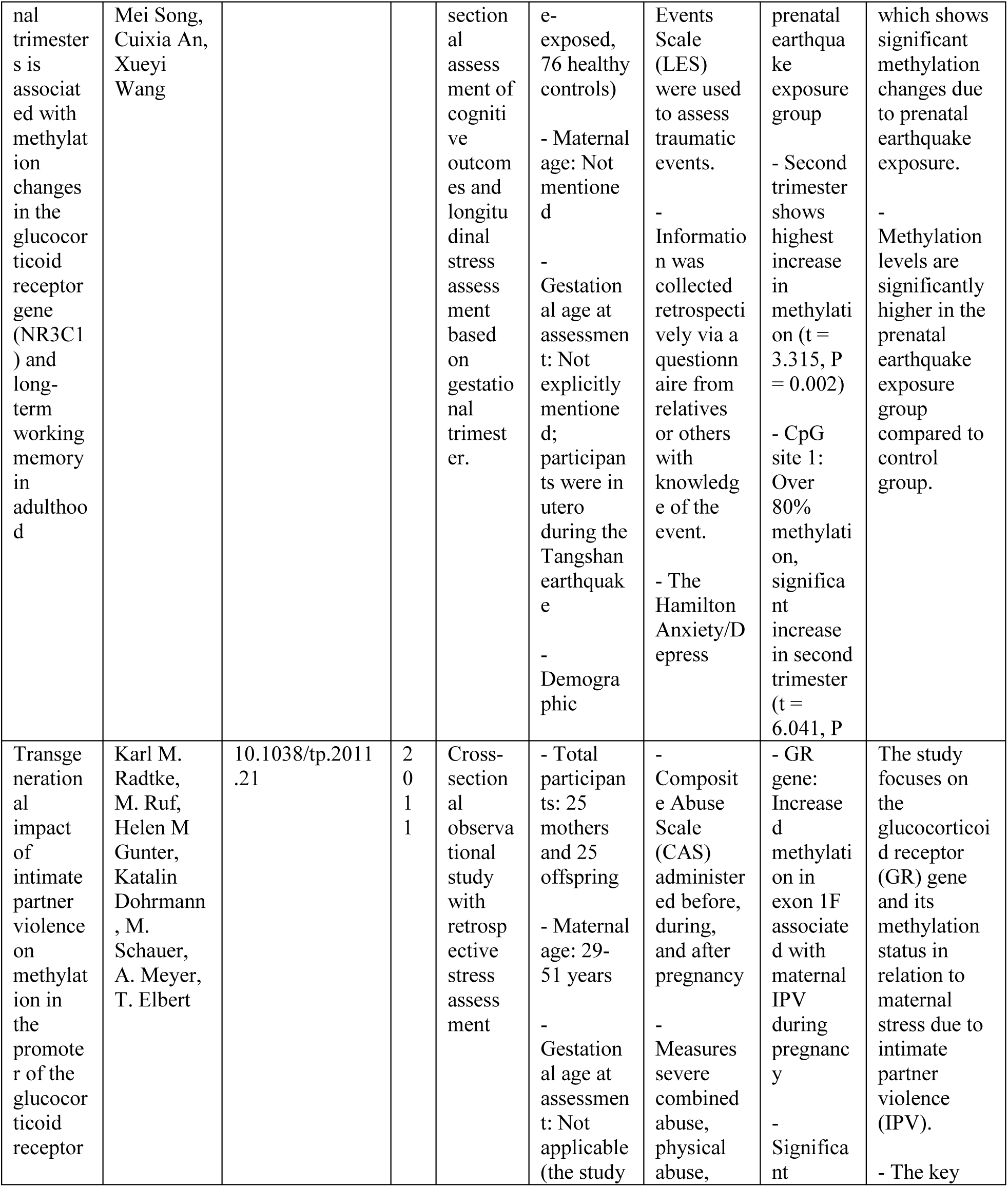
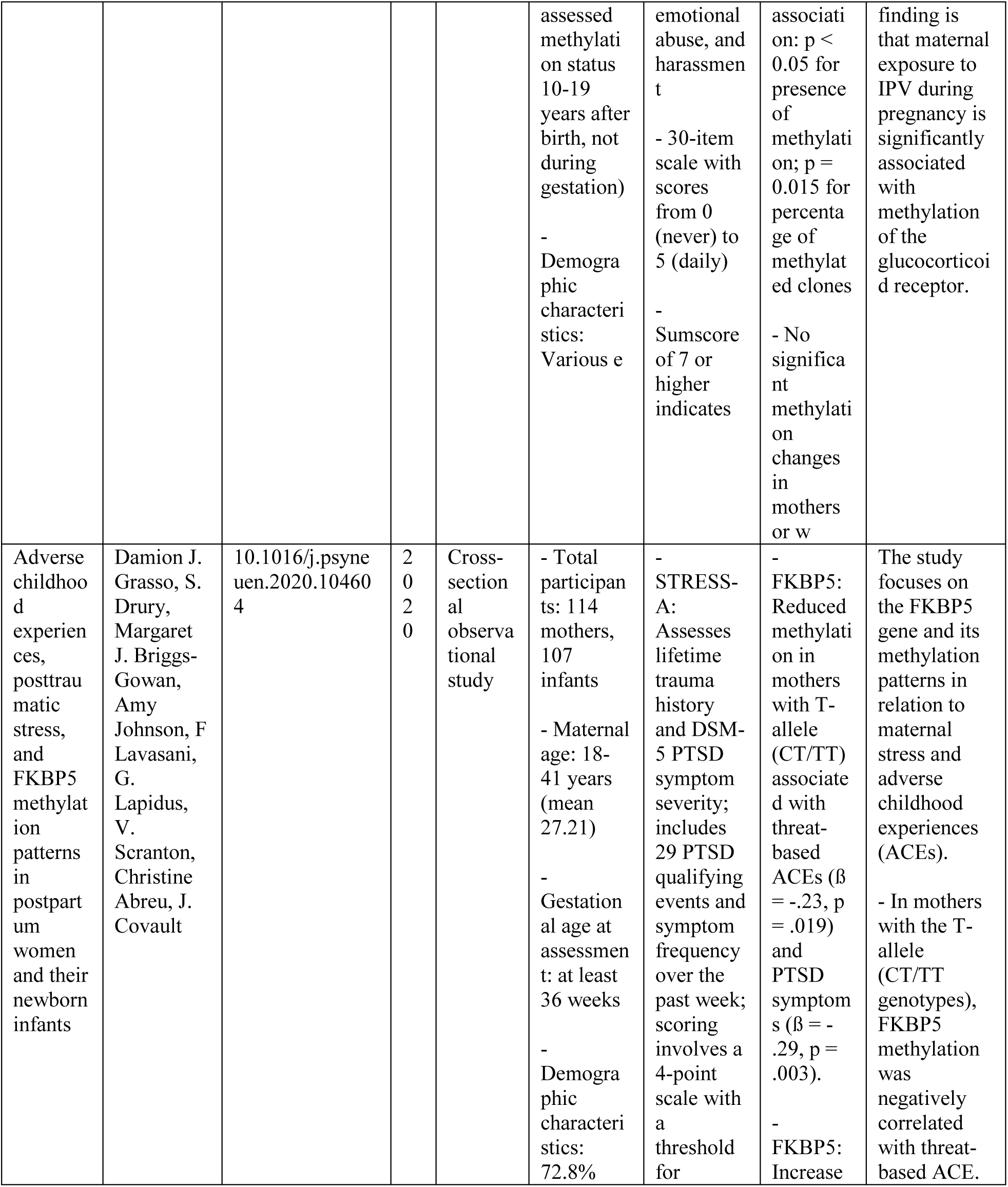
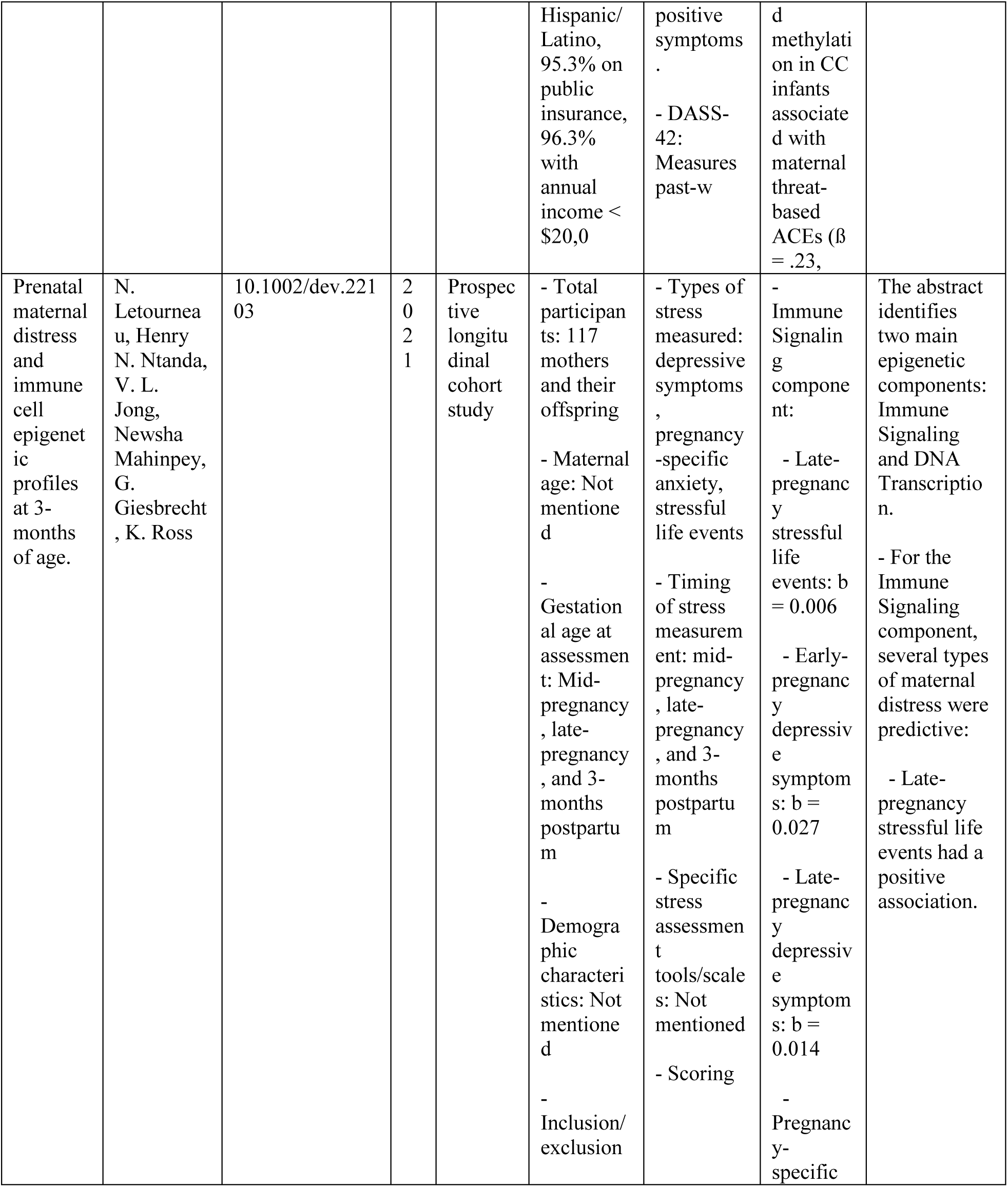
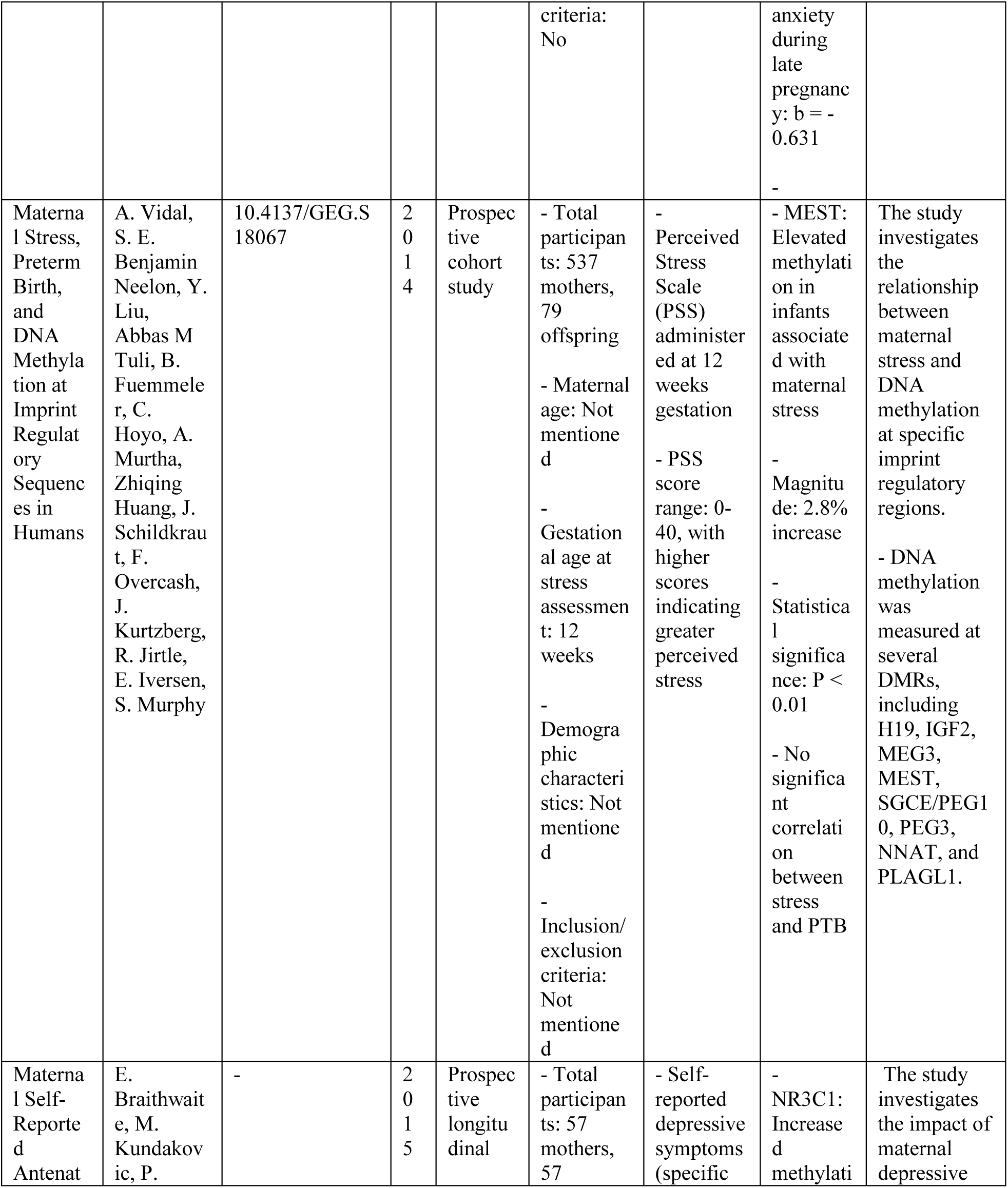
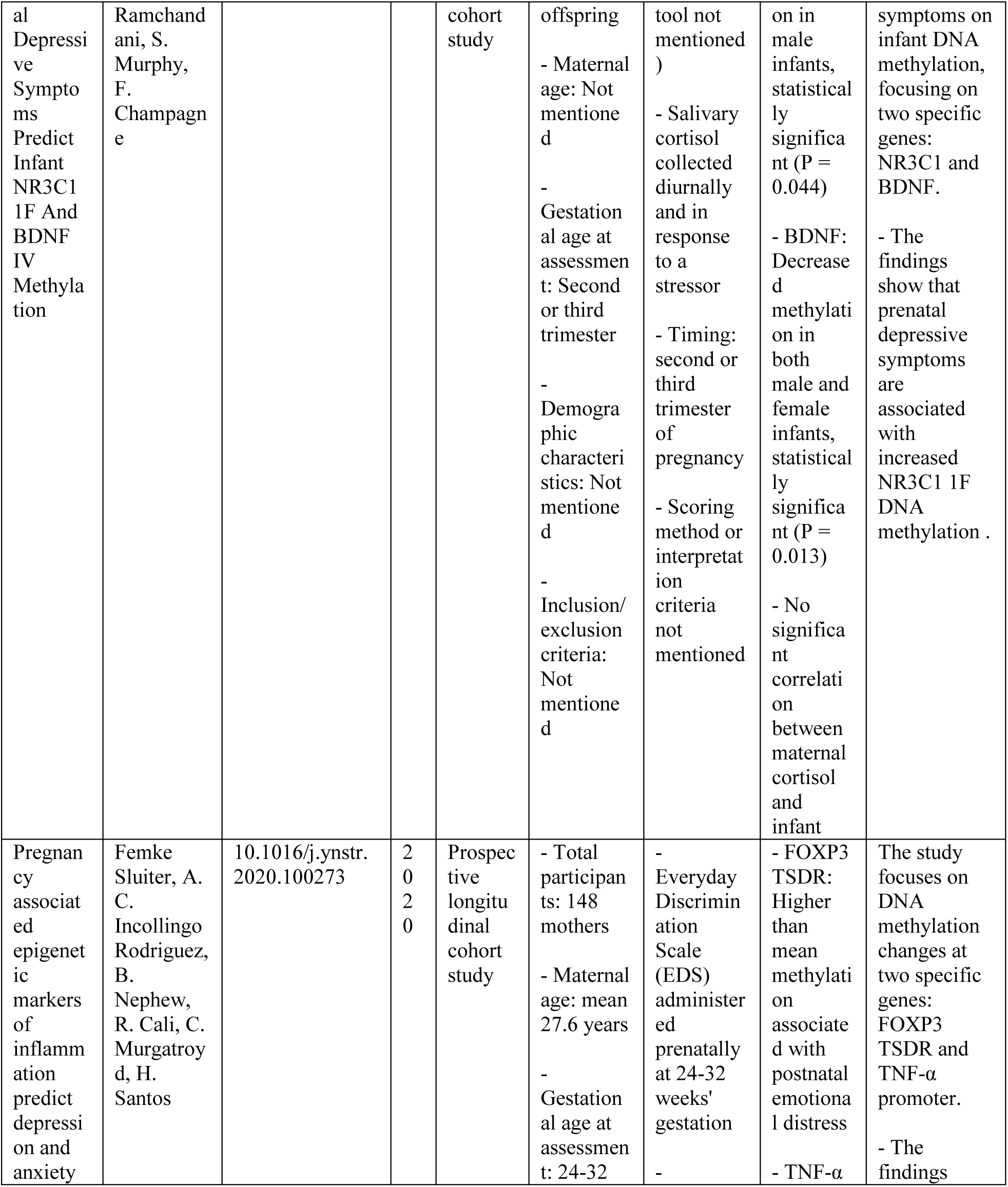
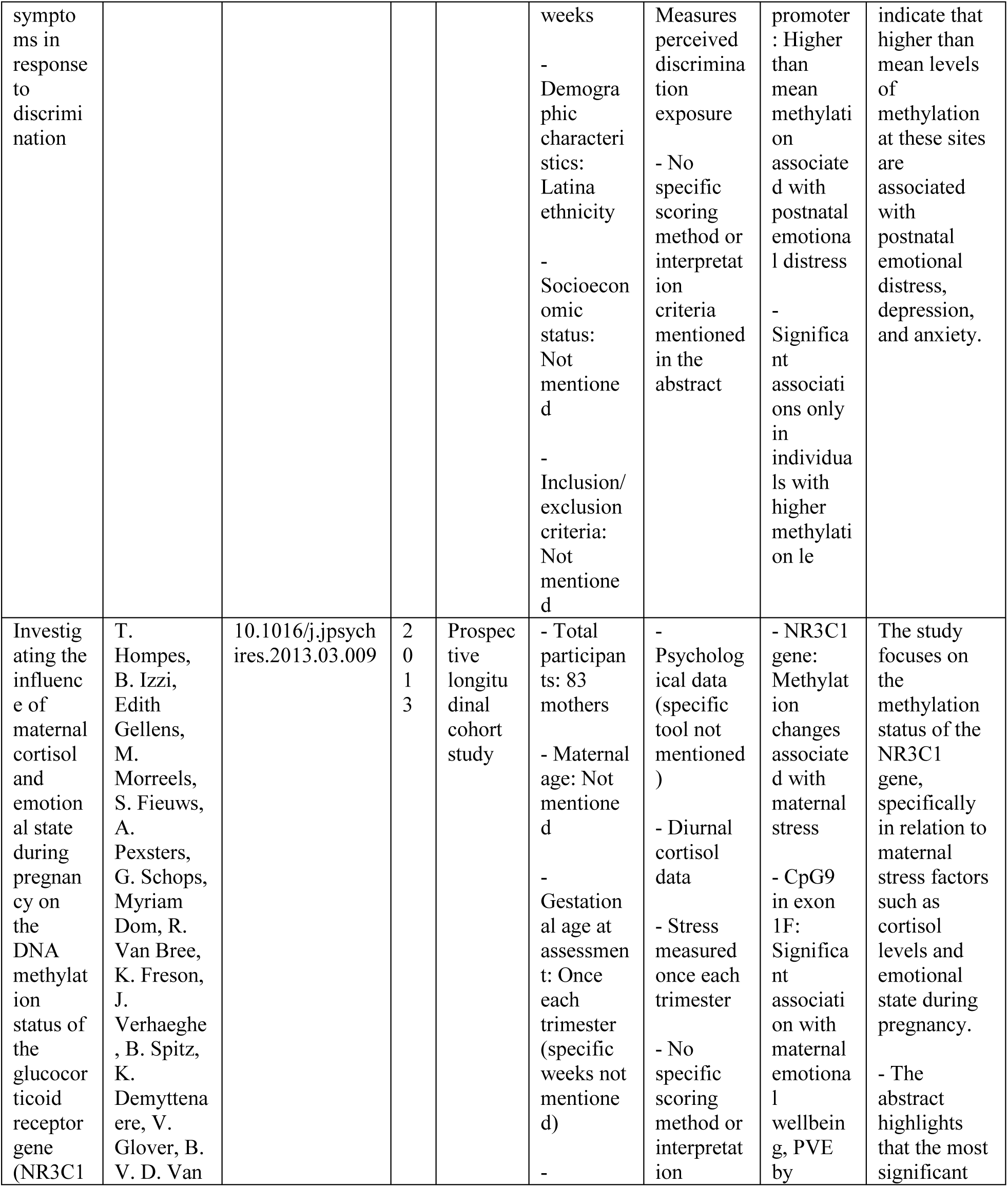
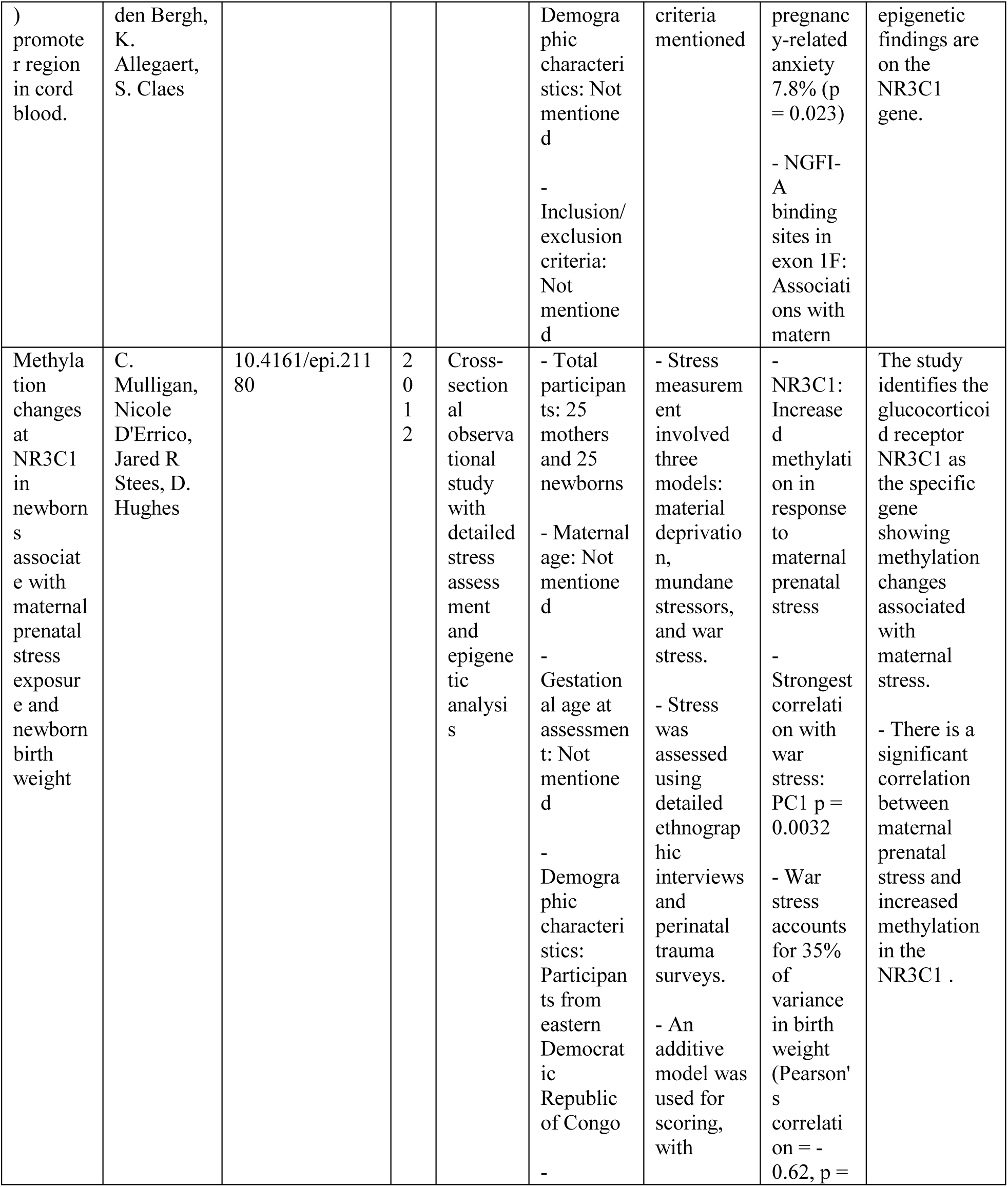
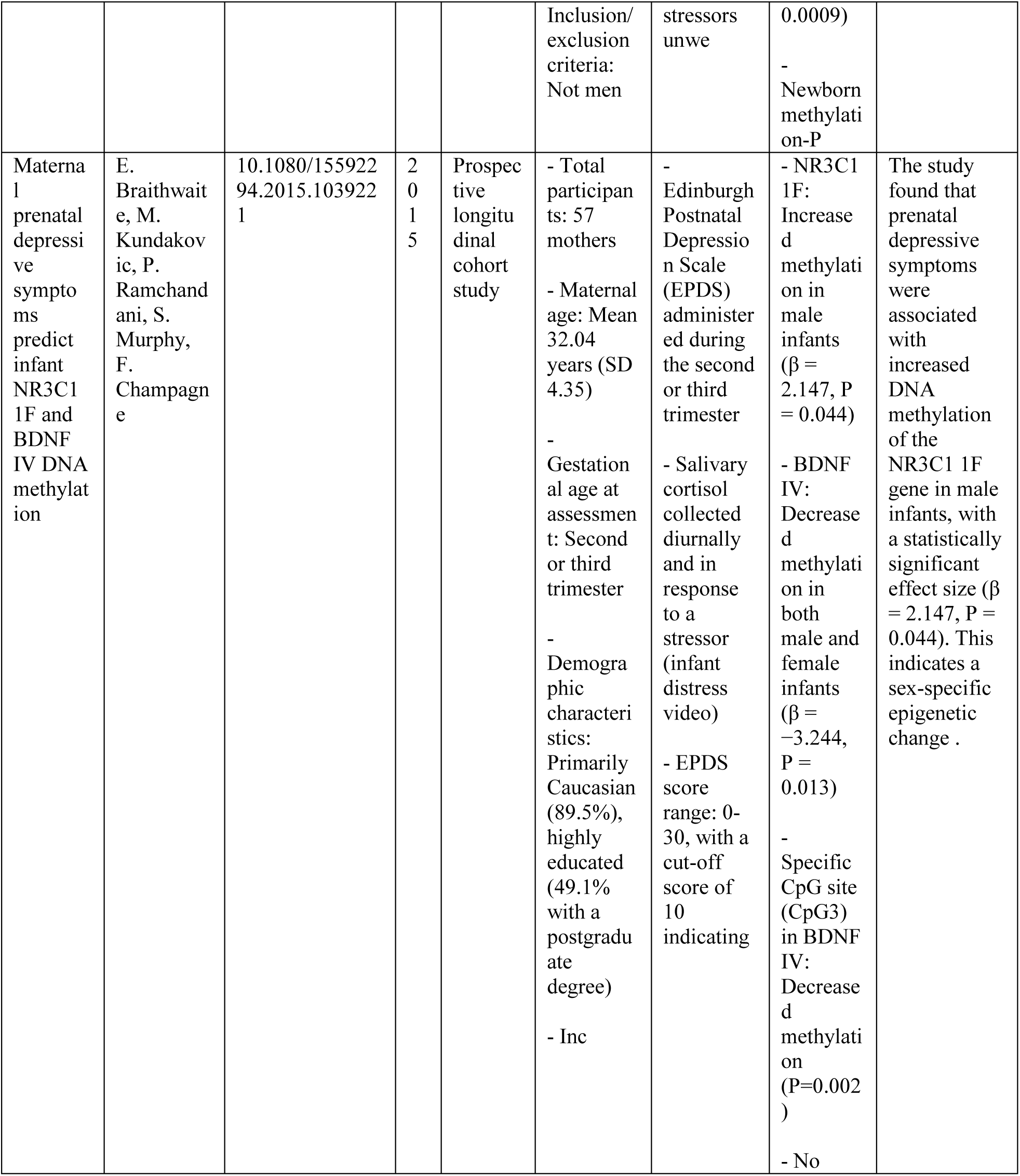
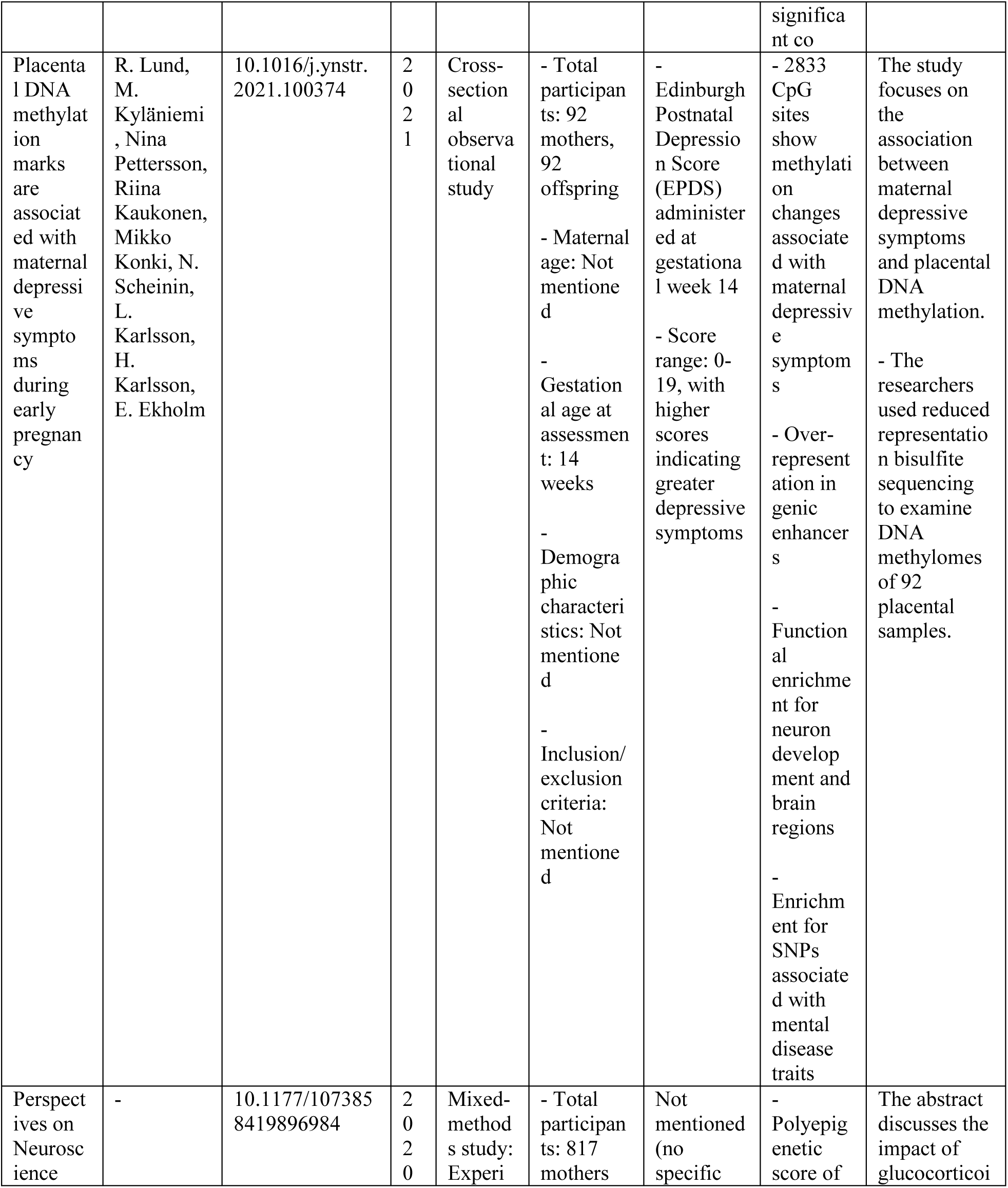
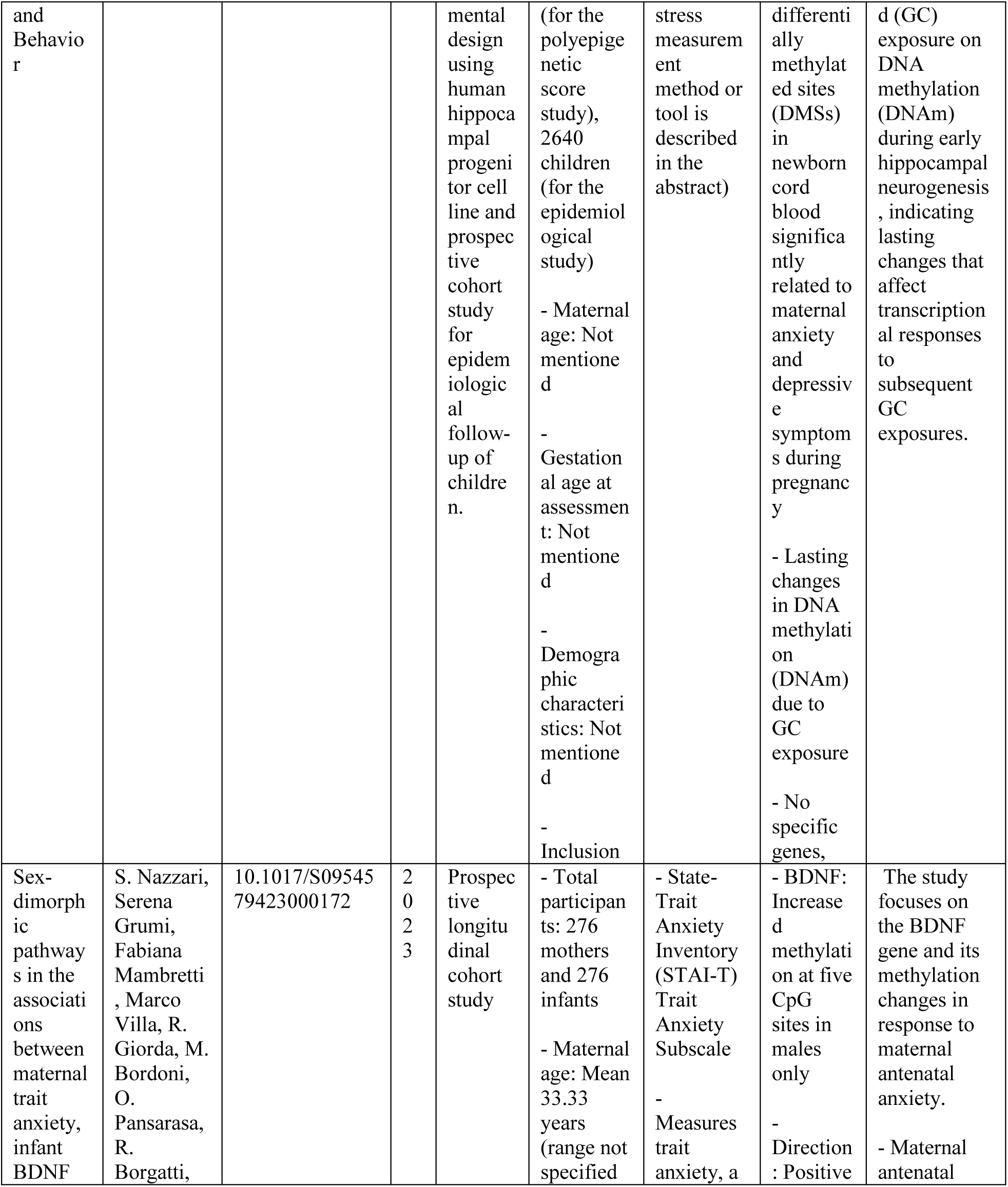
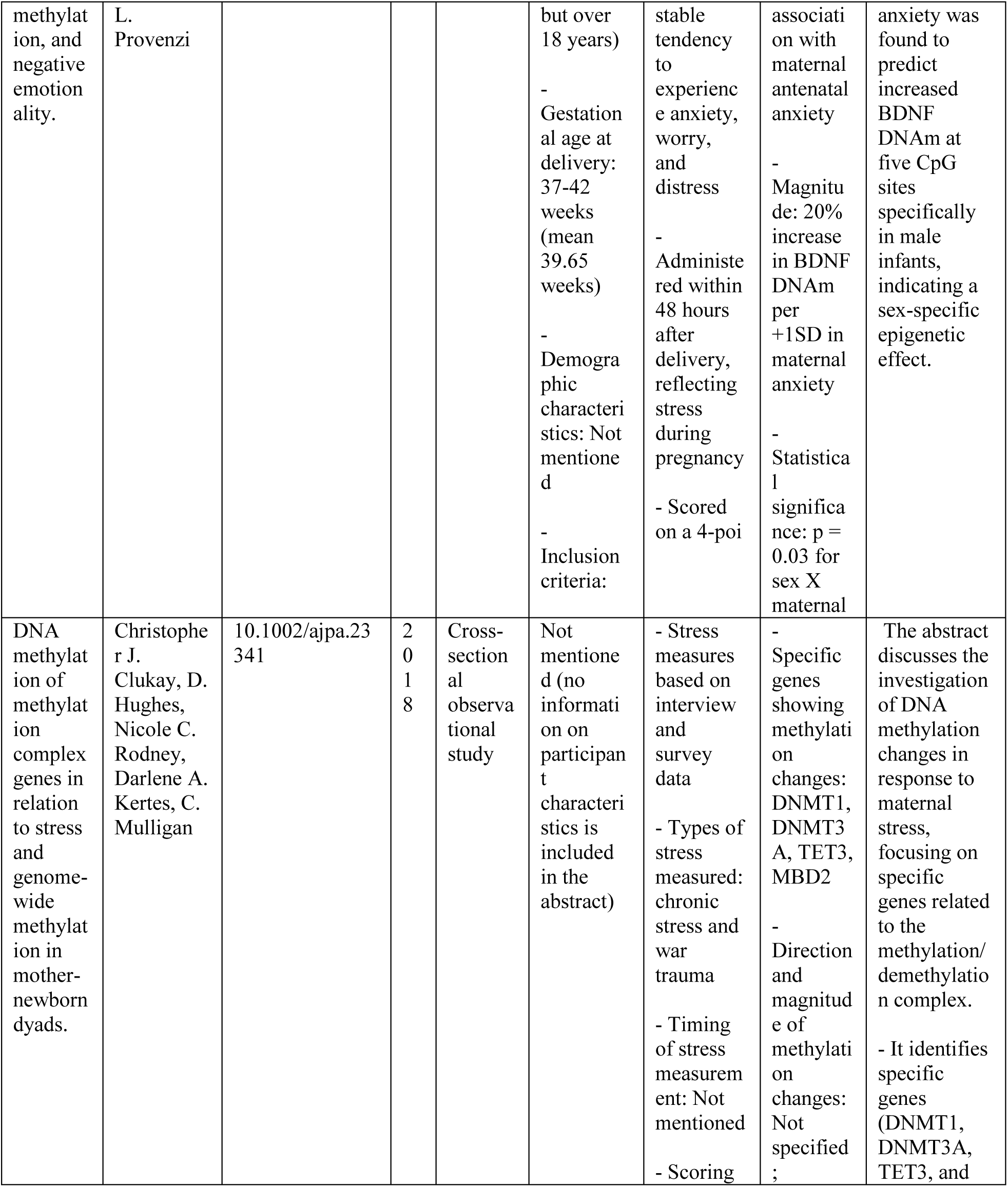
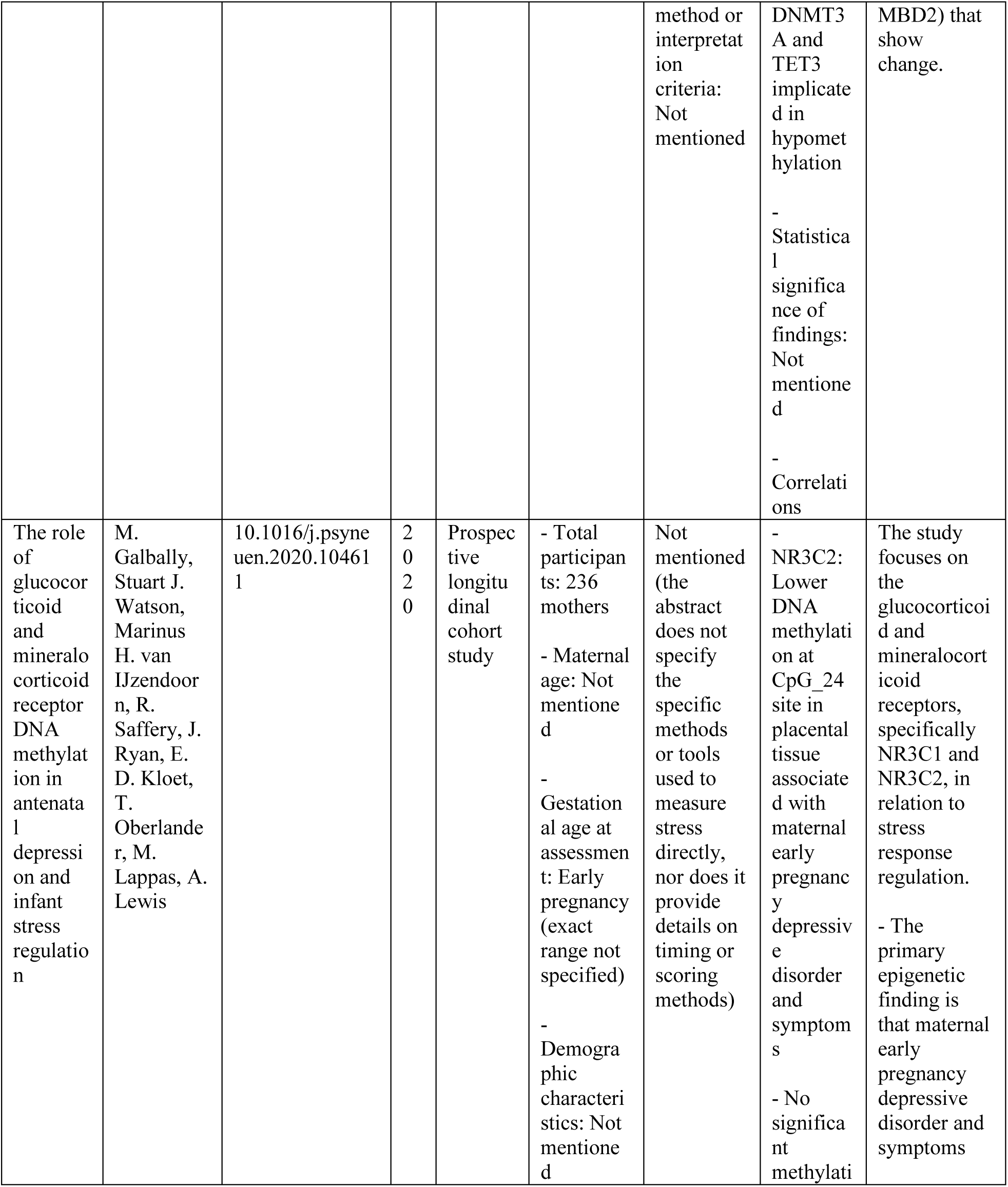
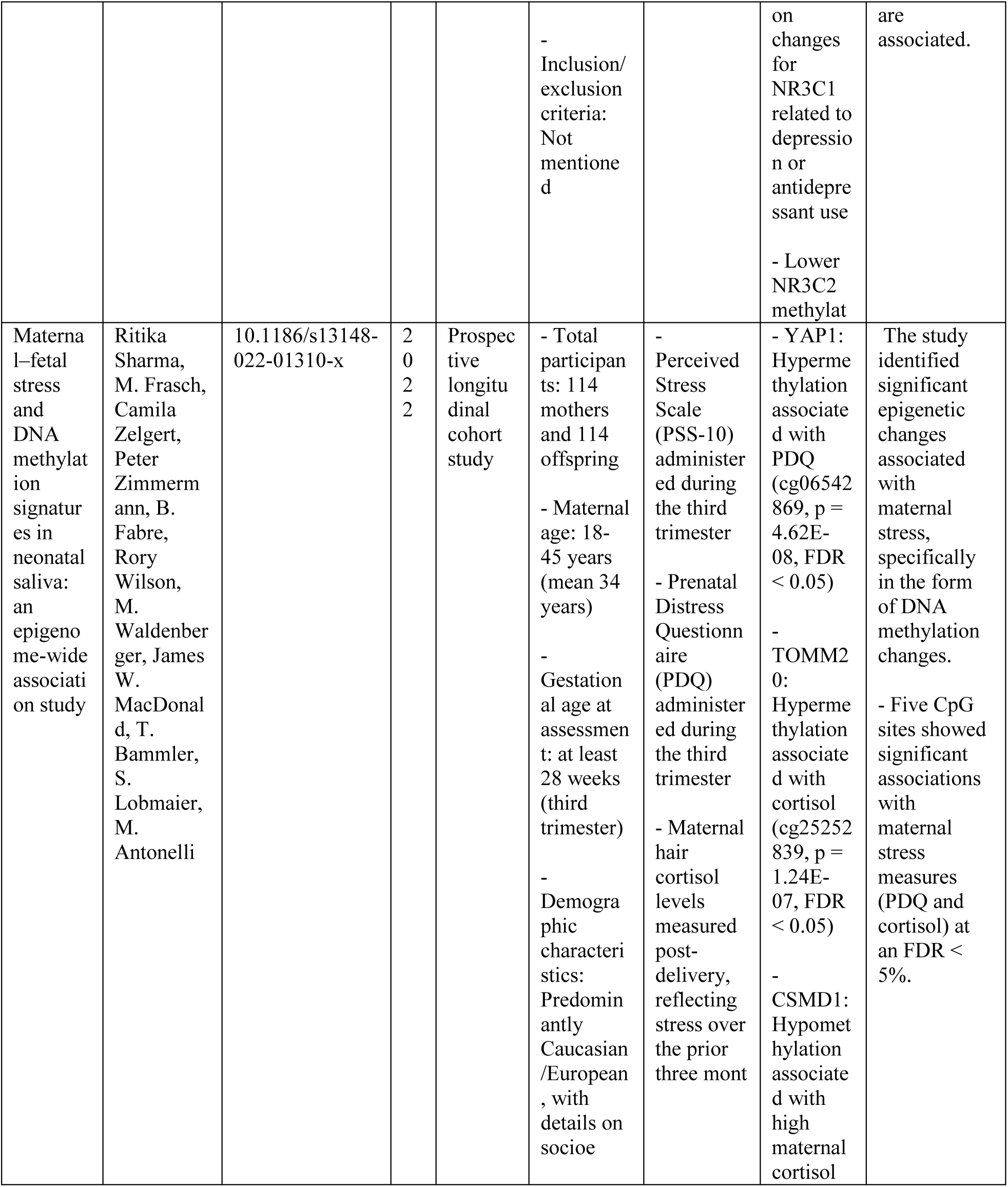
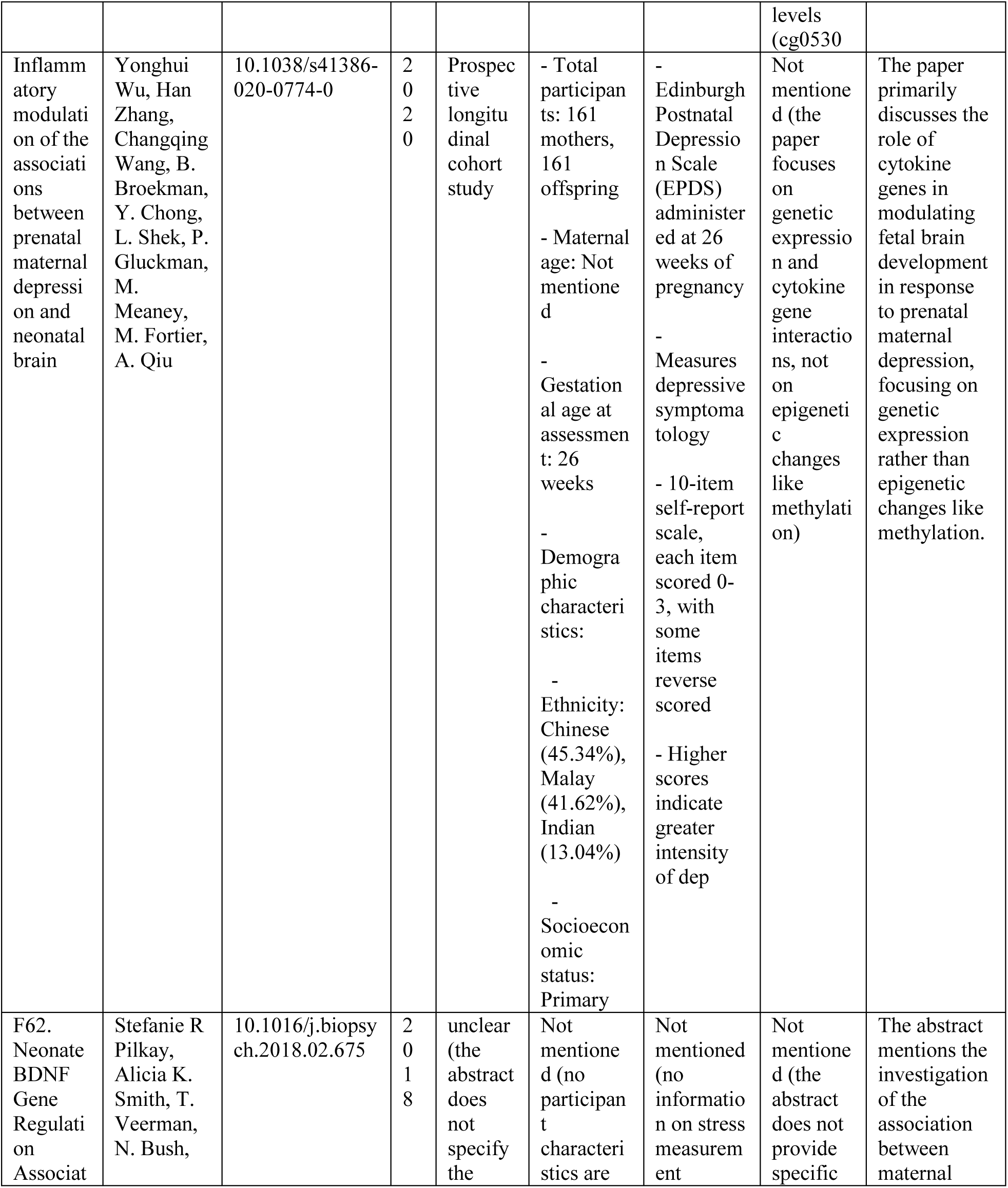
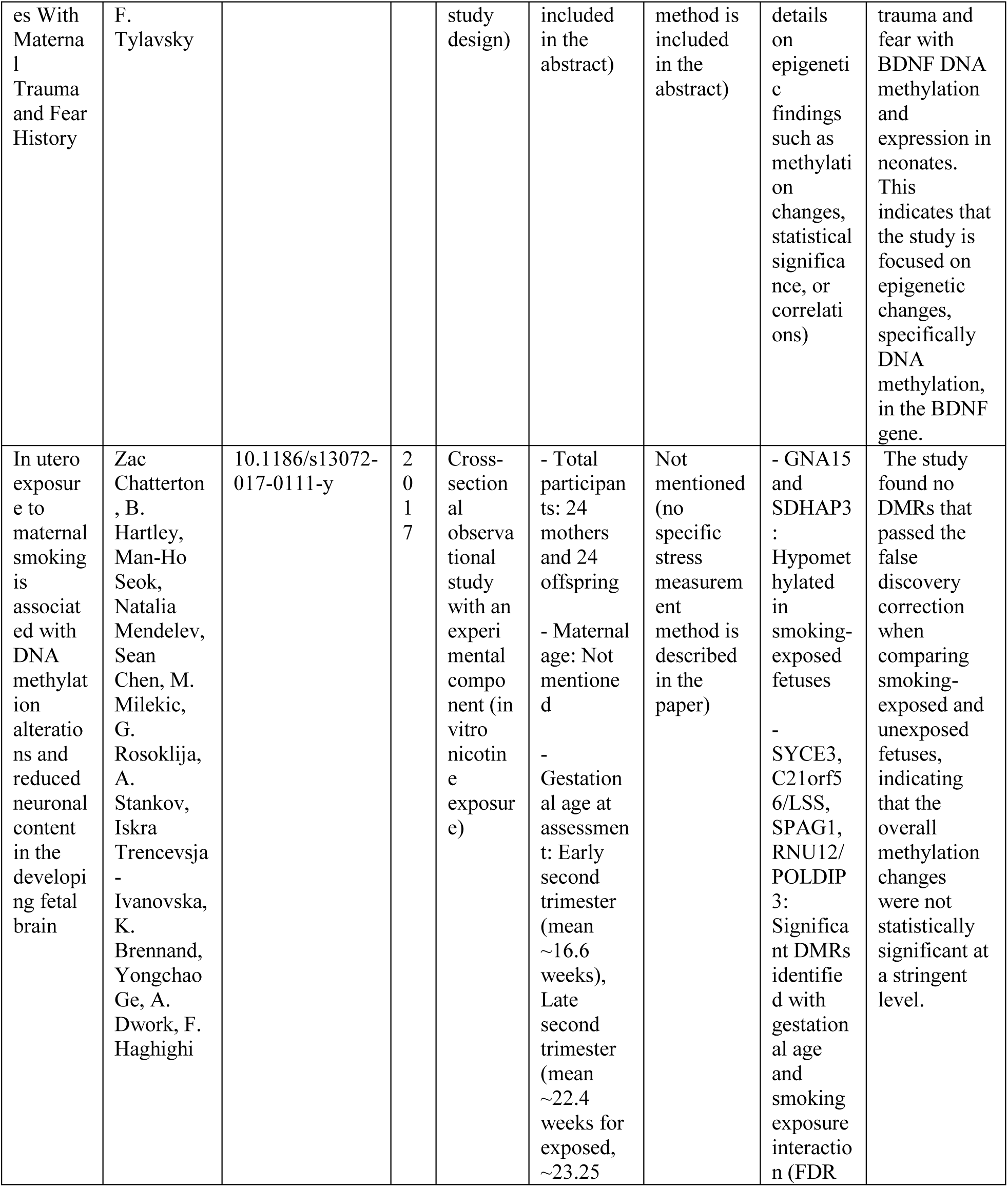
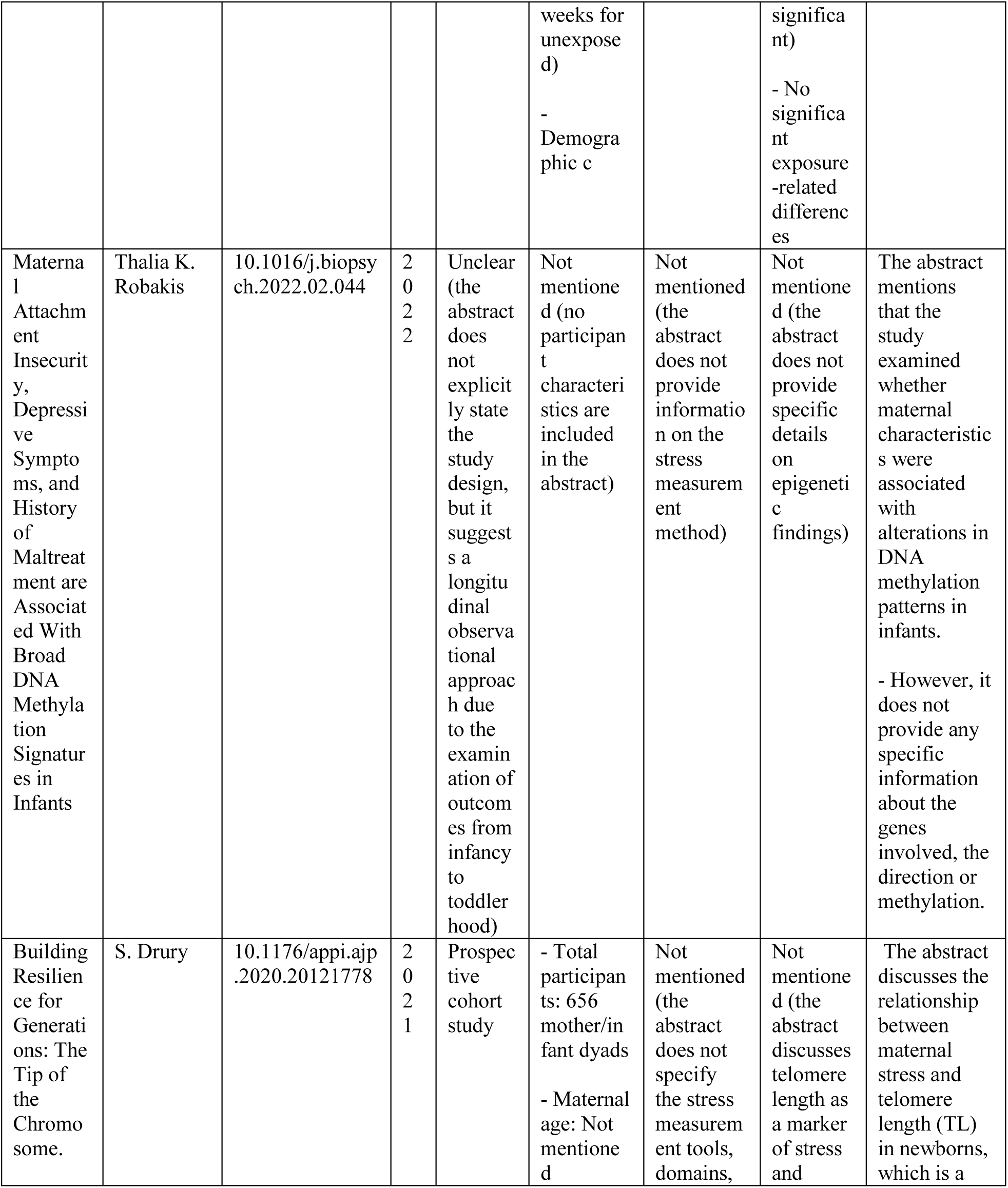
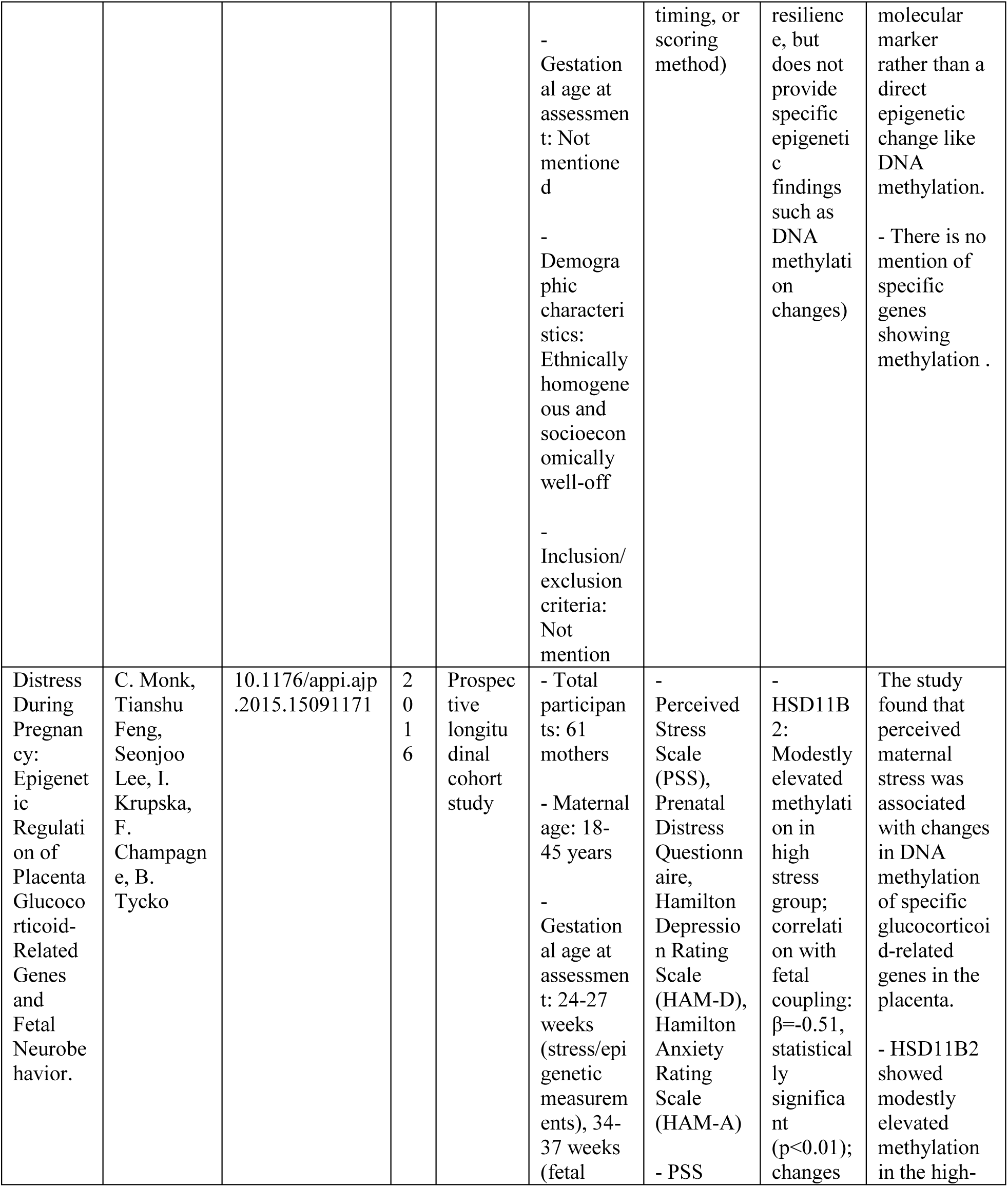
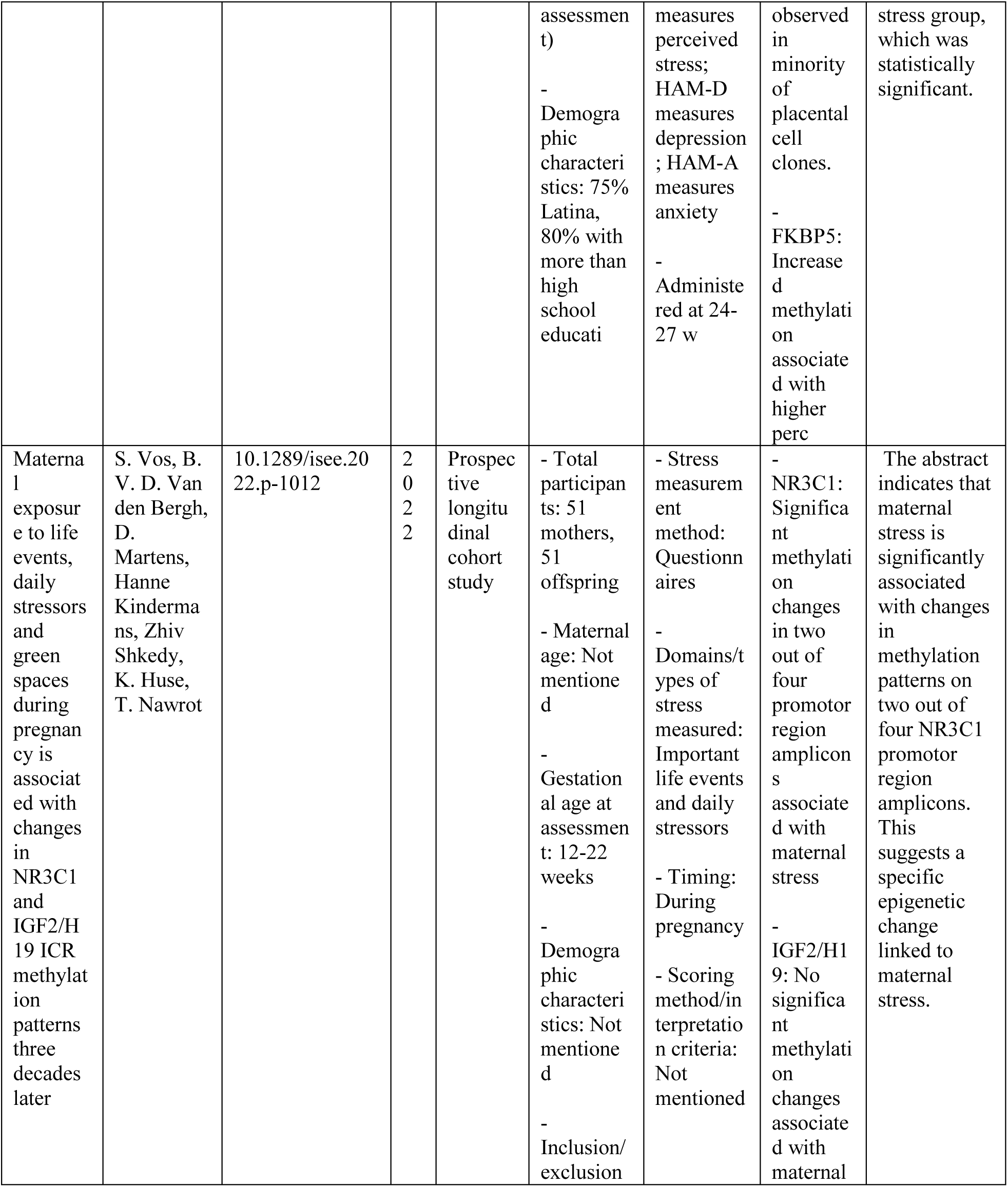
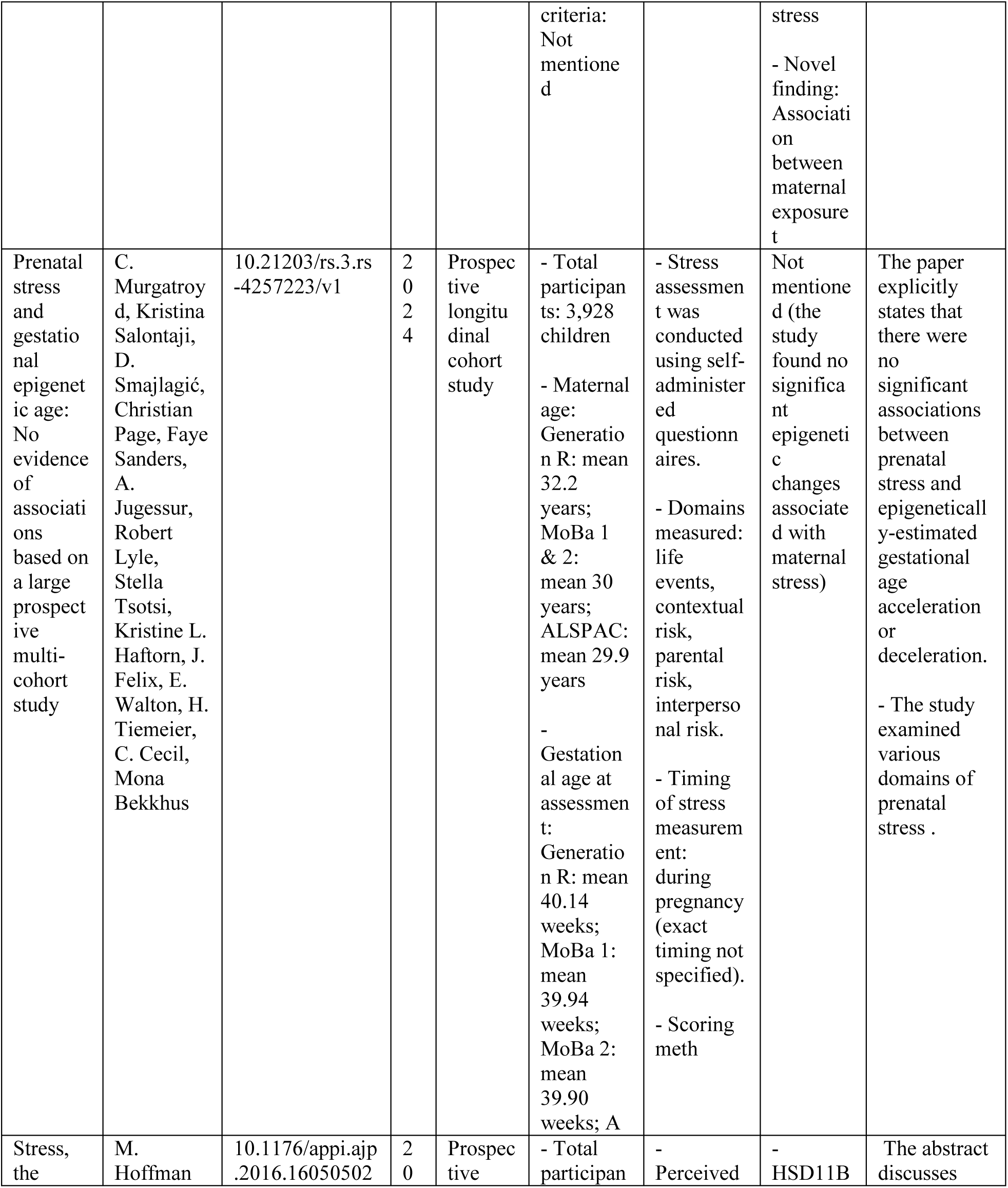
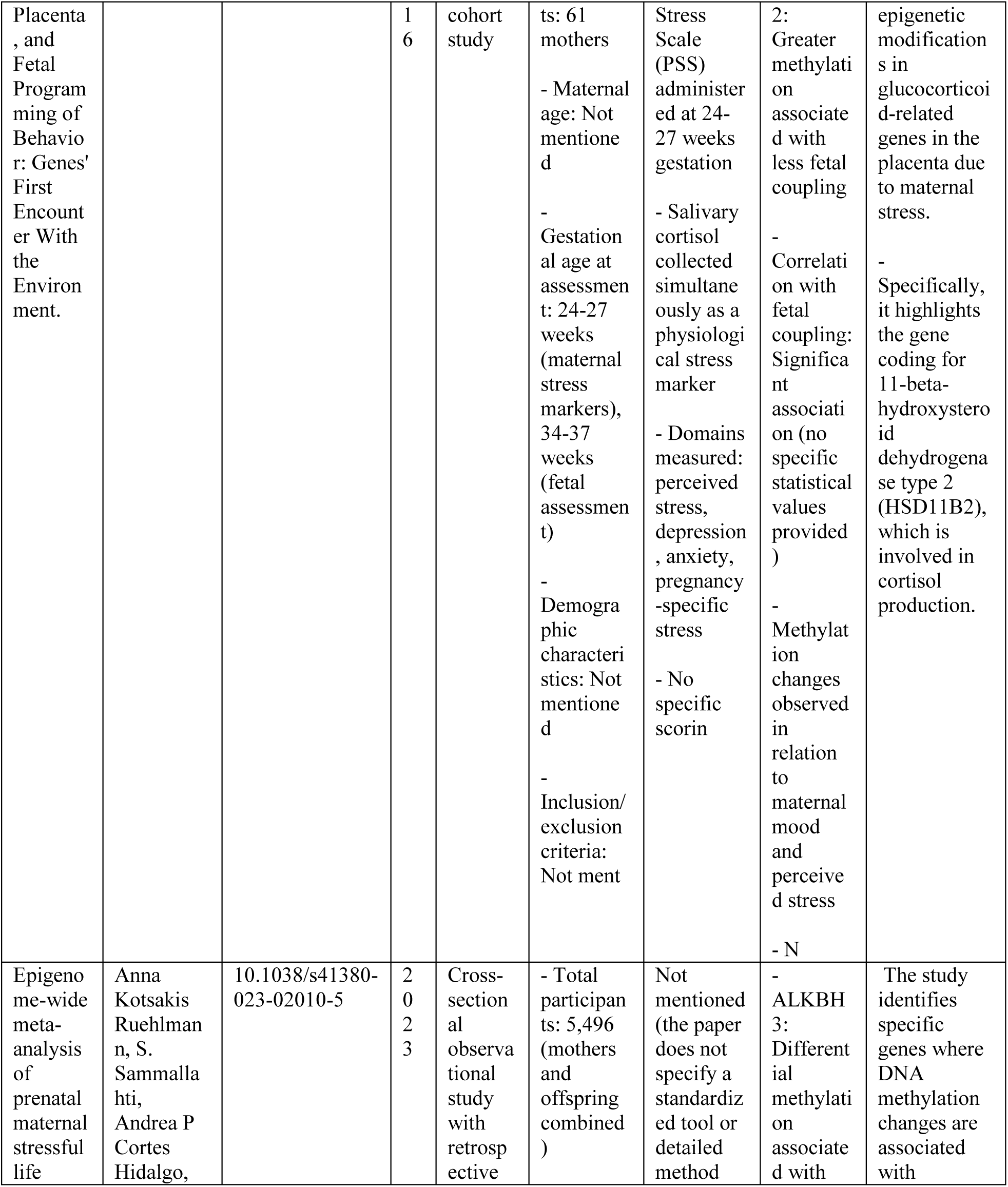
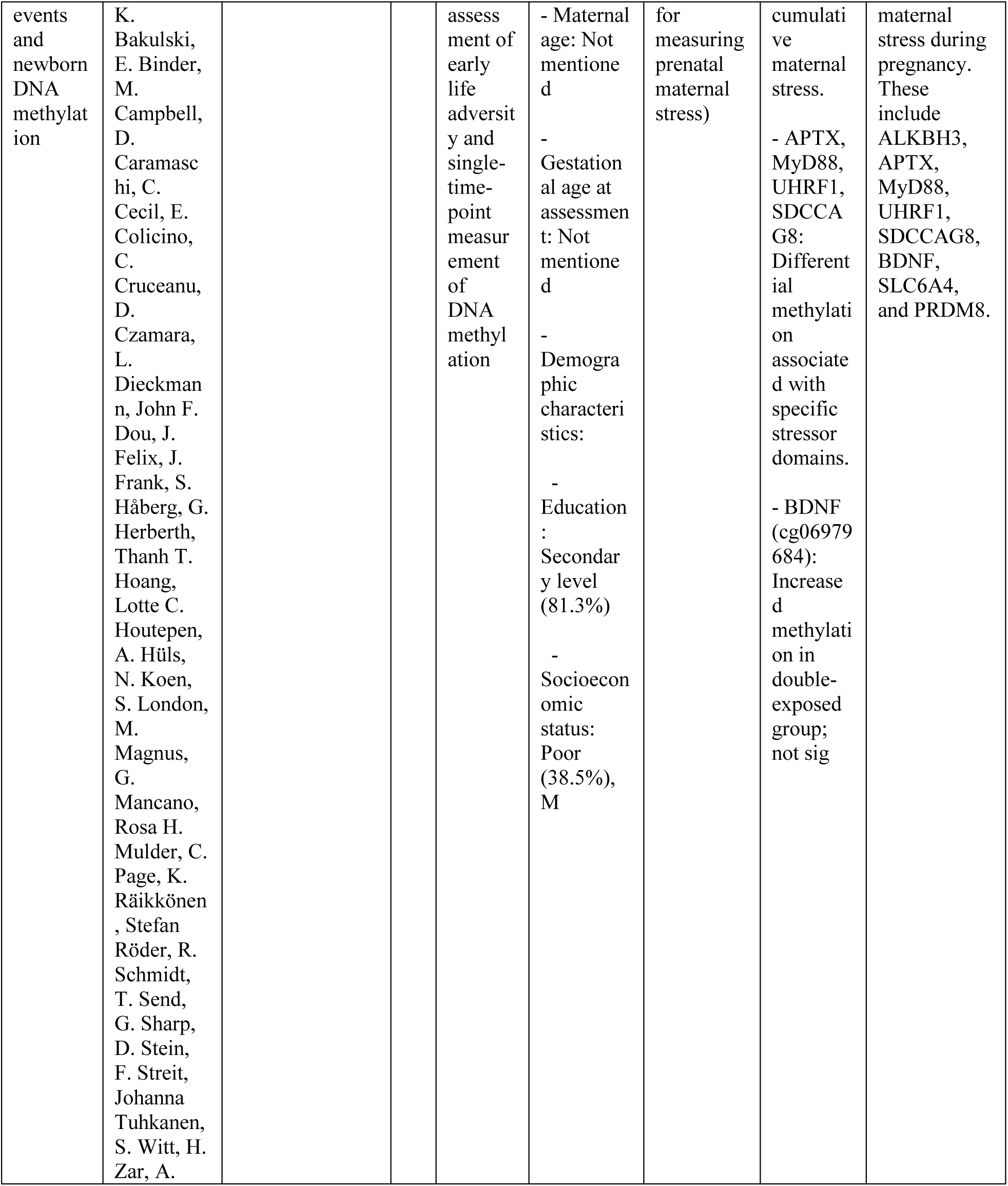
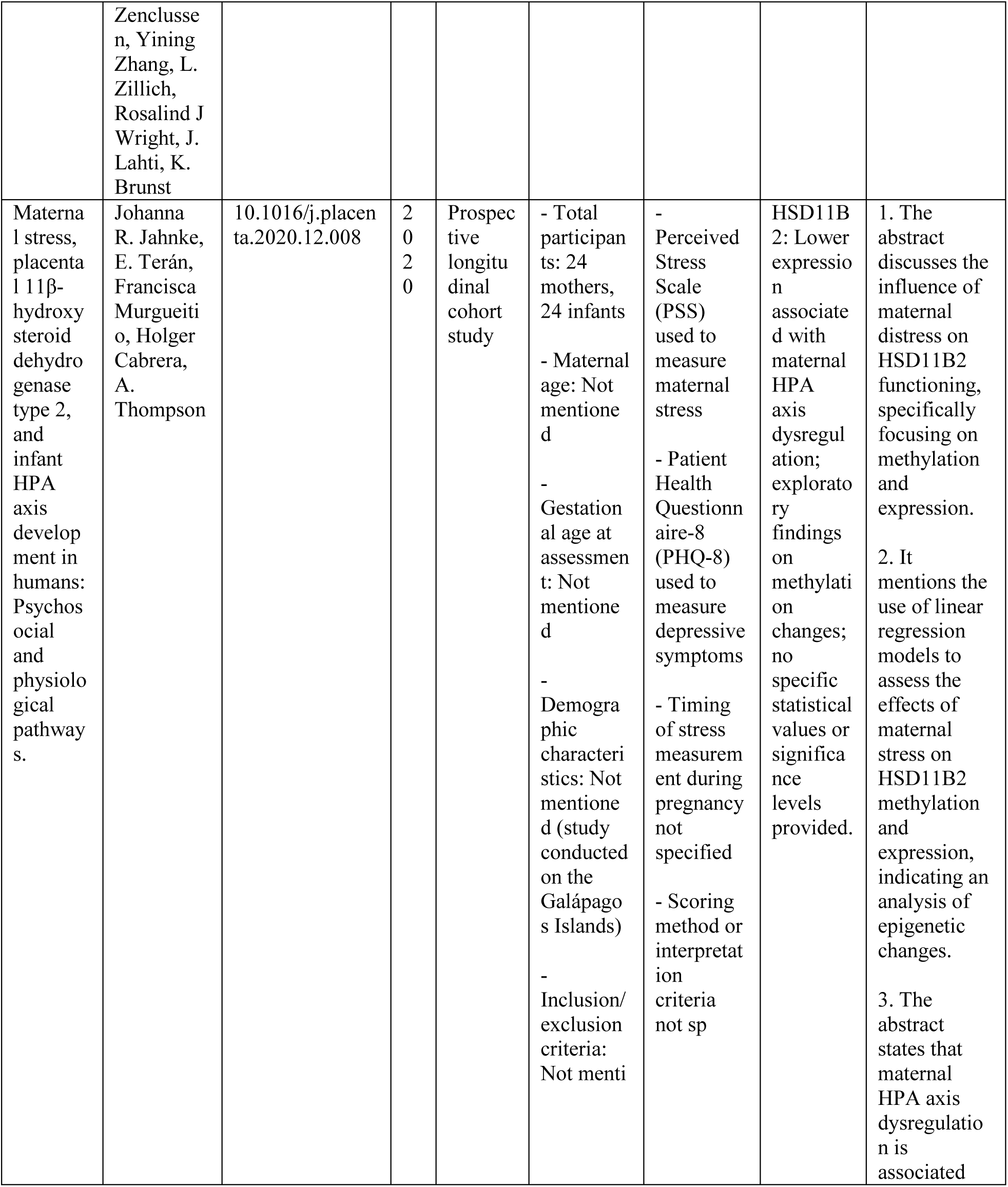
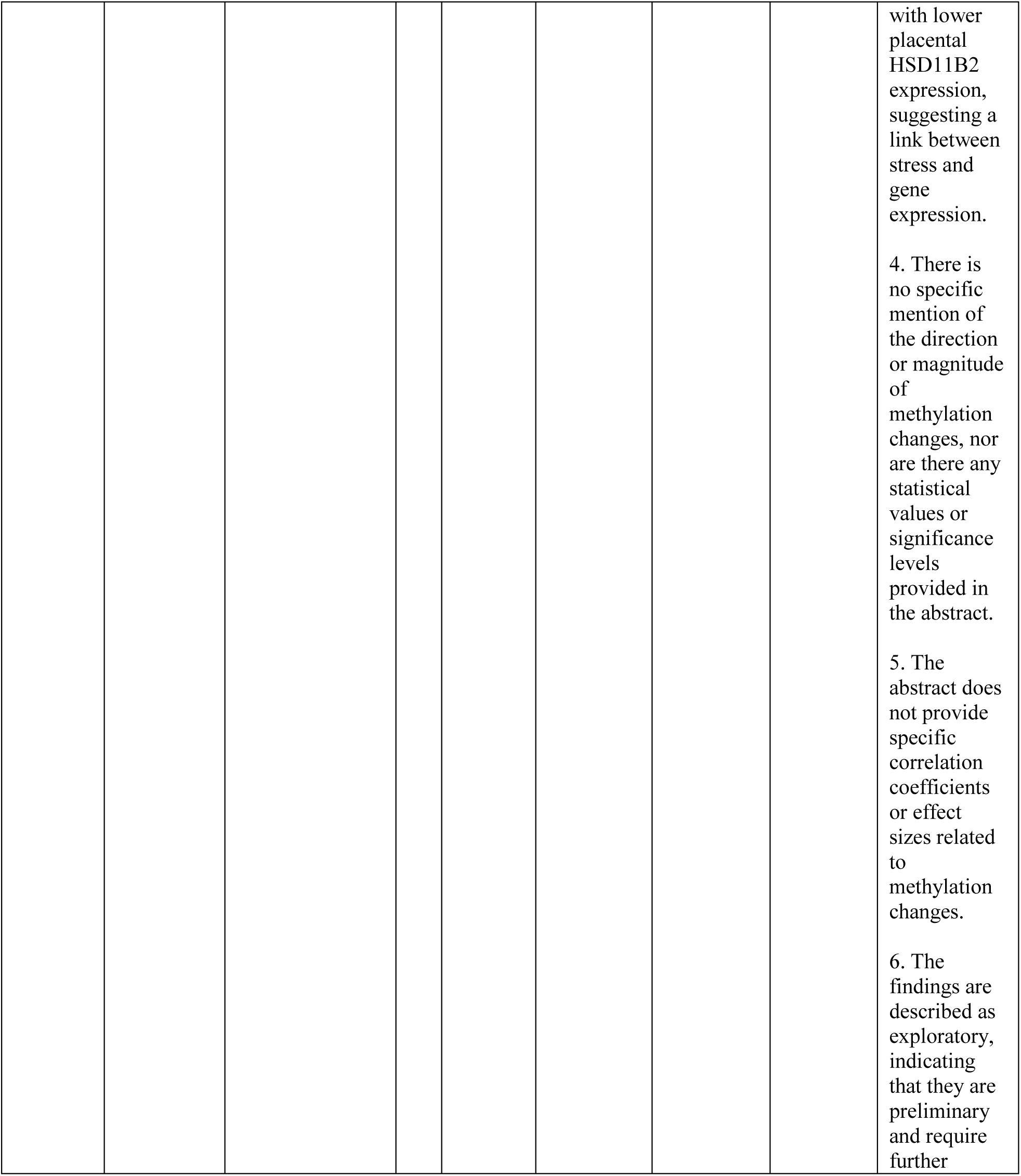
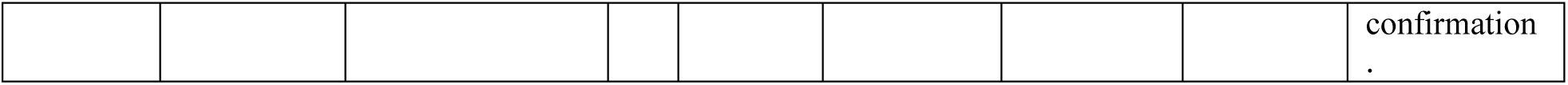

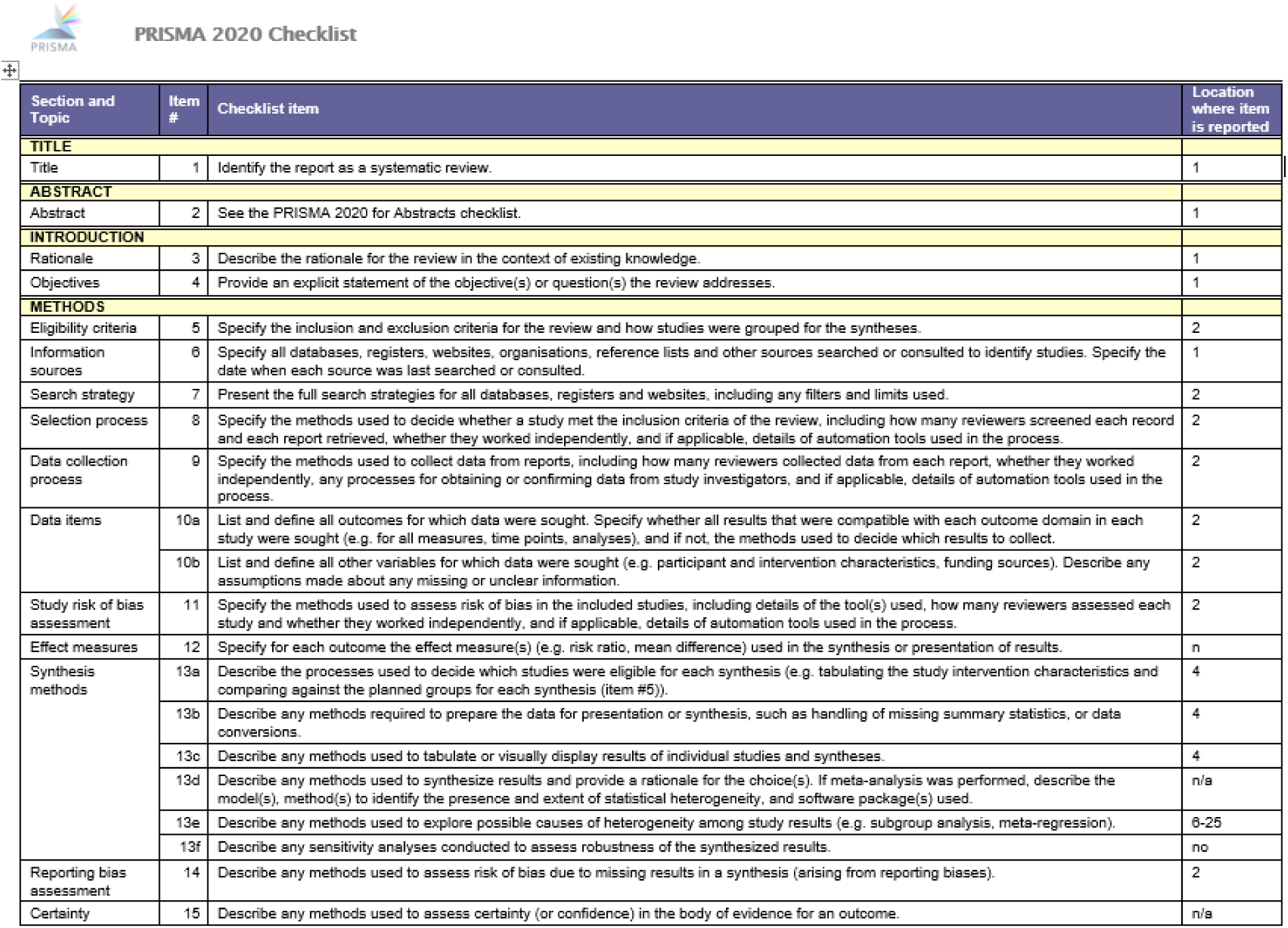
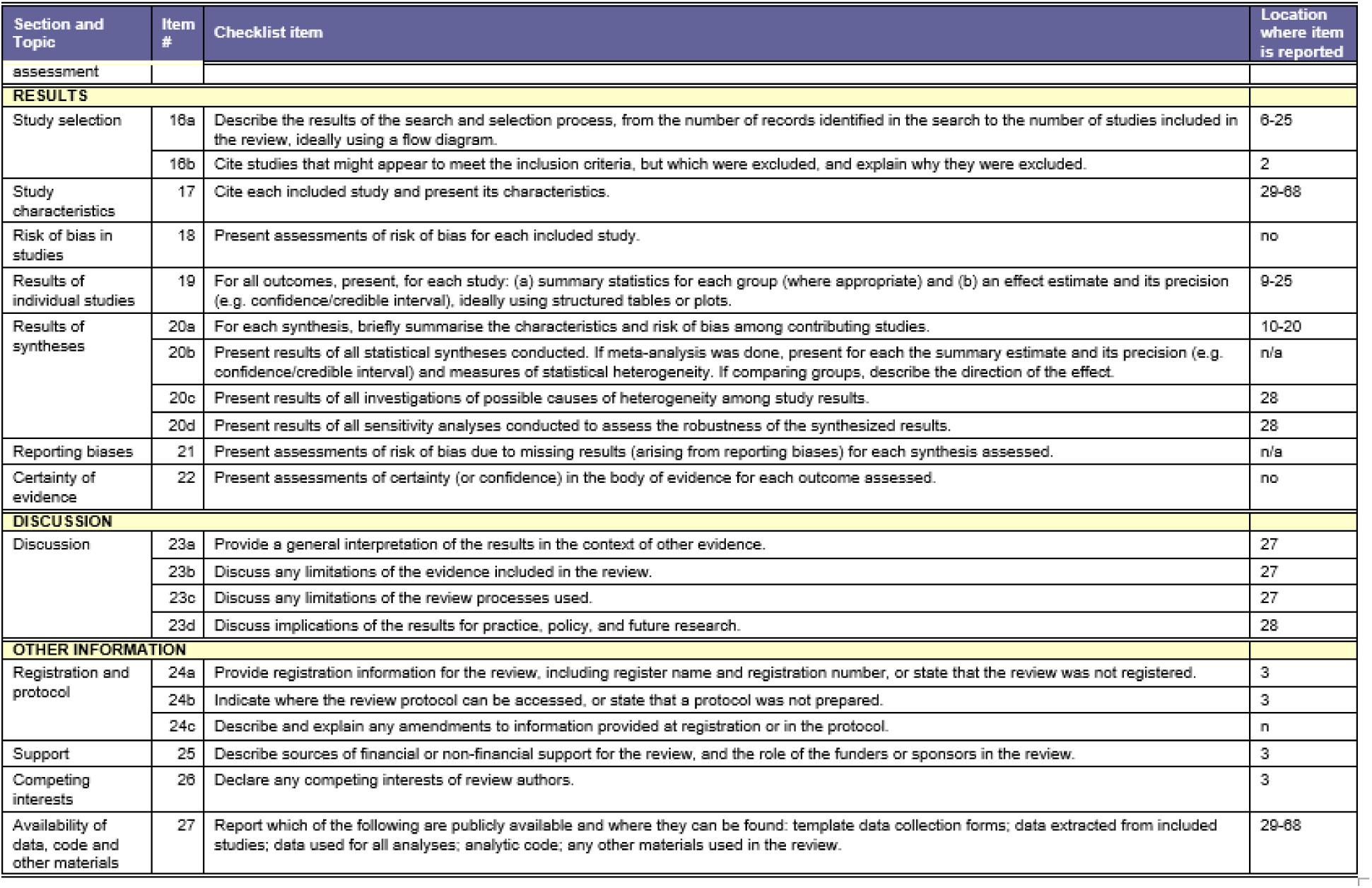

